# Adapting the BOADICEA breast and ovarian cancer risk models for the ethnically diverse UK population

**DOI:** 10.1101/2025.02.14.25322307

**Authors:** Lorenzo Ficorella, Xin Yang, Nasim Mavaddat, Tim Carver, Hend Hassan, Joe Dennis, Jonathan Tyrer, Weang-Kee Ho, Soo-Hwang Teo, Mikael Hartman, Jingmei Li, Mikael Eriksson, Kamila Czene, Per Hall, Tameera Rahman, Andrew Bacon, Steven Hardy, Adam E. Stokes, Francisca Stutzin Donoso, Stephanie Archer, Jacques Simard, Paul D. P. Pharoah, Juliet A. Usher-Smith, Marc Tischkowitz, Douglas F. Easton, Antonis C. Antoniou

## Abstract

**Background:** BOADICEA is a widely used algorithm for predicting breast and ovarian cancer risks, using a combination of genetic and lifestyle/environmental risk factors. However, it has largely been developed using data from individuals of White ethnicity.

**Methods:** We utilised data from multiple sources to derive estimates for the distributions of risk factors and their effect sizes in major UK ethnic groups (White, Black, South Asian, East Asian, and Mixed). We combined these with ethnicity-specific population cancer incidences to update BOADICEA so that it provides ethnicity-specific risk estimates. We also developed and included a method for deriving adjusted polygenic scores for individuals of mixed genetic ancestry.

**Results:** The predicted average absolute risks were smaller in all non-White ethnic groups than in Whites, and the risk distributions were narrower. The proportion of women classified as at moderate or high risk of breast or ovarian cancer, according to national guidelines, was considerably smaller in non-White women.

**Discussion:** The updated BOADICEA (v7), available in the CanRisk tool (www.canrisk.org), is based on estimates more appropriate for non-White women in the UK. Further validation of the model in prospective studies is required. Considering these findings, risk classification guidelines for non- White women may need to be revised.

## Background

Breast cancer (BC) is the most common cancer in women in the UK, with 55,920 new cases diagnosed and 11,499 deaths (annual averages, 2017-2019 (1)). Epithelial tubo-ovarian cancer (EOC) is less common but is associated with significant mortality, with 7,495 EOC cases diagnosed and 4,142 deaths in the UK (annual averages, 2017-2019 (1)). Surveillance (2–5) and prevention (6–11) options are available; however, applying them universally is impractical and expensive (12), and may be associated with adverse effects (13, 14). Personalised cancer risk assessment tools enable the classification of women into different risk categories, facilitating targeted screening and prophylaxis for those who would benefit most.

BOADICEA (the Breast and Ovarian Analysis of Disease Incidence and Carrier Estimation Algorithm) (15–17) incorporates multifactorial BC and EOC risk prediction models; it is available via the CanRisk tool (www.canrisk.org, (18)). The models predict future risks of BC and EOC by combining information on lifestyle/hormonal/reproductive risk factors (questionnaire-based risk factors, QRFs), rare moderate- and high-risk pathogenic variants (PVs) in cancer susceptibility genes, joint effects of common genetic variants summarised via polygenic scores (PGS), cancer family history (FH), mammographic density (MD), BC pathology information, and population cancer incidences. Like most risk prediction models, BOADICEA (BC model v6 and earlier; EOC model v2) was largely developed and validated using data from populations of White ethnicity/European ancestry (19–21). A key challenge in developing models for non-White ethnicities has been their under-representation in studies of both genetic and lifestyle/environmental risk factors (22, 23).

BC and EOC incidences vary among countries and across ethnic groups, even within the same country (24–26). The distributions of established lifestyle and hormonal risk factors also vary by ethnicity (27, 28). Ethnicity has also been associated with differences in breast tumour pathology; in particular, triple negative BC is more common in individuals of African descent (29–32). There are also important differences in the effects of genetic risk factors by population. BOADICEA incorporates a 313-SNP PGS for BC (33) and a 36-SNP PGS for EOC (34), developed using data from individuals of European ancestry (19, 33, 35–38). Although these PGS are associated with cancer risks for Asian and African ancestry women, the associations are attenuated; moreover, the PGS distributions vary by ancestry (34, 39–41). Therefore, to allow broad applicability to diverse populations, the BOADICEA models need to be adapted to account for these differences.

In the UK, 18.3% of the population self-identified as non-White in 2021 (42). Here, we describe our approach to adapting the BOADICEA BC and EOC risk prediction models to ethnically diverse populations (BC model v7, EOC model v3). This involved updating the population cancer incidences, relative risk (RRs), and joint risk factor distributions with ethnicity-specific estimates using a synthetic modelling approach. (15, 43). We also developed a novel approach for incorporating PGS for individuals of mixed genetic ancestry. We have focused specifically on developing a model applicable to the UK population, but the approach could be used to develop similar generalised adaptations of BOADICEA for other countries.

## Methods

### Underlying cancer models

In BOADICEA, the BC and EOC incidences are modelled as functions of the effects of QRFs, MD, PVs in cancer susceptibility genes, PGS, and a residual polygenic component modelling residual familial effects on cancer risks ((16, 17), Supplementary Materials). There are separate models for BC and EOC, developed using the same framework, which consider (partially) different cancer susceptibility genes.

The incidences of each cancer are assumed to be independent, conditional on the genotypes and QRFs in the model. The genotype- and QRF-specific incidences are calculated by constraining the total age- specific incidences to agree with the overall population incidences (15), which are birth cohort- and country-specific (17).

The models also incorporate data on breast tumour pathology, in the overall population and for PV carriers. For this purpose, tumours are classified into three subtypes: 1) oestrogen receptor (ER) positive; 2) triple negative (TN), i.e. ER, progesterone receptor, and human epidermal growth factor receptor 2 negative; and 3) ER-negative but not TN. The models use age-specific estimates of tumour subtype distributions, considering the known differences in subtype distributions between PV carriers and non-carriers. (16, 44–47).

For this BOADICEA extension, we examined and, if necessary, updated all parameters while preserving the underlying models structure.

### Ancestries and ethnicities considered

*Ethnicity* refers to a social group identity that is based on shared characteristics, such as cultural traditions, ancestry, language, religion, or social experiences (48) (Supplementary Materials). Since ethnicity could influence behavioural choices and risk factors, we used data on self-reported ethnicity to obtain risk estimates for lifestyle, hormonal, and reproductive risk factors. To adapt BOADICEA to the major ethnic groups in the UK, we adopted the ethnic categories employed in UK Biobank, which correspond to those collected in recent UK Censuses (42). We considered six broad ethnic groups: White (British, Irish, other White), South Asian (Bangladeshi, Indian, Pakistani, other South Mixed (White and Black African, White and Black Caribbean, White and Asian, other Mixed) and Other.

*Genetic ancestry* refers to the ’complex inheritance of one’s genetic material’ (49). Statistical methods identify groups of individuals with high (genetic) affinity; this allows measurement of the genetic similarity between populations and individuals (Supplementary Materials). Typically, genetic factors are driven by ancestry and related data are ancestry-specific. Therefore, for genetic risk factors, we based estimates on genetic ancestry rather than self-reported ethnicity. For this purpose, ancestry was classified as European, African, South Asian, East Asian, and Mixed.

### Datasets used in the modelling process

The study populations used to define the different model components are described in detail in the Supplementary Materials. We used data from UK Biobank(50) and the KARMA (51), MyBrCa (52), SGBCC (53), and BCSC (54) studies.

### Cancer incidences and tumour subtype distributions

#### Ethnicity-specific population cancer incidences

We considered breast, ovarian, and pancreatic cancer incidences for women and pancreatic and prostate cancer incidences for men. Due to data limitations, we could not derive ethnicity-specific incidences for male BC, and these were assumed to be independent of ethnicity. We derived ethnicity- specific cancer incidences for White, Black, Mixed, and East/South Asian UK populations; due to sample size limitations, South and East Asians were grouped together.

For each cancer type, we used published incidence rate ratios by ethnic group relative to Whites, based on Public Health England data (2013–2017) (25). These ratios were then re-standardised to obtain incidence rate ratios relative to the whole UK population. The proportion of each ethnic group in the UK population (**Table S1**) was estimated from the Office for National Statistics (ONS, (55)); the re-standardisation procedure is described in the Supplementary Materials. We applied these incidence rate ratios to the UK population cancer incidences currently used in BOADICEA (16), thus generating ethnicity- and birth cohort-specific population cancer incidences. This approach ensures that the models remain internally consistent and backward compatible when ethnicity information is missing.

#### Gene- and ethnicity-specific tumour subtype distributions

We used published estimates from NCRAS (National Cancer Registration and Analysis Service) data (56, 57) on the distribution, by ethnicity and age at cancer diagnosis, of the three BC subtypes included in BOADICEA (Supplementary Materials). We derived age-, ethnicity and gene-specific subtype proportions for PV carriers by combining those estimates with published ORs for PV carriers and the respective age-interactions for each subtype (47).

### Genetic components

#### Polygenic Scores

##### PGS models

Given a set of SNPs, a PGS (*R_PGS_*) for a given individual is of the form:

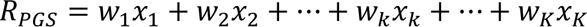

where *x_k_* is the allele dosage for SNP *k*, *w_k_* is the corresponding weight and K is the total number of SNPs in the set. The weights are assumed to be the same for each ancestry so that the same raw PGS would be calculated regardless of the individual’s ancestry. In the results section, ‘PGS’ refers to this raw score.

A ‘PGS model’ here is defined by the list of SNPs, their corresponding weights and ancestry-specific frequencies, and the mean (*μ*_*i*_) and standard deviation (*σ*_*i*_) in each genetically defined ancestry group (*i*). For each individual, *μ*_*i*_ and *σ*_*i*_ are used to obtain the normalised PGS (*Z*_*i*_), which depends on ancestry *i* even though the raw PGS is the same:

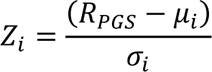

The PGS models we focused on here were derived from validated breast and ovarian cancer PGS models already incorporated into BOADICEA. However, the algorithm can be adapted to other PGS (58).

The BC PGS models were based on the 313-SNP PGS (33) and ancestry-specific parameter estimates from UK Biobank. Imputed genotypes were available in UK Biobank for 309 of 313 SNPs. Therefore, we constructed a 309-SNP PGS model, using the same weights as in the 313-SNP PGS model but excluding the 4 unavailable SNPs. SNP frequencies were estimated for all ancestries. We also constructed a 307-SNP PGS model by excluding two further SNPs that are correlated with the *CHEK2* PTV c.1100del (22_29203724_C_T and 22_29551872_A_G) (58). Parameters for the 313-SNP, 309- SNP, and 307-SNP PGS models were also estimated using Asian ancestry data from the MyBrCa (52) and SGBCC (53) studies, together with those for a 303-SNP PGS that included only SNPs with an imputation accuracy of >0.5 in both Europeans and Asians.

For EOC, we used the 36-SNP PGS model, developed by Dareng *et al.* (34), that is already included in the BOADICEA EOC risk model (17); SNP frequencies were estimated for all ancestries using UK Biobank data.

##### Use of PGS in BOADICEA

The PGS is incorporated into BOADICEA by partitioning the total (normalised) polygenic component into the sum of a known component, measured by the normalised PGS (*Z)*, and an unmeasured residual component (15):

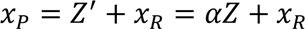

*α*^2^represents the proportion of the model polygenic variance attributable to the PGS, which can be ancestry-specific. *x_R_* is normally distributed with mean 0 and variance 1 − *α*^2^. **Figure 1** summarises the workflow for generating the final PGS models implemented in the updated BOADICEA.

**Figure 1.**
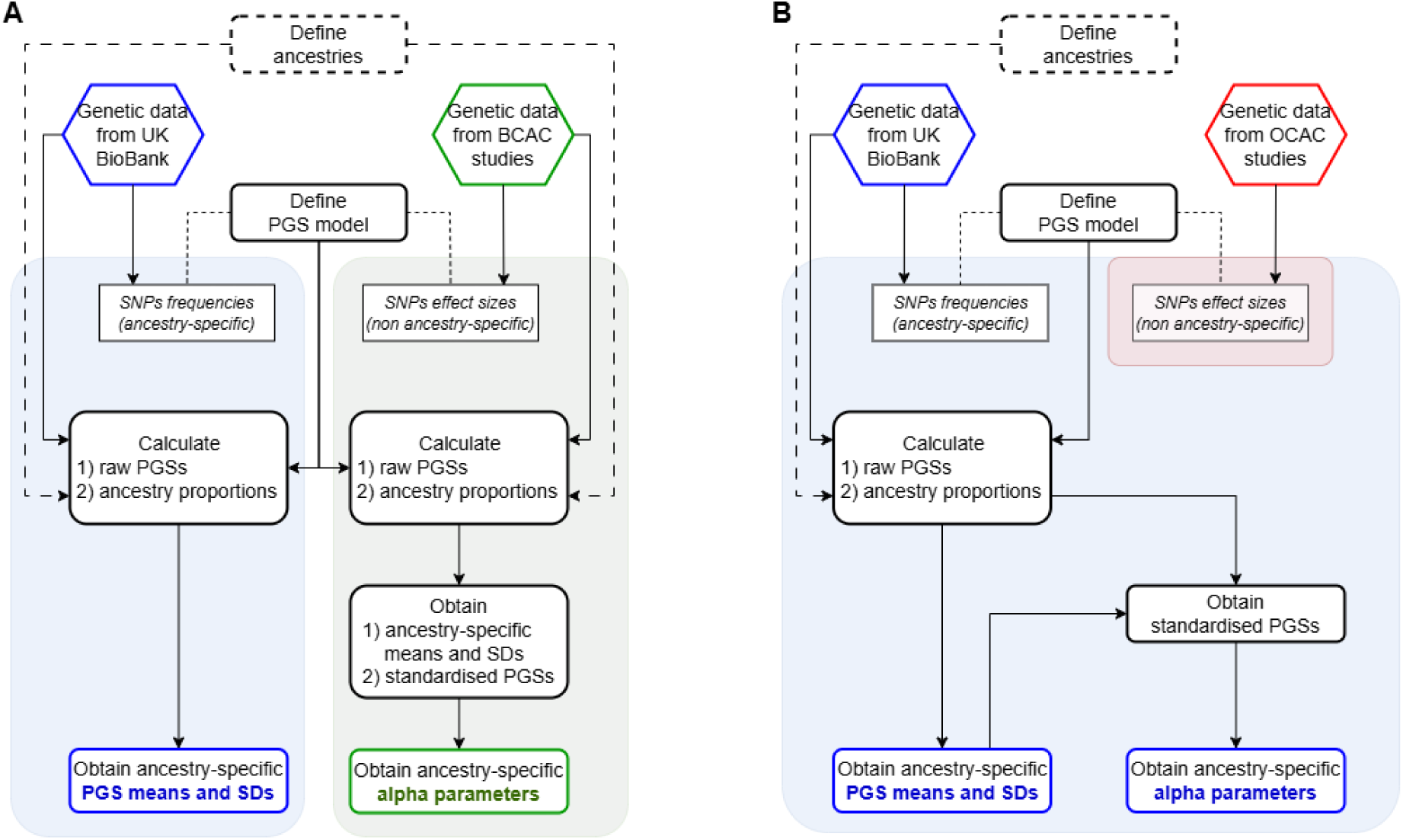
Process followed for defining the ancestry-specific PGS models, for BC (panel A) and EOC (panel B). Blue-shaded area: workflow and parameters derived from UK BioBank data. Green-shaded area: workflow and parameters derived from ‘BCAC data’. Red-shaded area: parameters derived from published OCAC (Ovarian Cancer Association Consortium) studies (34). Outputs: ancestry-specific means, standard deviations (SDs), and proportions of the polygenic variance explained (*alpha*).

For BC, a retrospective likelihood (RL) method was used to estimate the ancestry-specific *alphas* (*α*_*i*_) for South and East Asian women, using data from the MyBrCa (52) and SGBCC (53) studies. This approach models the probability of observing the PGS conditional on the phenotypes of the individuals (age of diagnosis and case/control status, Supplementary Materials and (58)). Comparable data were not available for Africans; therefore, we estimated *α* indirectly from three published estimates of the OR per standard deviation (SD) (59–61), assuming that *α* was proportional to the log- odds ratio and using the effect sizes in East Asians as the comparison (Supplementary Materials). For Europeans, we calculated *α* using published estimates (33) and procedures to incorporate alternative PGS into BOADICEA (58). Data used for calculating *alphas* (for BC PGS models) are referred to collectively as ’Breast Cancer Association Consortium (BCAC) data’.

For EOC, we first calculated the variance explained by the PGS using ancestry-specific allele frequencies estimated from UK Biobank and published log-odds ratios estimated from OCAC (34). These variances were then used to calculate the corresponding *α*_*i*_ parameters (Supplementary Materials). For Europeans, we used published *α* estimates (17) already implemented in CanRisk.

##### PGS for individuals of mixed genetic ancestry

The ancestry-specific *alphas (α*_*i*_), and normalised polygenic scores (*Z*_*i*_) assume that each individual can be assigned unequivocally to one of the ancestry groups. We consider individuals to be ‘single-ancestry’ if the estimated proportion of their genome derived from their main ancestry is ≥0.8 (0.7 for South Asian); otherwise, we consider them as ‘Mixed’. For a ‘Mixed’ individual, corrected normalised score (*Z*) and overall *alpha* (*α*) can be computed if the estimates of the proportions of the genome attributable to each ancestry (*P*_*i*_) are known. *Z* and *α* are given by:

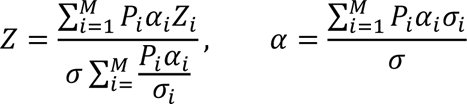

where *㲳*_*i*_ *σ*_*i*_, *α*_*i*_are ancestry-specific parameters, *Z*_*i*_is the polygenic scores normalised using the ancestry-specific *μ*_*i*_ and *σ*_*i*_, *P*_*i*_ is the ancestry proportion for ancestry *i*, and M is the number of ancestry groups considered (here, 4). The variance term *σ*^2^is given by

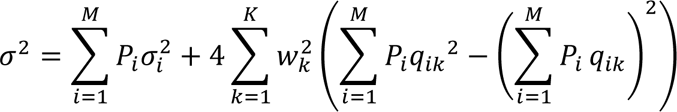

where *q_ik_* is the ancestry-specific frequency of allele *k* and *w*_*k*_ is its corresponding weight. The variance depends on the proportion of each ancestry in the individual but also accounts for the fact that SNP frequencies vary among ancestries (Supplementary Materials for derivations).

#### Rare genetic variants

No systematic reviews are currently available on the risks and frequencies of rare genetic variants associated with BC or EOC across multiple ethnicities/ancestries. Therefore, we conducted a literature search using PubMed in October 2024. The keywords used in the search can be found in Supplementary Materials. We manually reviewed articles and selected those that reported either RRs (or ORs) for the associations of germline PVs with primary BC or EOC, and/or PV frequencies in different populations.

### Lifestyle, hormonal, and reproductive risk factors

A recent scoping review (22) examined the associations of BC QRFs by ethnicity and concluded that there was no convincing evidence of differences in RRs among different ethnic groups for the QRFs included in BOADICEA. This was also confirmed in other systematic reviews, e.g. on Asian women (62). Given that the largest studies and most robust studies to date have been conducted in White populations (22), we assumed that the RRs currently used in the BC model (16) apply to all ethnic groups. Similarly, previous studies have found no clear evidence for differences in RRs for EOC across different ethnic groups, for the QRFs in BOADICEA (63–68). We therefore assumed that the RRs currently used in the EOC model are also applicable to all ethnic groups(17).

However, the distributions of these QRFs vary across different ethnicities (62). We used data from UK Biobank to estimate the BC and EOC QRFs distributions for each ethnic group. It is well recognised that UK Biobank is a highly selected cohort and may not be representative of the entire UK population. To assess the impact of using the UK Biobank QRFs distributions on the predicted risks we compared the 10-year BC risk predicted in the KARolinska Mammography Project for Risk Prediction of Breast Cancer (KARMA) cohort using 1) the default QRFs distributions in BOADICEA, which were estimated from large epidemiological studies (15), and 2) the QRFs distributions in the White populations in UK Biobank.

### Mammographic Density

BOADICEA incorporates MD, assessed using the four-category BIRADS scale (Breast Imaging Reporting & Data System, version 4) (15). We used the public version of the BCSC (Breast Cancer Surveillance Consortium) risk estimation dataset (54) to estimate the ORs and distribution of BIRADS categories for each ethnic group (Supplementary Materials and **Tables S2, S3**).

### Assessing risk stratification

We used the final models to assess the BC and EOC risk distributions by ethnicity. Risks were calculated for hypothetical UK women considering all possible combinations of risk predictors (QRFs, MD and PGS); distributions were then obtained by considering the frequency of each combination in the reference population of each model. We then estimated the proportion of women in different risk categories, given the risk factors studied. For BC, we used the risk categories employed in the NICE guidelines (8): population (<17% lifetime risk), moderate (17%-30%), and high (>30%). For EOC, NICE guidelines (11, 69) set a single threshold (5% lifetime risk) for recommending risk-reducing surgery. We considered this threshold and an additional intermediate threshold of 3.5% (21).

## Results

### Cancer incidences and tumour subtype distributions

#### Ethnicity-specific population cancer incidences

**Table 1** shows the incidence rate ratios for each ethnic group relative to the whole UK population, for the four cancers considered in BOADICEA. **Figure S1** shows the corresponding population cancer incidences for the 1980-1989 birth cohort, by age and ethnicity. The incidences of all cancer types are lower in each of the non-White ethnicities than in Whites, except for pancreatic and prostate cancer incidences for Blacks. The largest difference is for prostate cancer incidence, which is approximately twice as high in Black men but half as high in Asian men compared to the UK male population overall.

**Table 1.**
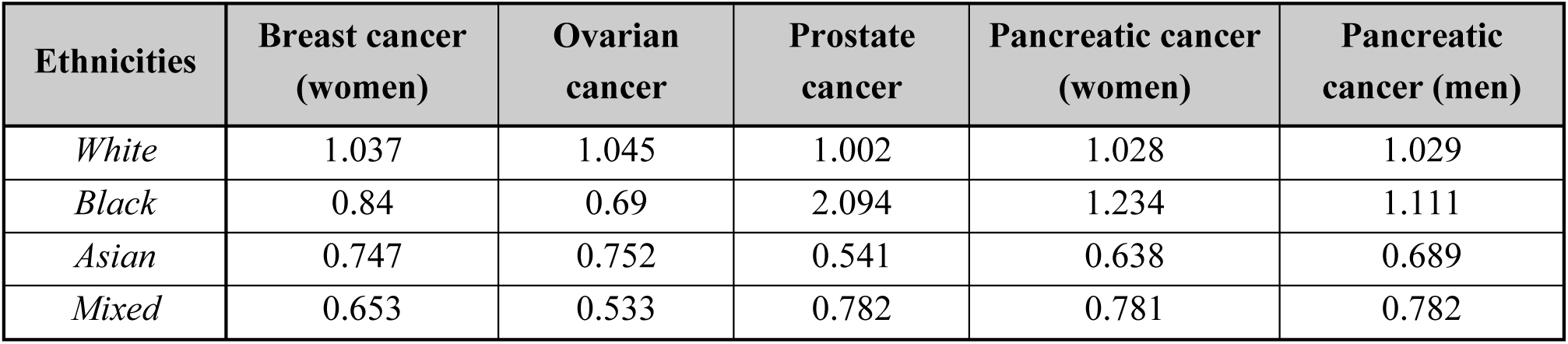
Incidence rate ratios by ethnic group and cancer type, relative to UK population (assuming 2016 census ethnicity distributions). South and East Asian ethnicities were grouped into a single Asian group due to small numbers.

| **Ethnicities** | **Breast cancer**<br>**(women)** | **Ovarian**<br>**cancer** | **Prostate**<br>**cancer** | **Pancreatic cancer**<br>**(women)** | **Pancreatic**<br>**cancer (men)** |
| --- | --- | --- | --- | --- | --- |
| *White* | 1.037 | 1.045 | 1.002 | 1.028 | 1.029 |
| *Black* | 0.84 | 0.69 | 2.094 | 1.234 | 1.111 |
| *Asian* | 0.747 | 0.752 | 0.541 | 0.638 | 0.689 |
| *Mixed* | 0.653 | 0.533 | 0.782 | 0.781 | 0.782 |

#### Gene- and ethnicity-specific tumour subtype distributions

The distributions of breast tumour subtypes by age at cancer diagnosis and ethnicity, based on published estimates from NCRAS data (56, 57), are shown in **Figures S2-S4**. The distributions of tumour subtypes in the general population varied by ethnicity, with higher proportions of ER-negative and TN breast cancers in non-Whites (particularly Black women, Supplementary Materials). The derived age- and gene-specific subtype proportions for PV carriers by ethnicity are shown in **Tables S4-S6** and **Figures S5-S7**; differences between ethnicities were particularly marked for *BRCA1, BRCA2, PALB2, BARD1, RAD51C*, and *RAD51D* carriers.

### Genetic components

#### Polygenic Scores

##### PGS distribution in European and non-European individuals from UK Biobank

**Tables S7-S10** contain the SNP weights (*w*_*k*_) and ancestry-specific allele frequencies (*q*_*ik*_) for the BC PGS models (**S7** and **S8**, respectively), along with the ancestry-specific means and SDs (**S9** and **S10**). **Table S11** reports the ancestry distributions in UK Biobank participants. Since the PGS means estimated in women and men were similar, we used the parameter estimates based on the combined sample of men and women.

For both 309-SNP and 307-SNP PGS models, the mean PGS was highest in African individuals, followed by South and East Asians, and lowest in Europeans. ‘Mixed’ had intermediate means, similar to the South Asians. The SDs were generally similar, but slightly higher among Europeans than other populations and highest among the ‘Mixed’ individuals.

**Table S12** contains the *w*_*k*_ and *q*_*ik*_ parameters for the EOC PGS model, and **Table S13** contains the corresponding ancestry-specific means and SDs. The mean EOC PGS was highest among European individuals, and similar among individuals of African, South Asian, and East Asian ancestry; the SD was also higher for European ancestry individuals than other ancestries.

##### Association between PGS and cancer risk, and proportion of polygenic variance explained

**Table S14** shows the parameters for the different BC PGS models by ancestry, estimated using ‘BCAC data’. The means and SDs of the BC 309-SNP and 307-SNP PGS were very similar to those estimated in the corresponding ancestry groups in UK Biobank above.

The OR per 1 SD associated with the 309-SNP PGS model was slightly lower in both South and East Asian women (1.49 and 1.53 respectively) than that reported for European women (OR=1.64). The corresponding *alphas* (α_RL_) were markedly lower in South and East Asian ancestry women (0.326 and 0.331 respectively) than the corresponding European parameter (0.50). Very similar α_RL_ estimates were obtained for the other PGS. Removing the two *CHEK2*-correlated SNPs resulted in large differences in the PGS mean. For African women, we indirectly estimated α to be ∼0.196 for all the BC PGS models. For the EOC PGS model, the estimated α was highest in Europeans (α=0.223) and lowest for East Asians (0.171) (**Table S13**).

##### PGS for individuals of mixed genetic ancestry

Figure S8 shows the distribution of *σ*, the standard deviation of the raw PGS, calculated over a range of combinations of ancestry proportions: *σ* is greater than the weighted average of the ancestry-specific standard deviations *σ*_*i*_, and the discrepancy is more evident when no ancestry dominates (i.e. when two or more ancestries have similar proportions). As a result, for ‘Mixed’ individuals, the normalised PGS and *α* are smaller than the estimates obtained by calculating a simple weighted average of the ancestry-specific values. The effect of the PGS on the risk estimates is therefore attenuated, relative to the predictions assuming a simple weighted average (**Figure S9**).

Figure 2 outlines the steps needed to include PGS in BOADICEA risk calculations, for individuals of single or ‘Mixed’ ancestry. **Table 2** summarises the corresponding ancestry-specific parameters (*σ*_*i*_, *σ*_*i*_ and *α*_*i*_) used in the models.

**Figure 2.**
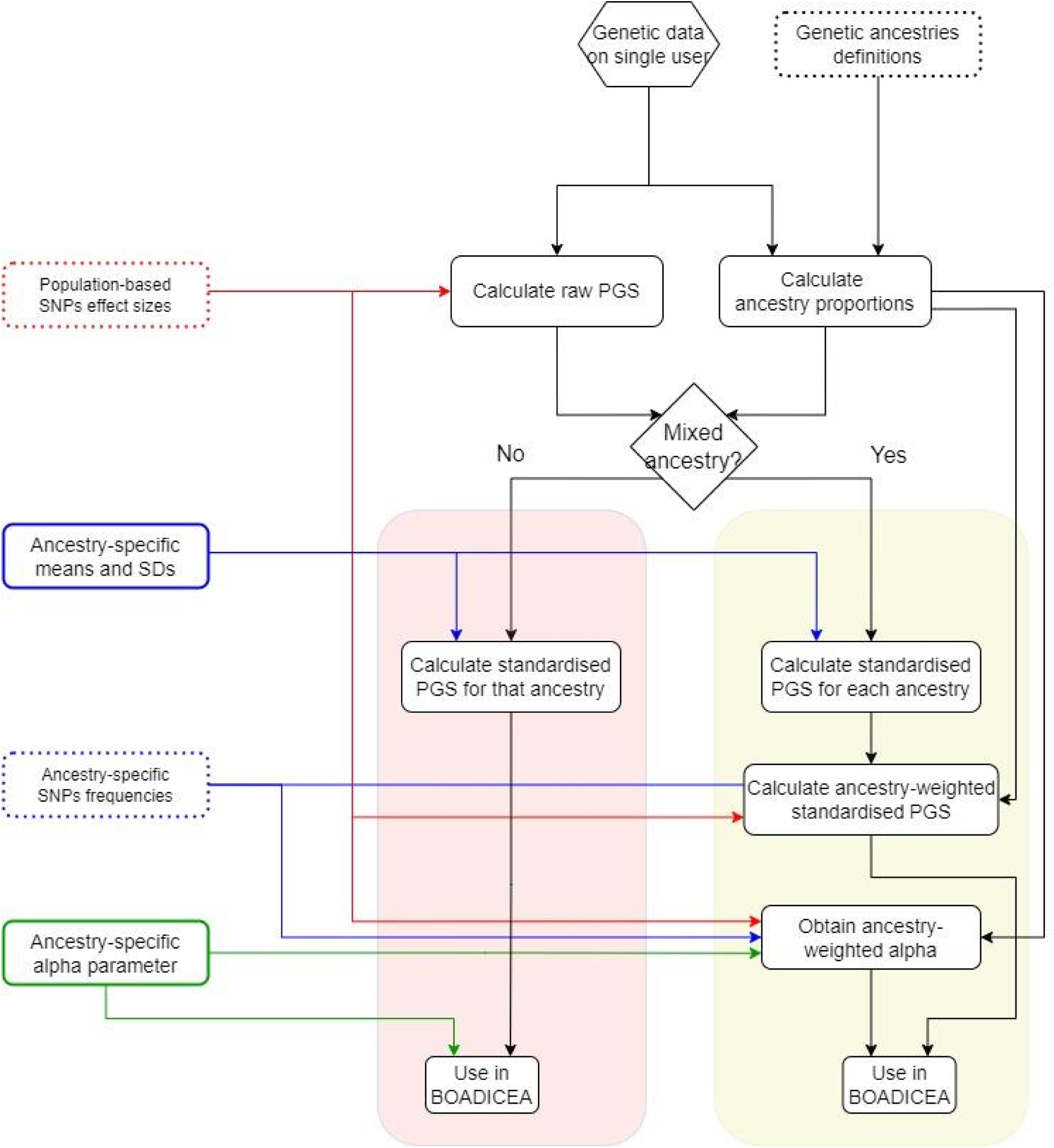
Workflow to be followed for employing ancestry-specific PGS in BOADICEA, in single- ancestry individuals (red-shaded area) and ‘Mixed’ ones (yellow-shaded area). Red lines: population- based parameters of the PGS model. Blue lines: ancestry-specific parameters of the PGS from UK BioBank. Green lines: ancestry-specific *alphas* (α_i_) from ‘BCAC data’ (BC) or UK BioBank (EOC).

**Table 2.** Summary of ancestry-specific parameters required for standardising the raw PGS in single- ancestry individuals, and for calculating the derived parameters in ‘Mixed’ individuals. For unspecified/unknown ancestry, or other ancestries outside the listed ones, we employed the European PGS parameters.

| Ancestries | Breast cancer |  |  |  |  |  | Ovarian cancer |  |  |
| --- | --- | --- | --- | --- | --- | --- | --- | --- | --- |
|  | 309-SNP PGS |  |  | 307-SNP PGS |  |  | 36-SNP PGS |  |  |
| | *Mean*<br>$\mu$ | *SD*<br>$\sigma$ | *Alpha*<br>$\alpha$ | *Mean*<br>$\mu$ | *SD*<br>$\sigma$ | *Alpha*<br>$\alpha$ | *Mean*<br>$\mu$ | *SD*<br>$\sigma$ | *Alpha*<br>$\alpha$ |
| *European* | -0.335 | 0.615 | 0.499 | -0.005 | 0.613 | 0.497 | 0.007 | 0.322 | 0.223 |
| *African* | 0.292 | 0.547 | 0.196 | 0.634 | 0.547 | 0.196 | -0.152 | 0.254 | 0.178 |
| *East Asian* | 0.268 | 0.555 | 0.326 | 0.611 | 0.555 | 0.326 | -0.161 | 0.235 | 0.171 |
| *South Asian* | -0.160 | 0.602 | 0.331 | 0.179 | 0.601 | 0.329 | -0.180 | 0.278 | 0.194 |

#### Rare genetic variants

Five large studies have examined the associations between rare PVs and BC risk in individuals from different ethnicities/ancestries (Supplementary Materials) (70–74). There were no significant differences in the BC OR estimates among ancestries for any of the genes in any of the studies **(Table S16)**. Similarly, two studies have assessed the associations between germline PVs and EOC risk in non-European populations (75, 76) and showed no significant differences in the ancestry-specific EOC ORs estimates. For instance, *Ho et al., 2024* estimated OR=39.8 (95% CI: 29.6–53.3) and OR=6.8 (95% CI: 3.9–11.9) in Asian women, for *BRCA1* and *BRCA2* respectively (76). We therefore assumed that the RRs for both BC and EOC associated with PV susceptibility genes, as previously implemented in BOADICEA for European women (16), are also applicable to non-European ancestries.

**Table S17** shows that the estimated PV allele frequencies were similar across ancestry groups except for *CHEK2*, where the overall PV frequency was lower among non-Europeans than in Europeans. This difference is driven by the c.1100del variant, which accounts for the majority of PV carriers in Western European populations but is much rarer in non-Europeans (70). The overall *CHEK2* PV frequency in non-Europeans was estimated to be 0.00109 (Supplementary Materials). The PV frequencies for the other susceptibility genes were assumed to be the same across all ancestries and equal to those previously assumed in BOADICEA (16).

### Lifestyle, hormonal, and reproductive risk factors

We estimated QRFs distributions using baseline data on 250,739 women in UK Biobank, including 235,843 Whites, 4,352 Blacks, 3,510 South Asians, 937 East Asians, 1,765 Mixed, and 4,332 of other or unknown ethnicity (**Table S18**). A comparison of the 10-year BC predicted risks in the KARMA study, using the default QRFs distributions in BOADICEA (16) with those assuming the estimated distributions in UK Biobank Whites showed only marginal differences in absolute risk predictions the risk distributions were similar, that the default QRFs distributions in BOADICEA (validated in Whites) were based on multiple large studies, and to maintain consistency with the previous BOADICEA model for Whites, we applied the current default distributions to Whites and individuals of other/unknown ethnicity. The estimated QRF distributions from UK Biobank were incorporated for Black, South Asian, East Asian, and Mixed ethnicities.

### Mammographic Density

We used the imputed BCSC dataset to estimate ORs for the associations of MD with BC risk by ethnicity and age category (**Table S19**). The OR estimates for Asian, Black, and Mixed ethnicities were associated with wide confidence intervals. Models that included an interaction term between ethnicity and MD (**Table S20**) did not fit significantly better: likelihood-ratio test (LRT) p-value=0.12 in the <50 years age group; LRT p-value=0.053 in the ≥ 50 years group. We therefore assumed that the RRs previously incorporated in BOADICEA could be applied to all ethnic groups.

The ethnicity-specific BIRADS distributions were estimated using the imputed BCSC dataset, (**Tables S18** and **S21**). The distributions differ by ethnicity in both the <50 and ≥50 years age groups. (Pearson’s Chi-squared test, p-value <10^-5^). MD was highest in Asians (e.g. category D was 1.7-1.8 times more frequent than in Whites, in both age groups) and lowest in Black women (e.g. category A was 1.1-1.3 times more frequent than in Whites).

### Multifactorial models

#### Implications for risk stratification

**Figures S11** and **S12** show the cumulative BC and EOC risks for PV carriers. The estimated risks are highest for White ethnicity PV carriers. For example, the BC risk by age 80 ranged from 47% for Mixed to 50% for Asian, 54% for Black, and 58% for White *BRCA2* PV carriers. Similarly, the corresponding EOC risks were 8% for Mixed, 10% for Black, 11% for Asian, and 15% for White *BRCA2* PV carriers.

Figure 3 shows the distribution of lifetime BC and EOC risks for women of different ethnicities, considering all risk predictors available in the models; ethnicity-specific parameters are summarised in **Table S22**. The distributions of BC and EOC risks were widest for White women; all other ethnicities had similar narrower ranges and lower median risks. Supplementary **Figures S13**-**S20** further show the distribution of lifetime and 10-year BC and EOC risks by ethnicity and by predictor combinations; **Figure S21** shows the distribution of lifetime BC risk in BRCA2 carrier women. These figures show that the wider risk distribution in Whites is primarily driven by the higher effect size of the PGS (*α*_*i*_) whereas the lower median risks in non-Whites are mainly driven by the ethnicity- specific incidences.

**Figure 3.**
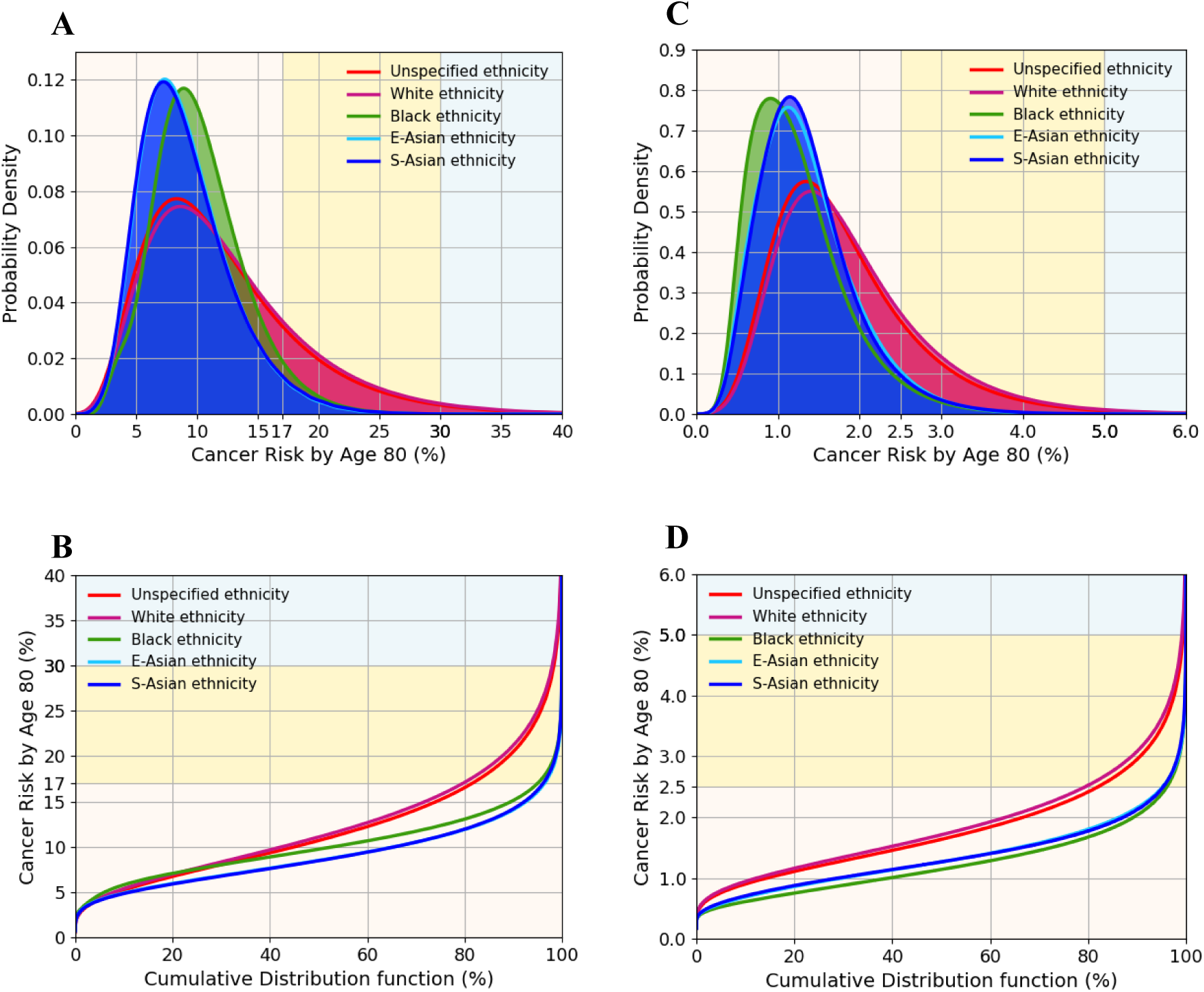
Predicted BC and EOC lifetime (age 20-80 years) risks for women of different ethnicities, untested for PVs and unknown family history; risks calculated using all risk predictors: PGS, QRFs, and MD (for BC model only). (A,B) BC risk; backgrounds are shaded to indicate three risk categories: <17% (light yellow); 17% - 30% (yellow); ≥ 30% (light blue). (C,D) EOC risk; backgrounds are shaded to indicate three risk categories: < 2.5% (light yellow); 2.5% - 5% (yellow); ≥ 5% (light blue). (A,C) probability density function against absolute risk; (B,D) absolute risk against cumulative distribution. BC, breast cancer; EOC, epithelial ovarian cancer; PGS, polygenic score; QRFs, questionnaire-based risk factors; MD, mammographic density.

**Table 3** and **Table S23** show the proportions of women in different BC and EOC risk categories, based on lifetime absolute risk thresholds (unknown FH and mother with BC/EOC cancer at 50 years, respectively). **Tables S24** and **S25** display the corresponding proportions for 10-year BC risk. The proportions of women at high or moderate risk of BC and EOC were markedly smaller among non- White ethnic groups, regardless of the predictors used. For example, using data on PGS, QRFs, and MD (unknown FH), 79.9% of White women were in the population-level category for BC, 18.2% in the moderate-risk category, and 1.86% in the high-risk category. In contrast, 95-96% of non-White women were in the population-level, 3-4% in the moderate-risk, and only 0.01% in the high-risk category. Similar patterns were seen for EOC lifetime risk.

**Table 3.**
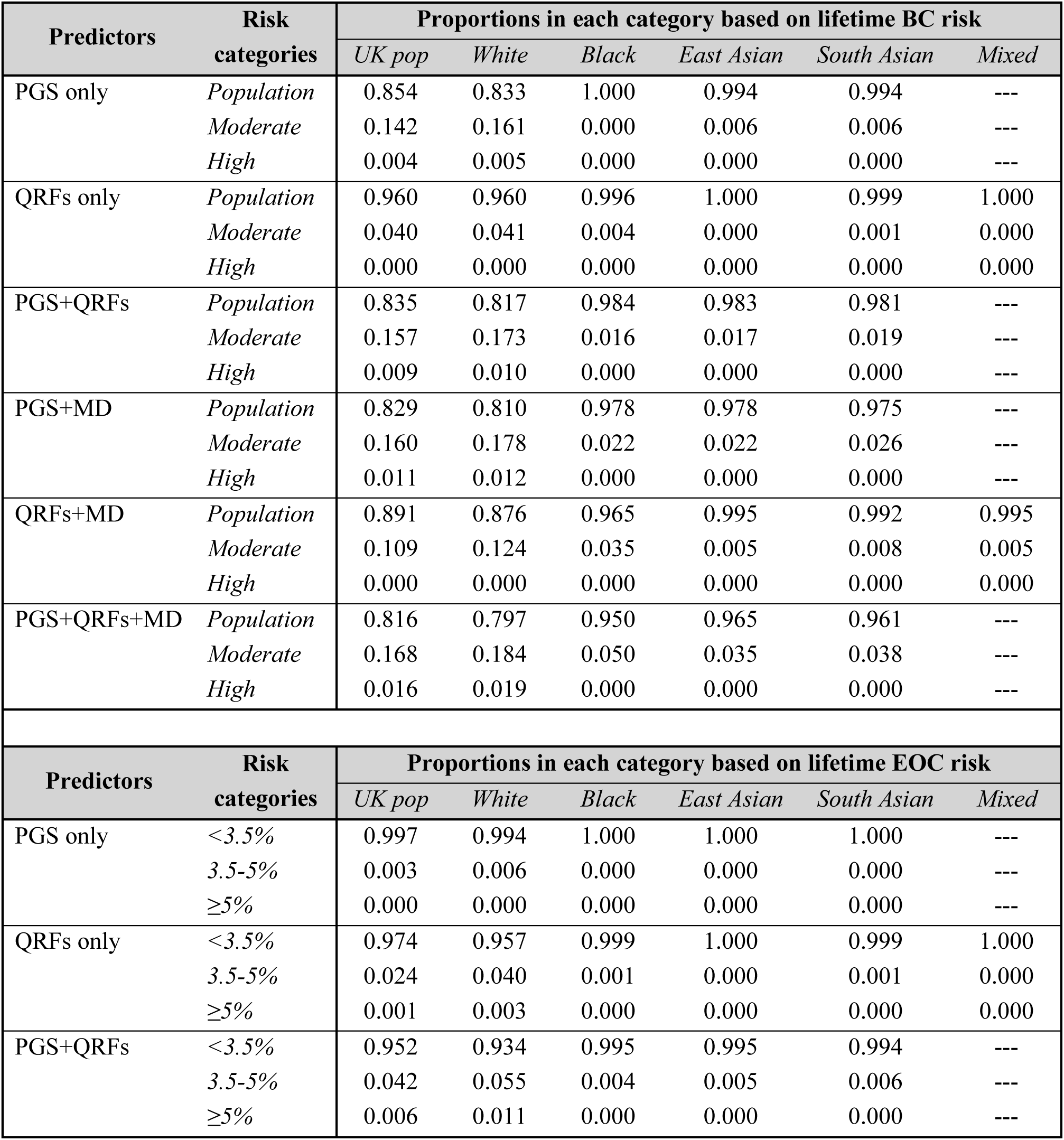
Proportion of women falling into different risk categories depending on their lifetime cancer risk (age 20 to 80 years) and their ethnicity, using different risk predictors. Unknown genetic background, unknown FH. First panel: lifetime BC risk; the thresholds between risk categories are 17%, between ‘population’ and ‘moderate’, and 30%, between ‘moderate’ and ‘high’ (NICE guidelines (8)). Second panel: lifetime EOC risk, with thresholds at 3.5% and 5% (threshold for risk- reducing surgery, NICE guidelines (11)).

## Discussion

In this manuscript, we present the methodological framework we used to adapt the multifactorial BOADICEA BC and EOC risk prediction models to ethnically diverse populations. This is one of the first risk prediction models to be adapted for multiple ethnicities. While we derived and implemented these models in the context of ethnic groups in the UK, the same framework could be used for other countries, provided that risk factor parameters and incidences are available for the relevant ethnic groups. We also present a novel approach for adjusting the PGS parameters for individuals of mixed genetic ancestry, which corrects the variance in the PGS due to the variability of individual SNPs across ancestries.

A key challenge in adapting cancer risk prediction models is the lack of data on different predictors in diverse populations. Although such data are becoming more readily available, no single dataset contains the necessary information on all risk factors. Therefore, we used a synthetic approach, combining data from several sources to inform ethnicity-specific parameters. We previously demonstrated that this approach is robust and yields models that provide valid risk estimates for both BC and EOC (15–17, 19–21). The models described here have been designed to apply to the four broad ethnicities most common in the UK: White, Black, South Asian, and East Asian. Further work will be necessary to adapt the model for specific subpopulations within these broad categories, and for other minority groups (e.g., from the Middle East or North Africa). Our flexible implementation allows for the models to be easily updated as more data become available.

Age-specific cancer incidence data by ethnicity are not currently available in the UK. We derived ethnicity-specific incidences using published incidence rate ratios (IRRs), which compare the incidence in each ethnic group relative to Whites. The resulting BC and EOC incidences are lower for Black and Asian women; the differences in incidence by ethnicity are broadly similar to those reported in other Western countries (77). The ethnicity-specific IRRs were assumed to be independent of age. However, the differences in IRRs by ethnicity may be age-dependent, being less marked at younger ages; hence, differences may be less marked in more recent birth cohorts. If so, the model may underestimate the lifetime risks in younger non-White women and will need further adaptation. Nevertheless, published IRRs for the <65 and ≥65 years age groups were similar (25), in line with the model assumptions.

The narrower range of BC and EOC risks observed among women of Asian and African ancestry is primarily due to the lower discrimination provided by the PGS (developed in European ancestry datasets) in these ancestries. Incorporating ancestry-specific data in PGS development has been shown to improve discrimination (78–80), and larger GWAS including diverse populations should generate PGS with better discrimination in non-Europeans (81). Incorporating such PGS into BOADICEA should then provide a model with a broader range of predicted risks in non-White populations.

Another assumption in our analyses is that the overall polygenic variance in each model is the same across all ethnic groups. These variances were previously estimated using segregation analyses and based predominantly on data from White women (82–84). There are no comparable segregation analyses in other populations; however, data from case-control studies suggest that the RRs associated with a positive family history of BC or EOC (which determines the polygenic variance) are similar across ethnic groups (22), indicating that this is a plausible assumption.

Due to limited available sample sizes in the UK, the proportions of the polygenic variance explained by the PGS models (*alphas*) were determined using ancestry-specific PGS effect sizes from non-UK studies. We used data on South and East Asian women from Malaysia/Singapore (39, 53, 79) and published data on, primarily, African American women (59–61). Given the similarities in PGS distributions between these studies and UK Biobank, this seems a reasonable assumption. However, there may be important differences that are not accounted for (e.g., the parameters for women of Afro- Caribbean ancestry could differ from African American women) and parameters may need further adjustment as more data become available.

Published evidence provides no convincing evidence of differences in RRs for different QRFs across ethnicities (22). Papers reporting RRs in single countries or smaller geographical areas (85–87) differ greatly in the methods employed, complicating comparisons. We therefore made the plausible simplifying assumption that the RRs associated with QRFs were independent of ethnicity. However, there are marked differences in the QRFs distributions by ethnic group. For this implementation, we used data from UK Biobank, which may not fully represent the broader UK population due to healthy volunteer bias (88–90). Unfortunately, population-level data on ethnicity-specific distributions of lifestyle, reproductive, or hormonal risk factors are currently lacking. To assess the impact of this assumption on the risk predictions, we conducted a sensitivity analysis in which we replaced the default QRFs distributions in BOADICEA (15) with the UK Biobank distributions for Whites and predicted BC risks in the KARMA cohort (51). The predicted risks were very similar **(Figure S10)** suggesting that using UK Biobank data for QRFs distributions does not induce any substantial bias in the predictions.

For MD, published studies vary in the methodological approaches used (87, 91, 92). We used the BCSC cohort to model ethnicity-specific components, ensuring a consistent methodological approach across ethnic groups. Our analyses suggest that the ORs for the association are similar across ethnic groups in the BCSC dataset, consistent with findings from other populations, which reported no statistically significant differences in the estimates across ethnicities (93, 94). We therefore assumed the same RR associations with MD for all ethnicities. One limitation of the BCSC public dataset is that it only includes data on incident cancers within one year of the baseline mammogram; datasets with longer follow-up would be valuable to confirm the consistency of MD associations across ethnicities. Another limitation of the model described here is the assumption that the density distribution, as for the QRFs, is independent of age; this limitation has been addressed in a separate model extension based on continuous MD measures (95).

Our implementation uses both self-reported ethnicity (to determine QRFs, pathology, and cancer incidence) and genetically determined ancestry (for the PGS). In practice, ancestry informative markers are not always available, e.g. when sequencing technologies are used to capture the PGS SNPs (96, 97). In such cases, self-reported ethnicity can serve as a proxy since it is highly correlated with genetic ancestry (**Table S11**). However, genetic data should ideally be used to determine ancestry for more accurate PGS estimation in individuals of mixed genetic ancestry.

Due to the lower incidence rates, the predicted absolute BC and EOC risks for non-White women are on average lower than for White women; moreover, the distribution of risk, based on currently available risk factors, is narrower for non-White women. This shows that the previous versions of the models, calibrated for White women, resulted in substantial overestimation of risk if used for women of other ethnicities; the new version corrects this. Incorporating ethnicity-specific parameters substantially impacts risk stratification, with far fewer Asian and Black women classified as moderate- or high-risk of developing cancer based on existing clinical management thresholds (8, 11). This raises the question as to whether current thresholds, primarily based on White populations, may need adjustment for other ethnic groups. Moreover, it is important to highlight that BOADICEA does not currently predict development of subtype specific disease or prognosis and survival, outcomes which also differ by ethnicity.

In summary, we have extended the BOADICEA BC and EOC risk prediction models to better apply to the diverse UK population. These updated models (BC v7, EOC v3) have been implemented in the CanRisk user-friendly interface (www.canrisk.org (18)), allowing the specification of self-reported ethnicity and genetic ancestry to determine the relevant parameters. Future large prospective studies will be required to provide direct validation of the BC and EOC risk models in non-White populations.

## Additional Information

## Acknowledgements

This research has been conducted using data from UK Biobank, a major biomedical database (50, 98). We thank the investigators of the KARMA (51), MyBrCa (52), and SGBCC (53) studies. MYBRCA thanks study participants and research staff (particularly Patsy Ng, Nurhidayu Hassan, Yoon Sook-Yee, Daphne Lee, Lee Sheau Yee, Phuah Sze Yee and Norhashimah Hassan) for their contributions and commitment to this study. SGBCC thanks the participants and all research coordinators for their excellent help with recruitment, data and sample collection. We also thank the BCSC (54) participants, investigators, mammography facilities, and radiologists for the data they have provided for this study.

## Authors’ contributions

Conceptualisation: LF, XY, NM, FSD, SA, JS, PDPP, JAUS, MT, DFE and ACA; data collection: XY, NM, HH, WKH, SHT, MH, JL, ME, KC, PH, TR, AB, SH; data analysis: LF, XY, NM, HH, JD, JT; model development: LF, TC, AES; writing in the initial draft: LF, XY, NM, HH, DFE and ACA. Funding: JAUS, MT, DFE, ACA. All authors reviewed the manuscript, provided feedback and approved the final manuscript text.

## Data availability

The study uses primarily UK Biobank (www.ukbiobank.ac.uk) and BCSC data, which are available at www.bcsc-research.org. Data from the KARMA study are available upon request from Karolinska Institutet, through the MTA form available at karmastudy.org/contact/data-access/. MyBrCa and SGBCC data may be requested via the study investigators of via application to the BCAC data access committee (www.ccge.medschl.cam.ac.uk/breast-cancer-association-consortium-bcac/data-data-access).

All parameters added to BOADICEA (BC v7, EOC v3) are summarised in T**able S22**; parameters for calculating raw PGS and standardising it for single-ancestry individuals are available in Supplementary Tables.

Scripts and parameters for standardising the raw PGS for ‘Mixed’ individuals and deriving the appropriate overall *alpha* are available on GitHub (https://github.com/CCGE-BOADICEA/SHARE-PRScalculation).

