## Supplementary Materials for "Adapting the BOADICEA breast and ovarian cancer risk models for the ethnically diverse UK population"

#### Methods

##### Underlying cancer models

The incidences of breast cancer (BC) and epithelial tubo-ovarian cancer (EOC) for individual $i$ at age $t$, $\lambda^{(i)}(t)$, depend on the baseline incidence $\lambda_0(t)$ and on their underlying genotype:

$$\lambda^{(i)}(t) = \lambda_0(t) \exp\left( \sum_{\mu=1}^{N_{MG}+1} \left[ \left( \beta_\mu(t) + \sum_\rho \boldsymbol{\beta}_{RF\rho\mu}(t) \cdot \boldsymbol{z}_{RF\rho}^{(i)} \right) \prod_{\nu=1}^{\mu-1} \left[ \left(1 - G_\nu^{(i)}\right) G_\mu^{(i)} \right] \right] + \beta_{PG}(t)\, x_P^{(i)} \right)$$

Considering $N_{MG}$ major genes in the model, $G_\mu^{(i)}$ is the indicator variable for the presence of PV in a major gene $\mu$ ($\mu = 1, \ldots, N_{MG}$), and $\beta_\mu(t)$ represent the age-specific log-relative risks (log-RRs) associated with $\mu$ relative to the baseline incidence. $G_{N_{MG}+1}^{(i)} = 1$ for non-carriers of any PV. $x_P^{(i)}$ is the polygenotype for individual $i$, assumed normally distributed in the general population (mean 0, standard deviation 1); $\beta_{PG}(t)$ is the age-specific log-RR per standard deviation associated with the polygene, relative to the baseline incidence.

Given a questionnaire-based risk factor (QRF) $\rho$ and a major gene $\mu$, $\beta_{RF\rho\mu}(t)$ is the vector of age-specific log-RRs for $\rho$; $z_{RF\rho}^{(i)}$ is the corresponding vector of indicator variables (0 or 1) that indicate the category of QRF $\rho$ for individual $i$. The models assume that RRs associated with PVs are log-additive (multiplicative) with the RRs associated with QRFs and the polygenic component.

Regarding the major genes, the BC model includes the effects of PVs in *BRCA1*, *BRCA2*, *PALB2*, *CHEK2*, *ATM*, *BARD1*, *RAD51C* and *RAD51D* (1), whereas the EOC model includes the effects of PVs in *BRCA1*, *BRCA2*, *RAD51D*, *RAD51C*, *BRIP1* and *PALB2* (2). Both models also consider the associations of PVs in *BRCA1*, *BRCA2,* and *PALB2* with pancreatic and prostate cancer (1, 2).

##### Ancestries and ethnicities considered

*Genetic ancestry* refers to the 'complex inheritance of one’s genetic material' (3), 'an individual’s biological ancestors'. All individuals have ancestors originating from multiple different locations at different time depth; so, it is more accurate to refer to proportions of genetic ancestries (4) noting that, while groupings allow analytical simplification, genetic ancestry is essentially continuous (4). Statistical methods identify groups of individuals with high (genetic) affinity, even though these measures are sensitive to the choice and labelling of reference populations (5). This allows measuring the *genetic* similarity between populations and individuals.

*Ethnicity* refers to a social group identity that is based on shared characteristics, such as cultural traditions, ancestry, language, religion, or social experiences. Ethnicity may vary according to geographical regions and can traverse different social classes, but the shared social experiences of an ethnic group can have health impacts over time (4). Self-identified ethnicity could therefore influence behavioural choices and risk factors.

#### Datasets used in the modelling process

*UK Biobank*

The UK Biobank dataset (6, 7) was used to determine ancestry-specific PGS distributions and ethnicity-specific QRFs distributions. A total of 226,340 women and 200,073 men without a cancer diagnosis (self-reported or registry reported-, excluding non-melanoma skin cancers) prior to or within six months of the start of follow-up were included in the analyses. Recruitment of volunteers took place between 2006 and 2010, and the resource includes detailed questionnaire, electronic health record and genetic data (8). Genotyping and sample quality control have been described previously (9). UK Biobank samples were genotyped using Affymetrix UK BiLEVE Axiom array and Affymetrix UK Biobank Axiom array and imputed to the combined 1000 Genomes Project v.3 and UK10K reference panels using SHAPEIT3 and IMPUTE3.18. Samples were included on the basis of female sex (genetic and self-reported). Duplicates, individuals with high degree of relatedness (>10 relatives), and one of each related pair of first-degree relatives were removed. Samples were also excluded using standard quality control criteria.

Genetically inferred ancestry information in UK Biobank was derived using iAdmix (10), which estimates maximum likelihood admixture proportions for an individual's ancestry using population allele frequencies from the 1000 Genomes Project. We estimated the ancestry proportions for four global populations: African, East Asian, European, South Asian. Individuals were classified as 'Mixed' if the proportion of their main ancestry did not reach 80% (70% for South Asian).

*KARMA*

The KARolinska Mammography Project for Risk Prediction of Breast Cancer (KARMA) cohort is a large prospective population-based cohort (11) of Swedish women (N=66,415), age range 40-74. For each participant, it contains data first-degree cancer family history (FH) and QRFs (self-reported), mammographic density (MD) as cBIRADS (12) and cancer diagnoses/deaths (from linkages to healthcare registers). KARMA was previously used for validating BOADICEA (13); here, we used it to assess the impact of using the UK Biobank QRFs distributions on the predicted risks.

*MyBrCa and SGBCC*

The proportions of the overall polygenic component explained by the PGS ('statistical methods' section) in women of East and South Asian ancestry were derived using data (14, 15) from two studies, from Malaysia (MyBrCa (16)) and Singapore (SGBCC (17)), participating in the Breast Cancer Association Consortium (BCAC).

For MyBrCa, cases were a mixture of prevalent and incident BCs, identified at the Breast Cancer Clinic in University Malaya Medical Centre (from January 2003 to July 2014) and Subang Jaya Medical Centre (from September 2012 to September 2014). Controls were cancer-free individuals (37-74 years) selected from women attending mammographic screening at the same hospitals. For

SGBCC, cases were a mixture of prevalent and incident BCs (in situ or invasive), diagnosed at National University Hospital between 2006 and 2013. Controls were Singaporeans or Singaporean Permanent Residents, 21 years and older; they were recruited between 2006 and 2010 through word-of-mouth, personal recommendations, 'cold-calling' and door-to-door invitations.

All samples were genotyped on the Oncoarray as described previously (18). Analyses were restricted to women with age of diagnosis or last observation less than 80 years and classified into either 'East Asian' or 'South Asian' according to genetic data. Women were from three main self-reported ethnic groups: Malay, Chinese and Indian. For analyses of East Asian women, samples from Chinese and Malay women were combined, and only women classified as 'East Asian' according to genetic data were included in the analyses. The South Asian group comprised women self-reporting as Indian that were also classified as 'South Asian' according to genetic data. **Table S15** reports studies, number of participants by ethnicity, and mean/SD age at diagnosis or interview (for cases and controls, respectively).

*Breast Cancer Surveillance Consortium*

To estimate the distributions of breast density (using BIRADS categories) for each ethnicity, and to calculate the corresponding odds ratios (ORs) for the association with BC risk, we used the public version of the risk estimation dataset from the Breast Cancer Surveillance Consortium (BCSC dataset, 2012 version) (19). It contains information on 1,007,657 unaffected women that had mammograms between 1996 and 2002. Each entry contains data on MD (as BIRADS category), age at screening (as 5-year aggregate), 'ethnicity' (Hispanic or not), 'race' (White, Asian, Black, Native American, Other/Mixed), number of first-degree relative affected with BC, and some QRFs (BMI, age at first birth, hormone replacement therapy (HRT) use). BCSC also has data on previous surgical procedures on breast or surgical menopause. For each woman, it is recorded whether she developed invasive BC within 1 year after the mammogram; due to data aggregation and anonymisation, the exact timing of the cancer diagnosis is unknown.

#### Cancer incidences and tumour subtype distributions

##### *Ethnicity-specific population cancer incidences*

For each cancer type (breast, ovarian, pancreatic, prostate), we considered that

$$I_{pop} = I_{asian} * P_{asian} + I_{black} * P_{black} + I_{mixed} * P_{mixed} + I_{white} * P_{white} + I_{other} * P_{other}$$

$$= I_{white} * (R_{asian} * P_{asian} + R_{black} * P_{black} + R_{mixed} * P_{mixed} + 1 * P_{white}) + I_{pop} * P_{other}$$

where $I_{pop}$ are the BOADICEA sex and cohort-specific population incidences and $I_{ethnicity}$ are the sex, cohort and ethnicity-specific incidences. $R_{ethnicity}$ are the published incidence rate ratios (20) ($R_{white} = 1$). Published incidence rate ratios were available for the <65 and the ≥65 years age groups. However, there was very little difference in the ratios between the two age groups for the different ethnicities, or there were very few cases in one of the two groups. Therefore, for simplicity, we assumed that the proportions were constant with age. $P_{ethnicity}$ are the ethnicity proportions obtained from the ONS (21). We used the ethnicity proportions from the 2016 census, matching the timespan (2013-2017) covered in the paper from Delon *et al.*, 2022 (20). $R_{ethnicity}$ and $P_{ethnicity}$ can be found in **Table S1**.

We defined

$$\theta = \frac{(1 - P_{other})}{(R_{asian} * P_{asian} + R_{black} * P_{black} + R_{mixed} * P_{mixed} + 1 * P_{white})}$$

thus obtaining the cancer incidences for each sex, cohort and ethnic group:

$$I_{ethnicity} = I_{pop} * R_{ethnicity} * \theta$$

*Gene- and ethnicity- specific tumour subtype distributions*

The age-specific subtype proportions for each ethnicity group were based on published estimates from NCRAS (National Cancer Registration and Analysis Service) data (22, 23). Those analyses involved fitting logistic regression models to predict the probability of the breast cancer (BC) subtype, after imputing missing data. Age at diagnosis was modelled using quadratic splines with knots placed at 35, 45 and 55 for the ER-positive subtype and 35 and 45 for the TN and ER-negative non-TN subtypes. Additionally, an interaction term between the age at BC diagnosis and ethnicity was included in each model (22, 23).

#### Genetic components

##### *Polygenic Scores*

*PGS parameters estimation for breast cancer*

The retrospective likelihood approach requires overall population age-specific incidence rates to be specified. In these analyses, population age-specific incidence rates by country (Singapore or Malaysia) and ethnicity (Malay, Chinese or Indian) were derived from the Singapore Cancer Registry (2013-2017) and the Penang Cancer Registry (2012-2016). Covariates included in these analyses were the country of origin of the study and 10 PCs (24). While the $\alpha_{RL}$ approach explicitly models the unmeasured polygenic component, another approach to estimating α makes the simplifying assumption that the polygenic standard deviation (SD) of the known polygenic component in BOADICEA can be approximated by the marginal age-specific log-odds ratio (log-OR) per unit SD of the PGS. The α parameter obtained with this method is denoted $\alpha_{GLM}$.

The mean and SD, the log-OR per unit SD of PGSs, and the parameters $\alpha_{GLM}$ and $\alpha_{RL}$ were estimated separately for East Asian and South Asian women.

For African women, there were insufficient data to estimate α directly. Therefore, α was estimated from published estimates of the OR per 1 SD, assuming that α is proportional to the OR. The ORs estimates from the three studies (25-27) were similar, with a combined estimate of 1.27 (95% CI: 1.23-1.31), based on a fixed-effect meta-analysis. Thus, using East Asians for comparisons, we calculated:

$$\alpha_{African} = \ln(1.27) * \alpha_{EastAsian} / \ln(OR)_{EastAsian}$$

East Asians were used as the reference because (a) the population is arguably more comparable to Africans than Europeans (lower risk, higher proportion of ER-negative etc.) and (b) this is a more conservative assumption. The populations differ with reference to allele frequencies and linkage disequilibrium; however, these affect the OR per 1 SD too and, in the absence of other evidence, the proportionality assumption seems reasonable.

*PGS parameters estimation for ovarian cancer*

For each SNP $i$, the ancestry-specific allele frequency $f_i$ was estimated using Plink2 from genotyped and imputed data. Assuming the log-odds ratio $\beta_i$ associated with each SNP remained the same across ancestries, the variance explained by the PGS was calculated using the formula (2):

$$\sigma_{PGS}^2 = \sum_{i=1}^{36} \ln\left(\frac{\left(1-f_i\right)^2 + 2\left(1-f_i\right)f_i e^{2\beta_i} + f_i^{\,2} e^{4\beta_i}}{\left(\left(1-f_i\right)^2 + 2\left(1-f_i\right)f_i e^{\beta_i} + f_i^{\,2} e^{2\beta_i}\right)^2}\right)$$

The square root of the proportion of the overall polygenic variance explained by the PGS, namely $\alpha_i$ for each ancestry, was calculated as:

$$\alpha = \frac{\sigma_{PGS}}{1.4156}$$

where 1.41562 is the EOC polygenic SD assumed in the model (2).

*PGS for individuals of mixed genetic ancestry*

Normalised polygenic scores are employed in BOADICEA; therefore, they depend on ancestry even though the raw PGS is the same. For ancestry $i$, we have:

$$R_{pgs} = \sum_{k=1}^{K} \beta_k g_k\,, \qquad Z_i = \frac{\left(R_{prs} - \mu_i\right)}{\sigma_i}$$

where

$$\mu_i = E\left(R_{prs} \middle| pop\ i\right) = 2\sum_{k=1}^{K} \beta_k \rho_{ik}\,, \qquad \sigma_i^2 = var(pop\ i) = 2\sum_{k=1}^{K} \rho_{ik}(1-\rho_{ik})\beta_k^2$$

$\rho_{ik}$ being the ancestry-specific frequency of locus $k$, $\beta_k$ being the (fixed) per-allele log-OR.

For 'Mixed' individuals, PGS must be normalised taking into account the proportion of each ancestry; the overall *alpha* ($\alpha$) must be similarly transformed. Given $P_i$ proportion, $\gamma_i$ effect size and $Z_i$ normalised PGS for ancestry $i$, we can define

$$S = \sum_{i=1}^{M} P_i \gamma_i Z_i$$

so that

$$E(S) = 0, \qquad var(S) = \left(\sum_{i=}^{M} \frac{P_i \gamma_i}{\sigma_i}\right)^2 var\left(R_{prs}\right) \equiv \left(\sum_{i=}^{M} \frac{P_i \gamma_i}{\sigma_i}\right)^2 \sigma^2$$

and therefore

$$Z = \frac{S - E(S)}{sd(S)} = \frac{\sum_{i=1}^{M} P_i \gamma_i Z_i}{\sigma \sum_{i=}^{M} \frac{P_i \gamma_i}{\sigma_i}}, \qquad \alpha = \frac{\sum_{i=1}^{M} P_i \alpha_i \sigma_i}{\sigma}$$

Clearly, $\sigma$ depends on the proportion of each ancestry, but it also considers that different loci could belong to different ancestries. Indeed we have

$$\sigma^2 = var(x_{prs}) = E(var(pop\ i)) + var(E(pop\ i)) = E(\sigma_i^2) + var(\mu_i)$$
$$= \sum_{i=1}^{M} P_i \sigma_i^2 + 4 \sum_{k=1}^{K} w_k^2 \left( \sum_{i=1}^{M} P_i {\rho_{ik}}^2 - \left( \sum_{i=1}^{M} P_i \rho_{ik} \right)^2 \right)$$

The full formula derivation can be found at the end of these Supplementary Materials.

##### *Rare genetic variants*

*Literature search on risks associated with rare genetic variants*

For the non-European populations, using the searching keywords 'breast cancer', 'germline pathogenic variants' and 'Asian' returned 75 articles. Using the searching keywords 'breast cancer', 'germline pathogenic variants' and 'African' returned 35 articles. Using the searching keywords 'ovarian cancer', 'germline pathogenic variants' and 'Asian' returned 30 articles. Using the search keywords 'ovarian cancer', 'germline pathogenic variants' and 'African' returned 9 articles.

#### Mammographic Density

In this analysis, we employed the public version of the BCSC dataset (2012 version), discarding data on women that had previous surgical procedures, obtaining a dataset of 718,889 entries. As ethnicities, we considered all combinations of the 'race' and 'ethnicity' categories in the dataset; however, we merged the 'Other' and 'Native American' categories, and we did not consider Hispanic women in the downstream analysis. **Table S2** shows the number of unaffected and affected women by ethnicity.

We employed Multivariate Imputation by Chained Equations (MICE, (28)) to impute missing values in the following categories: 'ethnicity' (Hispanic or not), body mass index (BMI), number of affected (1$^{st}$ degree) relatives, age at first live birth (for parous women), age at menopause and use of hormone replacement therapy (HRT) if applicable. Ten imputed datasets were generated through 20 iterations. **Table S3** shows the (average) number of unaffected and affected women by ethnicity after imputation.

We calculated ORs using logistic regression on the imputed datasets (*survival* R package, (29)), adjusting for all BOADICEA QRFs contained in the dataset and stratifying by age group (before age 50y, after age 50y). Two different methods were used for calculating ethnicity-specific ORs: analysing the entire dataset and fitting an interaction term between MD and ethnicity; and analysing each ethnic group separately. The ORs calculated from the imputed datasets were combined using the Rubin's rule; $D_2$ pooling method was applied to pool the statistics from likelihood-ratio tests.

In this analysis, Hispanics were considered as separate ethnic groups (White Hispanics, Black Hispanics etc.). However, since this ethnic group not considered in this work, their respective ORs were not reported in this manuscript, and the distribution of BIRADS categories was calculated for non-Hispanic women only.

### Results

#### Cancer incidences and tumour subtype distributions

##### *Gene- and ethnicity- specific tumour subtype distributions*

In White women, the proportion of ER-positive tumours increases with age and then plateaus at approximately age 50 years. The proportion of ER-positive tumours in East- and South-Asian women also increase with age but are lower than for Whites. In Black women, the proportions of ER-positive disease are lower and varies little with age. Conversely, Black women are more likely to be diagnosed with triple negative (TN) disease, with the proportion varying little with age. In White and Asian women, the proportion of TN cases decreases with age, but is higher in Asian women than in White women.

Age-, gene- and subtype specific proportions were calculated for women of White, Black, East Asian, South Asian, and Mixed ethnicity. Proportions in the 'Population' were based on 1) non-carrier proportions published in JAMA Oncology (30) and 2) general population published estimates from NCRAS data (22, 23). Estimates for White non-carrier women in BCAC dataset (from BRIDGES) (30) are also shown for comparison with NCRAS data (smoothed using *lowess* and quadratic splines, respectively).

#### Genetic components

##### *Rare genetic variants*

Five large studies have examined the associations between rare PVs and BC risk in individuals from different ethnicities/ancestries. The BRIDGES study (Breast Cancer Risk after Diagnostic Gene Sequencing) analysed a 34-gene panel using data from the Breast Cancer Association Consortium (BCAC).The study included 42,060 breast cancer cases of European ancestry and 6,279 cases of Asian ancestry, along with 44,035 cancer-free controls of European ancestry and 6,047 cancer-free controls of Asian ancestry (31). The CARRIERS sequenced the same gene panel in 32,247 BC cases and 32,544 controls. The study population included 1,282 BC cases and 1,269 controls of Asian ancestry, 3,946 cases and 4,954 controls of African American ancestry, and 25,287 cases and 24,770 controls of non-Hispanic White ancestry (32). The Ghana Breast Health Study (GBHS) sequenced the same gene panel in 871 cases and 1563 controls of African ancestry (33). The African American study sequenced 37-gene panel in 5054 cases and 4993 controls of African American ancestry (34). The Japanese study sequenced 11 BC genes in 7051 BC cases and 11241 controls of Japanese ancestry (35). An additional study estimated BC and EOC relative risks associated with *BRCA1* and *BRCA2* in 572 families from Malaysia and Singapore (36).

To estimate the overall *CHEK2* PV frequency in non-Europeans, we used the frequency of other (non c.1100del) PVs in controls in the BRIDGES dataset, combined with the c.1100delC frequency in East Asians. We adjusted for an assumed sensitivity of 0.957 and a proportion (0.16) of large rearrangements that would have been being undetectable (1). We assumed this frequency applied also to the other non-European ancestries.

#### Mammographic Density

The previous implementation of BOADICEA (BC model v6) assumed that QRFs distributions do not vary with age; for density it uses distributions for White women younger than 50 years, published in Tice *et al.*, 2008 (37). For consistency, here we implemented the distributions from the <50 years age group for all ethnicities. This coarse simplification does not consider the relevant change in MD with age; a concurrent model extension, based on continuous MD measures, incorporates age-specific MD density distributions into BOADICEA (38) for all ethnicities.

#### Multifactorial models

**Table S22** summarises the ethnicity and ancestry-specific data used for each model component. If known, genetic ancestry is employed for specifying the PGS model; *CHEK2 PV* frequency is always assumed from the self-reported ethnicity to allow for the fact that in practice not everyone with gene-panel sequencing will have genetic ancestry determined. When ethnicity is unknown, the default, population-wide parameters are employed. When the self-reported ethnicity is 'Other', the model assumes the default parameters apart from the *CHEK2* PV frequency, where the frequency for non-Europeans is used.

### Figure and table legends

**Figure S1.** Ethnicity-specific cancer incidences per 100,000 people in the UK, employed in the BC and EOC models (1980-1989 birth cohort). Panel A, breast cancer (BC); panel B, ovarian cancer (OC); panel C, pancreatic cancer (PanC); panel D, prostate cancer (ProC).

**Figure S2.** *[in separate file]* ER-positive predicted proportions by age at cancer diagnosis and ethnicity, based on published estimates from NCRAS data. Age at diagnosis was modelled using quadratic splines with knots placed at 35, 45 and 55 years.

**Figure S3.** *[in separate file]* ER-negative non-TN predicted proportions by age at cancer diagnosis and ethnicity, based on published estimates from NCRAS data. Age at diagnosis was modelled using quadratic splines with knots placed at 35 and 45 years.

**Figure S4.** *[in separate file]* TN predicted proportions by age at cancer diagnosis and ethnicity, based on published estimates from NCRAS data. Age at diagnosis was modelled using quadratic splines with knots placed at 35 and 45 years.

**Figure S5.** *[in separate file]* ER-positive predicted proportions by age at cancer diagnosis and ethnicity, in PV carriers and in the general population.

**Figure S6.** *[in separate file]* ER-negative non-TN predicted proportions by age at cancer diagnosis and ethnicity, in PV carriers and in the general population.

**Figure S7.** *[in separate file]* TN predicted proportions by age at cancer diagnosis and ethnicity, in PV carriers and in the general population.

**Figure S8.** Standard deviations (SDs) for multiple (random) combinations of ancestry proportions, calculated using the full formula detailed in Methods and by weighted averaging the ancestry-specific SDs. Each random combination is created by successive extraction of ancestry proportions from continuous uniform distributions: the first proportion (P1) is extracted from U(0,1); the second one (P2) from U(0,1-P1); the third one (P3) from U(0,1-P1-P2); the fourth proportion (P4) is set to 1 - P1 - P2 - P3.

**Figure S9.** Predicted breast cancer risks by age for a female born in 2000 (Mixed ethnicity, unknown family history and QRFs), based on ancestry and PGS. Ancestry-specific standardised PGS were calculated from the same raw PGS values, corresponding to 10th, 25th, 45th, 55th, 75th and 90th percentile of the standardised PGS distribution in Europeans. 'Mixed (simple)' was obtained with a simplified formula for calculating σ ($\boldsymbol{\sigma^2 = \sum_{i=1}^{M} P_i \sigma_i^2}$); 'Mixed (full)' was obtained with the complete formula for σ.

**Figure S10.** Comparison of 10-year breast cancer (BC) risk predictions in the KARMA cohort. X-axis shows the BC risks predicted by using the default QRFs distributions in BOADICEA (36) whereas y-axis shows the BC risks predicted by switching to use QRFs distributions estimated in the UK Biobank White populations. The dashed line is the diagonal line with slope equal to 1 (corresponding to no difference in the risk predictions).

**Figure S11.** Predicted epithelial ovarian cancer risks by age for a female born in 1985 (unknown family history and QRFs) based on ethnicity and pathogenic variant carrier status (for the major genes considered in the EOC model).

**Figure S12.** Predicted breast cancer risks by age for a female born in 1985 (unknown family history and QRFs) based on ethnicity and pathogenic variant carrier status (for the major genes considered in the BC model).

**Figure S13.** Predicted lifetime (age 20-80 years) BC risks for women of different ethnicities, untested for PVs and unknown family history. Risks are based on different combinations of risk predictors, each drawn in a separate panel: (A,B) PGS only; (C,D) QRFs only; (E,F) PGS and QRFs; (G,H) PGS and QRFs and MD. (A,C,E,G) Probability density function against absolute risk; (B,D,F,H) absolute risk against cumulative distribution. BC, breast cancer; PGS, polygenic score; QRFs, questionnaire-based risk factors; MD, mammographic density. The backgrounds of the graphs are shaded to indicate three lifetime BC risk categories: less than 17% (light yellow); between 17% and 30% (yellow); above 30% (light blue).

**Figure S14.** Predicted 10-year (to age 50 years) BC risks for women of different ethnicities, untested for PVs and unknown family history. Risks are based on different combinations of risk predictors, each drawn in a separate panel: (A,B) PGS only; (C,D) QRFs only; (E,F) PGS and QRFs; (G,H) PGS and QRFs and MD. (A,C,E,G) Probability density function against absolute risk; (B,D,F,H) absolute risk against cumulative distribution. BC, breast cancer; PGS, polygenic score; QRFs, questionnaire-based risk factors; MD, mammographic density. The backgrounds of the graphs are shaded to indicate three 10-year BC risk categories: less than 3% (light yellow); between 3% and 8% (yellow); above 8% (light blue).

**Figure S15.** Predicted lifetime (age 20-80 years) EOC risks for women of different ethnicities, untested for PVs and unknown family history. Risks are based on different combinations of risk predictors, each drawn in a separate panel: (A,B) PGS only; (C,D) QRFs only; (E,F) PGS and QRFs. (A,C,E) Probability density function against absolute risk; (B,D,F) absolute risk against cumulative distribution. EOC, epithelial ovarian cancer; PGS, polygenic score; QRFs, questionnaire-based risk factors. The backgrounds of the graphs are shaded to indicate three lifetime EOC risk categories: less than 2.5% (light yellow); between 2.5% and 5% (yellow); above 5% (light blue).

**Figure S16.** Predicted 10-year (to age 50 years) EOC risks for women of different ethnicities, untested for PVs and unknown family history. Risks are based on different combinations of risk predictors, each drawn in a separate panel: (A,B) PGS only; (C,D) QRFs only; (E,F) PGS and QRFs. (A,C,E) Probability density function against absolute risk; (B,D,F) absolute risk against cumulative distribution. EOC, epithelial ovarian cancer; PGS, polygenic score; QRFs, questionnaire-based risk factors.

**Figure S17.** Predicted lifetime (age 20-80 years) BC risks for women of different ethnicities, untested for PVs and unknown family history, based on different combinations of risk predictors. Distributions for each ethnicity are drawn in separate panels: (A,B) Population/Unspecified; (C,D) White; (E,F) Black; (G,H) East Asian; (I,J) South Asian; (K,L) Mixed. The straight lines (labelled 'ethnicity only') are equivalent to the "population" risk of BC for the specific ethnicity plotted. (A,C,E,G,I,K) Probability density function against absolute risk; (B,D,F,H,J,L) absolute risk against cumulative distribution. BC, breast cancer; PGS, Polygenic Score; QRFs, questionnaire-based risk factors; MD, mammographic density. The backgrounds of the graphs are shaded to indicate three lifetime BC risk categories: less than 17% (light yellow); between 17% and 30% (yellow); above 30% (light blue).

**Figure S18.** Predicted 10-year (to age 50 years) BC risks for women of different ethnicities, untested for PVs and unknown family history, based on different combinations of risk predictors. Distributions for each ethnicity are drawn in separate panels: (A,B) Population/Unspecified; (C,D) White; (E,F) Black; (G,H) East Asian; (I,J) South Asian; (K,L) Mixed. The straight lines (labelled 'ethnicity only') are equivalent to the "population" risk of BC for the specific plotted. (A,C,E,G,I,K) Probability density function against absolute risk; (B,D,F,H,J,L) absolute risk against cumulative distribution. BC, breast cancer; PGS, polygenic score; QRFs, questionnaire-based risk factors; MD, mammographic density. The backgrounds of the graphs are shaded to indicate three 10-year BC risk categories: less than 3% (light yellow); between 3% and 8% (yellow); above 8% (light blue).

**Figure S19.** Predicted lifetime (age 20-80 years) EOC risks for women of different ethnicities, untested for PVs and unknown family history, based on different combinations of risk predictors. Distributions for each ethnicity are drawn in separate panels: (A,B) Population/Unspecified; (C,D) White; (E,F) Black; (G,H) East Asian; (I,J) South Asian; (K,L) Mixed. The straight lines (labelled 'ethnicity only') are equivalent to the "population" risk of EOC for the specific ethnicity plotted. (A,C,E,G,I,K) Probability density function against absolute risk; (B,D,F,H,J,L) absolute risk against cumulative distribution. EOC, epithelial ovarian cancer; PGS, polygenic score; QRFs, questionnaire-based risk factors. The backgrounds of the graphs are shaded to indicate three lifetime EOC risk categories: less than 2.5% (light yellow); between 2.5% and 5% (yellow); above 5% (light blue).

**Figure S20.** Predicted 10-year (to age 50 years) EOC risks for women of different ethnicities, untested for PVs and unknown family history, based on different combinations of risk predictors. Distributions for each ethnicity are drawn in separate panels: (A,B) Population/Unspecified; (C,D) White; (E,F) Black; (G,H) East Asian; (I,J) South Asian; (K,L) Mixed. The straight lines (labelled 'ethnicity only') are equivalent to the "population" risk of EOC for the specific ethnicity plotted. (A,C,E,G,I,K) Probability density function against absolute risk; (B,D,F,H,J,L) absolute risk against cumulative distribution. EOC, epithelial ovarian cancer; PGS, polygenic score; QRFs, questionnaire-based risk factors.

**Figure S21.** Predicted BC and EOC lifetime (age 20-80 years) risks for women of different ethnicities, BRCA2 PV carrier and unknown family history; risks calculated using all risk predictors: PGS, QRFs, and MD. (A,B) BC risk; backgrounds are shaded to indicate three risk categories: <17% (light yellow); 17% - 30% (yellow); ≥ 30% (light blue). (C,D) EOC risk; backgrounds are shaded to indicate three risk categories: < 2.5% (light yellow); 2.5% - 5% (yellow); ≥ 5% (light blue). (A,C) probability density function against absolute risk; (B,D) absolute risk against cumulative distribution. BC, breast cancer; EOC, epithelial ovarian cancer; PGS, polygenic score; QRFs, questionnaire-based risk factors; MD, mammographic density.

**Table S1**. Ethnicity proportions ($P_{ethnicity}$) obtained from the ONS (19) and incidence rate ratios by ethnic group and cancer type ($R_{ethnicity}$), relative to White (18). East and South Asian ethnicities were grouped in a single group due to small numbers.

**Table S2**. Summary of the BCSC dataset used: distribution of women across all 'races' and 'ethnicities', by age group (split age: 50 years) and by diagnosis within 1 year from the mammogram screening.

**Table S3**. Summary of the BCSC dataset used, after imputation: distribution of women across all 'races' and 'ethnicities', by age group (split age: 50 years) and by diagnosis within 1 year from the mammogram screening.

**Table S4**. *[in separate file]* ER-positive predicted proportions by age at cancer diagnosis and ethnicity, in PV carriers and in the general population.

**Table S5.** *[in separate file]* ER-negative non-TN predicted proportions by age at cancer diagnosis and ethnicity, in PV carriers and in the general population.

**Table S6.** *[in separate file]* TN predicted proportions by age at cancer diagnosis and ethnicity, in PV carriers and in the general population.

**Table S7.** *[in separate file]* SNPs and weights included in the construction of the PGS models. There are three surrogates correlated with original SNP from Mavaddat et al. 2019 at r2>0.9). The 307-SNP PGS does not include 22_29203724_C_T and 22_29551872_A_G; the 303-SNP PGS also excludes in addition SNPs 6_27425644_G_C, 15_46680811_C_A, 3_29294845_C_T, and 11_46318032_C_G. [a]SNP variant name; [b]Position based on build 37.

**Table S8.** *[in separate file]* Allele frequencies (EAF)s by genetic ancestry for SNPs included in the PGS models. EAFs were estimated in UK Biobank individuals without a diagnosis of cancer or mastectomy prior to the start of follow-up. Genetic ancestry groupings are according to iadmix, derived as described in the Methods: EUR, European; AFR, African; EAS, East Asian; SAS, South Asian. [a]SNP variant name.

**Table S9**. *[in separate file]* Mean and standard deviation (SD) of the 309-SNP BC PGS in UK Biobank individuals without a diagnosis of cancer or mastectomy prior to the start of follow-up. Genetic ancestry groupings are according to iadmix, derived as described in the Methods; self-reported ethnicity groupings are as described in the Methods.

**Table S10**. *[in separate file]* Mean and standard deviation (SD) of the 307-SNP BC PGS in UK Biobank individuals without a diagnosis of cancer or mastectomy prior to the start of follow-up. Genetic ancestry groupings are according to iadmix, derived as described in the Methods; self-reported ethnicity groupings are as described in the Methods. The 307-SNP PGS does not include 22_29203724_C_T and 22_29551872_A_G.

**Table S11**. *[in separate file]* Self-reported ethnicity vs genetic ancestry (iadmix) in UK Biobank for individuals without a diagnosis of cancer. Genetic ancestry groupings are according to iadmix, derived as described in the Methods: EUR, European; AFR, African; EAS, East Asian; SAS, South Asian. Self-reported ethnicity groupings are as described in the Methods. In total, 226340 women, 200073 men and 426413 men and women were included in each analysis.

**Table S12.** *[in separate file]* SNPs, weights and allele frequencies (EAFs) by genetic ancestry included in the construction of the PGS model. EAFs were estimated in UK Biobank individuals without a diagnosis of cancer prior to the start of follow-up. SNPs log-OR were derived from OCAC published data. Genetic ancestry groupings are according to iadmix, derived as described in the Methods. [a]SNP variant name; [b]Position based on build 37.

**Table S13**. *[in separate file]* Mean, standard deviation (SD) and *alpha* parameters of the 36-SNP OC PGS. Mean and SD estimated in UK Biobank individuals without a diagnosis of cancer prior to the start of follow-up. α parameter estimated using SNPs frequencies from UK Biobank and SNPs log-OR from OCAC published data. Genetic ancestry groupings are according to iadmix, derived as described in the Methods.

**Table S14**. *[in separate file]* Mean, standard deviation (SD) and *alpha* parameters for BC PGS models. For Asians, parameters estimated using data from the MyBrCa and SGBCC studies. For Europeans, parameters based on the prospective and validation sets from Mavaddat *et al.*, 2019 , and the published procedure from Mavaddat *et al.*, 2023. [a]Number of SNPs included in PGS models: all 313 SNPs from Mavaddat *et al.*, 2019 (313-SNP PGS), UKB SNPs (309-SNP PGS), UKB SNPs excluding two CHEK2 SNPs (307-SNP PGS), no CHEK2 variants and imputation accuracy r2>0.5 (303-SNP PGS). [b]αGLM is based on logistic regression adjusted for country in which studies were conducted and principal components. [c]αRL is based on retrospective likelihood method as outlined in the Methods. [d]Results based on 6394 controls and 6150 cases among Chinese and Malay women classified as 'East-Asian' according to genetic data, with known age and age less than 80. [e]Results based on 1000 controls and 568 cases among Indian women classified as 'South-Asian' according to genetic data, with known age and age less than 80. [f]Results based on 22767 controls and 16151 cases; European women with known age and age less than 80.

**Table S15**. *[in separate file]* Statistics on the MyBrCa and SGBCC studies; for cases and controls, number of individuals and mean/standard deviation (SD) age at diagnosis/interview (years).

**Table S16**. Summary of published associations between breast cancer and rare pathogenic variants by different ancestries. Odds ratios [95% confidence intervals] were reported.

**Table S17**. Summary of published pathogenic variant frequencies (%) in the population by different ancestries without large rearrangement sensitivity adjustment.

**Table S18**. Estimated QRFs distributions in UK Biobank and MD distribution from BCSC. QRFs: questionnaire-based risk factors; MD: mammographic density; HRT: hormone replacement therapy; OC: oral contraceptive.

**Table S19**. Odds ratios [95% confidence intervals] for different BIRADS categories, estimated for each ethnicity separately. BIRADS B is the reference category for individuals of age less than 50 years (upper panel); BIRADS A is the reference category for individuals of age 50 years and over (lower panel).

**Table S20**. Ethnicity-specific Odds ratio estimates [95% confidence intervals], using entire BCSC population and considering an interaction term between MD and ethnicity. BIRADS B is the reference category for individuals of age less than 50 years (upper panel); BIRADS A is the reference category for individuals of age 50 years and over (lower panel).

**Table S21**. Proportion [95% confidence intervals] of BIRADS categories in different ethnic groups, for individuals of age less than 50 years (upper panel) and for individuals of age 50 years and over (lower panel).

**Table S22**. Source of the model parameters for each ethnic/ancestry group (as described in this manuscript). 'default' indicates parameters in previous BOADICEA models (BC v6, EOC v2).

**Table S23**. Proportion of women falling in different risk categories depending on their lifetime cancer risk (age 20 to 80 years) and their ethnicity, using different risk predictors. Unknown genetic background, mother with BC/EOC at 50y. First panel: lifetime BC risk; the thresholds between risk categories are 17%, between 'population' and 'moderate', and 30%, between 'moderate' and 'high' (NICE guidelines). Second panel: lifetime EOC risk, with thresholds at 3.5% and 5% (threshold for risk-reducing surgery, NICE guidelines).

**Table S24**. Proportion of women falling in different risk categories depending on their 10-year BC risk (age 40 to 50 years) and their ethnicity, using different risk predictors. Unknown genetic background, unknown FH. The thresholds between risk categories are 3%, between 'population' and 'moderate', and 8%, between 'moderate' and 'high' (NICE guidelines).

**Table S25**. Proportion of women falling in different risk categories depending on their 10-year BC risk (age 40 to 50 years) and their ethnicity, using different risk predictors. Unknown genetic background, mother with BC at 50y. The thresholds between risk categories are 3%, between 'population' and 'moderate', and 8%, between 'moderate' and 'high' (NICE guidelines).

### Figures

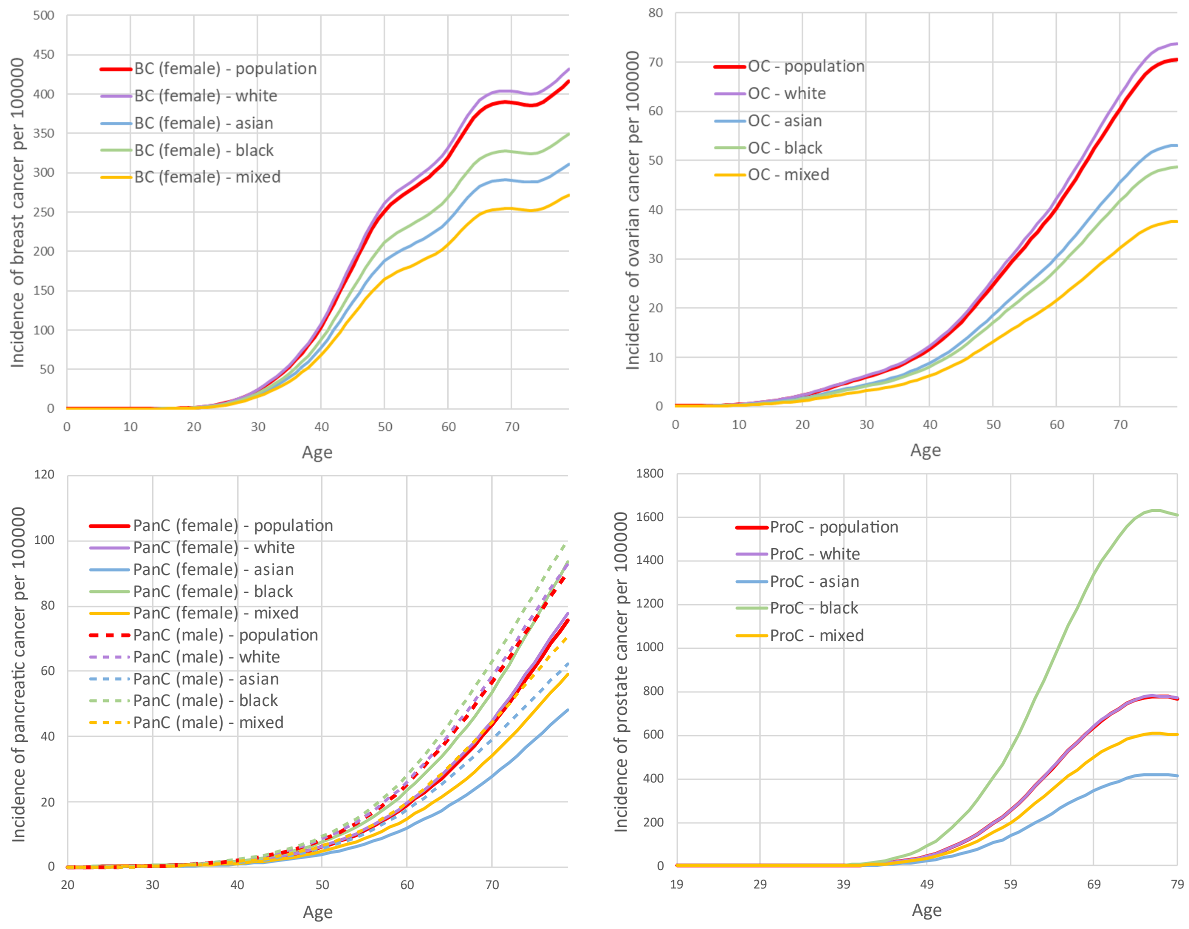

**Figure S1.** Ethnicity-specific cancer incidences per 100,000 people in the UK, employed in the BC and EOC models (1980-1989 birth cohort). Panel A, breast cancer (BC); panel B, ovarian cancer (OC); panel C, pancreatic cancer (PanC); panel D, prostate cancer (ProC).

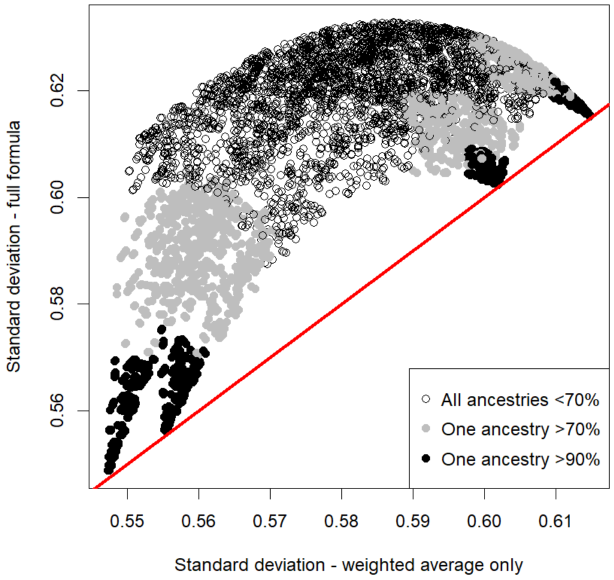

**Figure S8.** Standard deviations (SDs) for multiple (random) combinations of ancestry proportions, calculated using the full formula detailed in Methods and by weighted averaging the ancestry-specific SDs. Each random combination is created by successive extraction of ancestry proportions from continuous uniform distributions: the first proportion (P1) is extracted from $U(0,1)$; the second one (P2) from $U(0,1\text{-}P1)$; the third one (P3) from $U(0,1\text{-}P1\text{-}P2)$; the fourth proportion (P4) is set to 1 - P1 - P2 - P3.

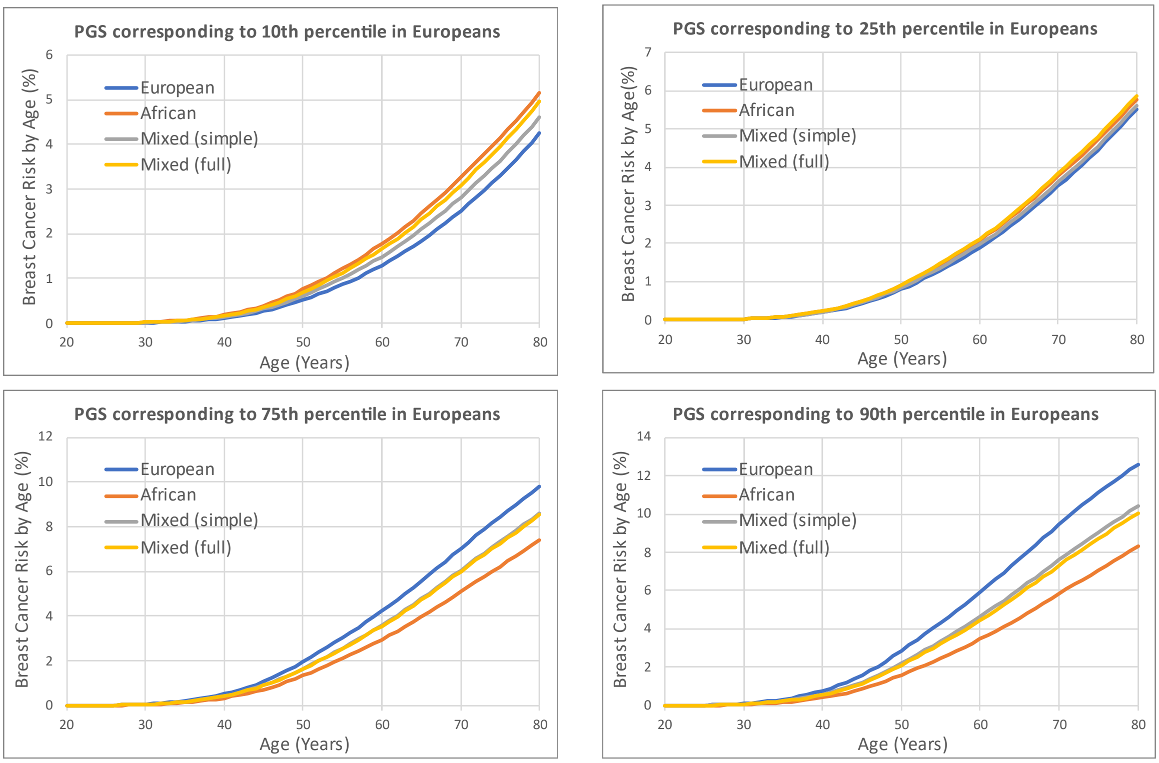

**Figure S9.** Predicted breast cancer risks by age for a female born in 2000 (Mixed ethnicity, unknown family history and QRFs), based on ancestry and PGS. Ancestry-specific standardised PGS were calculated from the same raw PGS values, corresponding to 10th, 25th, 45th, 55th, 75th and 90th percentile of the standardised PGS distribution in Europeans. ‘Mixed (simple)’ was obtained with a simplified formula for calculating σ ($\boldsymbol{\sigma^2} = \sum_{i=1}^{M} \boldsymbol{P_i \sigma_i^2}$); ‘Mixed (full)’ was obtained with the complete formula for σ.

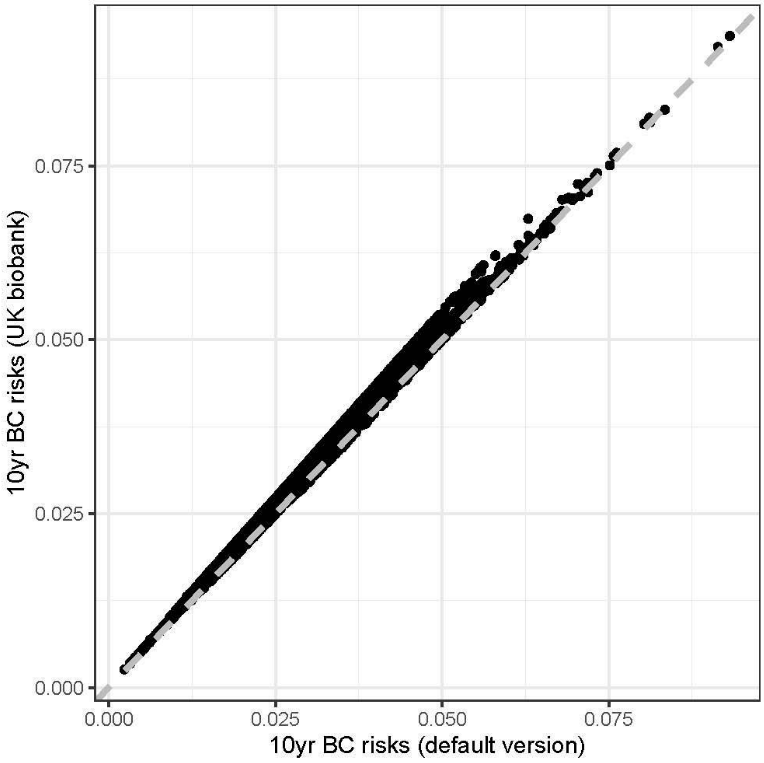

**Figure S10.** Comparison of 10-year breast cancer (BC) risk predictions in the KARMA cohort. X-axis shows the BC risks predicted by using the default QRFs distributions in BOADICEA (1) whereas y-axis shows the BC risks predicted by switching to use QRFs distributions estimated in the UK Biobank White populations. The dashed line is the diagonal line with slope equal to 1 (corresponding to no difference in the risk predictions).

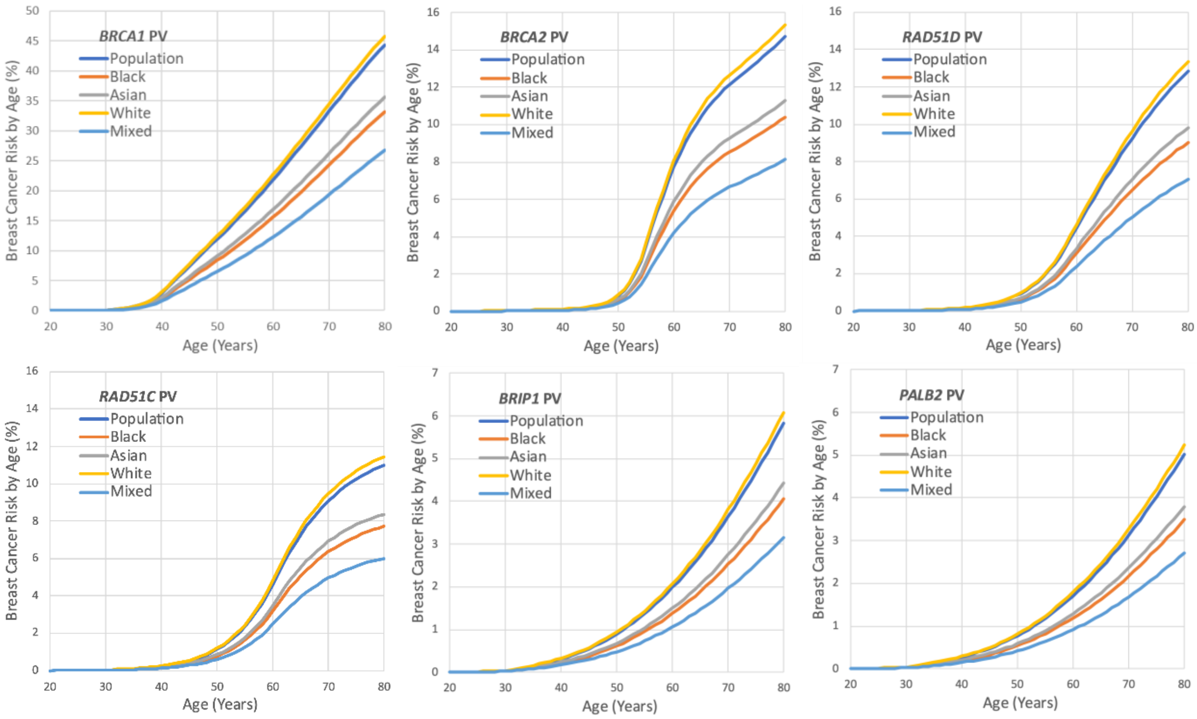

**Figure S11.** Predicted epithelial ovarian cancer risks by age for a female born in 1985 (unknown family history and QRFs) based on ethnicity and pathogenic variant carrier status (for the major genes considered in the EOC model).

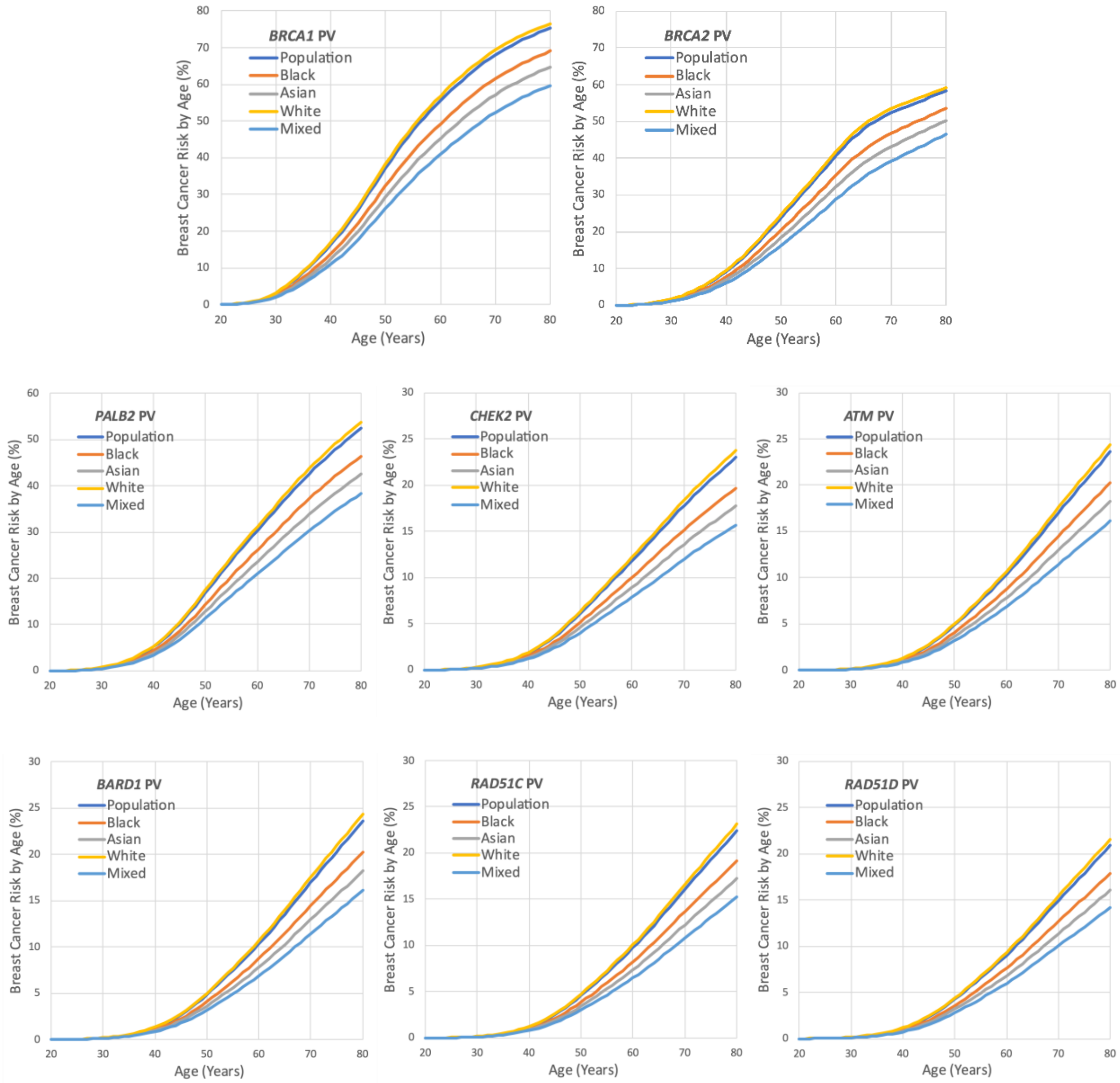

**Figure S12.** Predicted breast cancer risks by age for a female born in 1985 (unknown family history and QRFs) based on ethnicity and pathogenic variant carrier status (for the major genes considered in the BC model).

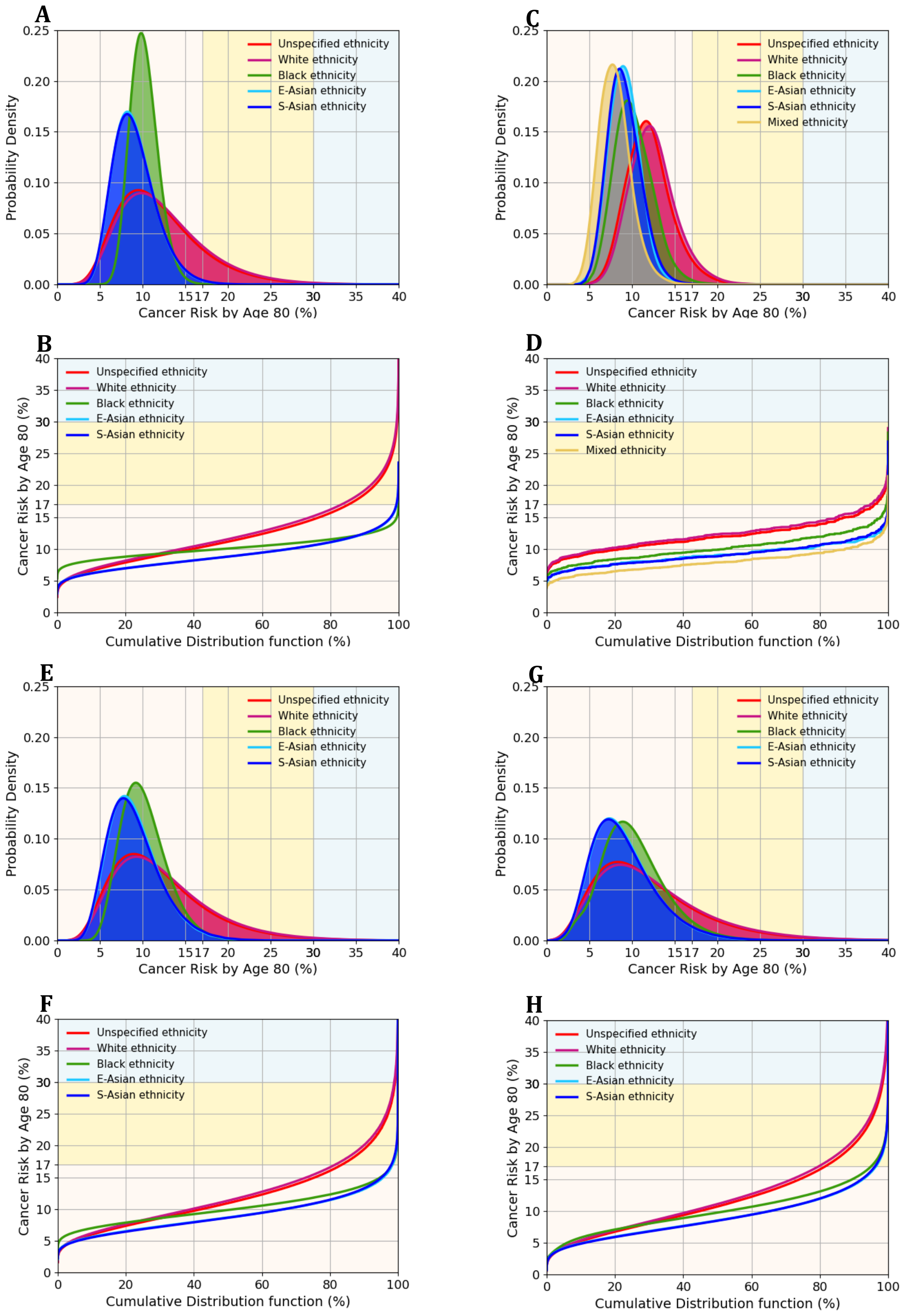

**Figure S13**. Predicted lifetime (age 20-80 years) BC risks for women of different ethnicities, untested for PVs and unknown family history. Risks are based on different combinations of risk predictors, each drawn in a separate panel: (A,B) PGS only; (C,D) QRFs only; (E,F) PGS and QRFs; (G,H) PGS and QRFs and MD. (A,C,E,G) Probability density function against absolute risk; (B,D,F,H) absolute risk against cumulative distribution. BC, breast cancer; PGS, polygenic score; QRFs, questionnaire-based risk factors; MD, mammographic density. The backgrounds of the graphs are shaded to indicate three lifetime BC risk categories: less than 17% (light yellow); between 17% and 30% (yellow); above 30% (light blue).

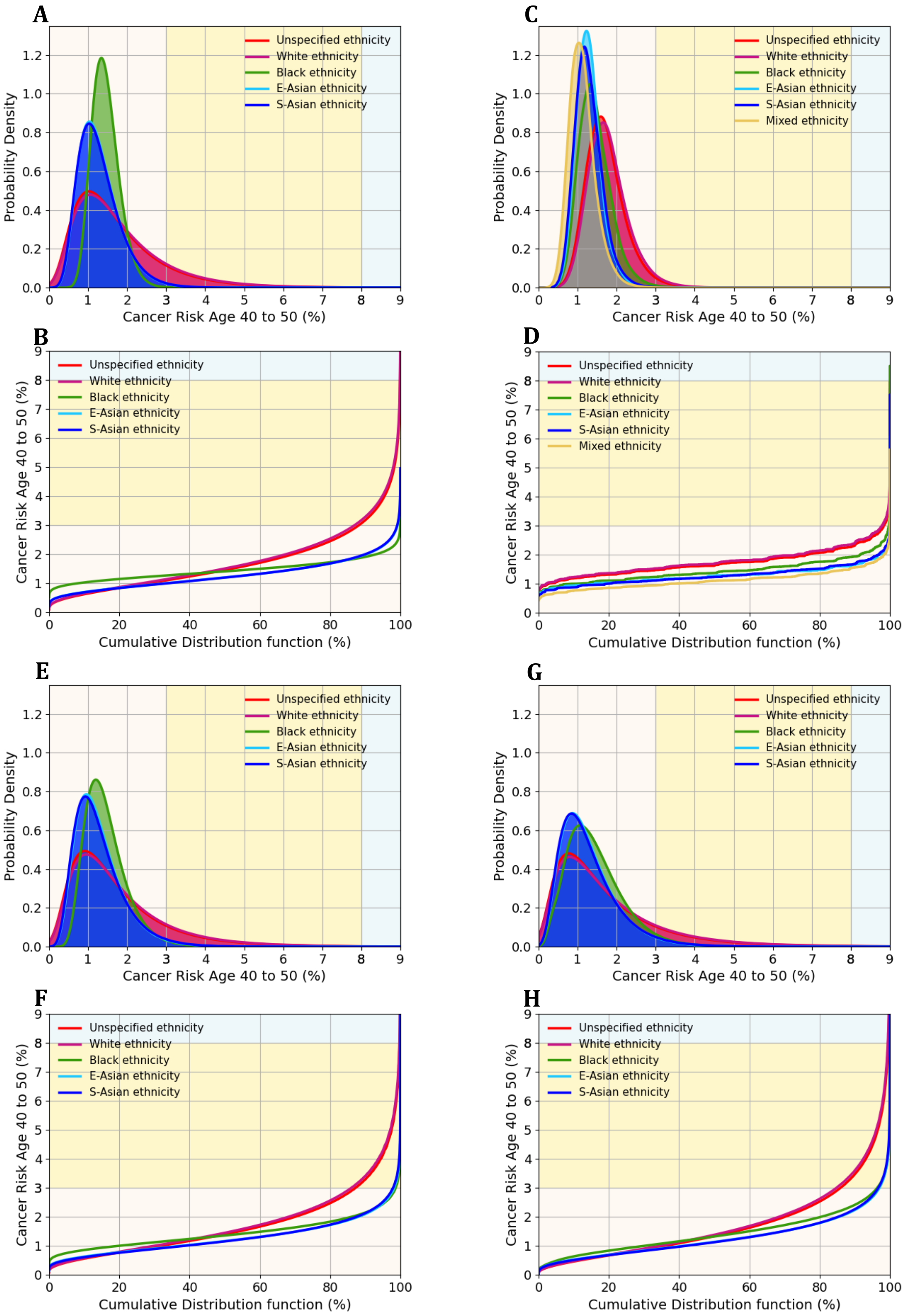

**Figure S14.** Predicted 10-year (to age 50 years) BC risks for women of different ethnicities, untested for PVs and unknown family history. Risks are based on different combinations of risk predictors, each drawn in a separate panel: (A,B) PGS only; (C,D) QRFs only; (E,F) PGS and QRFs; (G,H) PGS and QRFs and MD. (A,C,E,G) Probability density function against absolute risk; (B,D,F,H) absolute risk against cumulative distribution. BC, breast cancer; PGS, polygenic score; QRFs, questionnaire-based risk factors; MD, mammographic density. The backgrounds of the graphs are shaded to indicate three 10-year BC risk categories: less than 3% (light yellow); between 3% and 8% (yellow); above 8% (light blue).

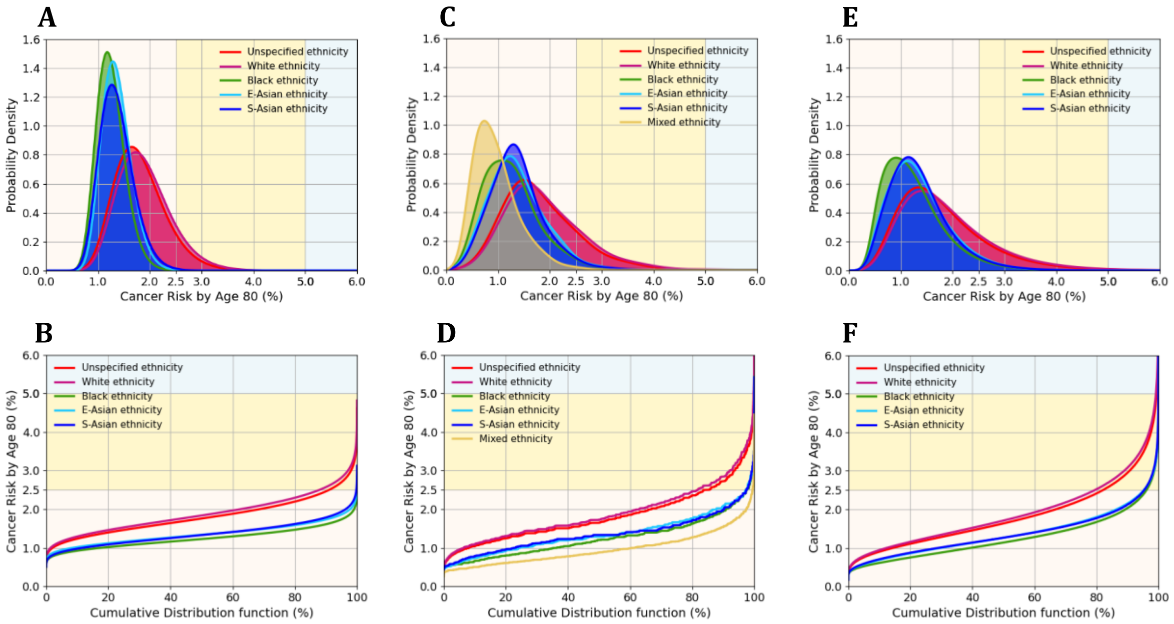

**Figure S15.** Predicted lifetime (age 20-80 years) EOC risks for women of different ethnicities, untested for PVs and unknown family history. Risks are based on different combinations of risk predictors, each drawn in a separate panel: (A,B) PGS only; (C,D) QRFs only; (E,F) PGS and QRFs. (A,C,E) Probability density function against absolute risk; (B,D,F) absolute risk against cumulative distribution. EOC, epithelial ovarian cancer; PGS, polygenic score; QRFs, questionnaire-based risk factors. The backgrounds of the graphs are shaded to indicate three lifetime EOC risk categories: less than 2.5% (light yellow); between 2.5% and 5% (yellow); above 5% (light blue).

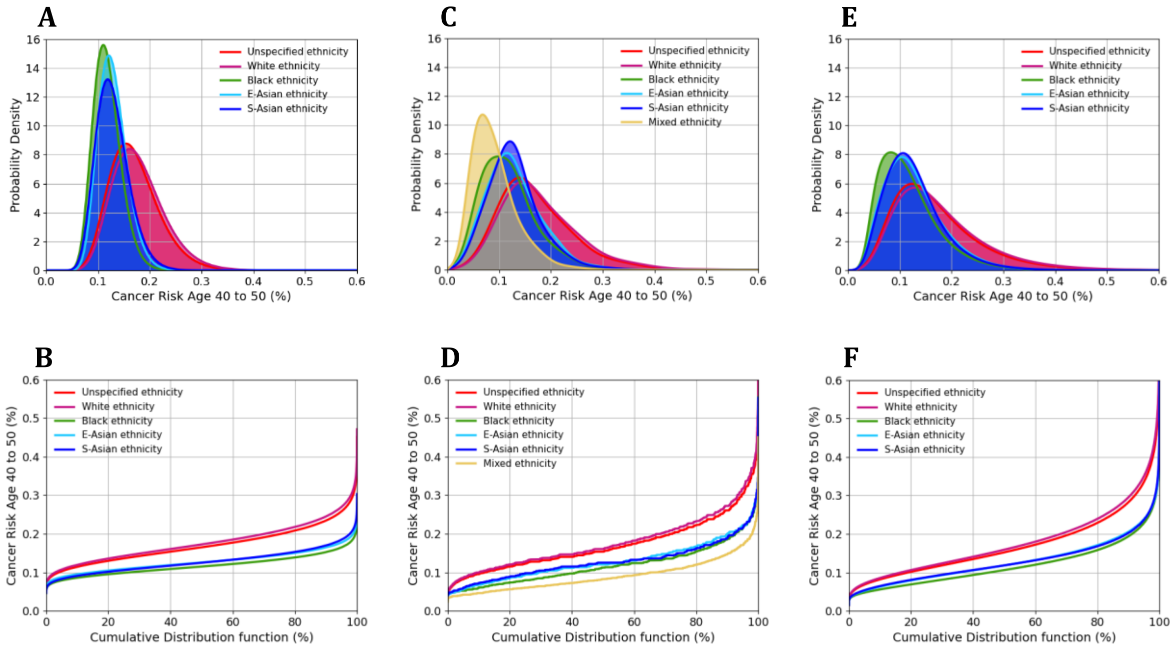

**Figure S16.** Predicted 10-year (to age 50 years) EOC risks for women of different ethnicities, untested for PVs and unknown family history. Risks are based on different combinations of risk predictors, each drawn in a separate panel: (A,B) PGS only; (C,D) QRFs only; (E,F) PGS and QRFs. (A,C,E) Probability density function against absolute risk; (B,D,F) absolute risk against cumulative distribution. EOC, epithelial ovarian cancer; PGS, polygenic score; QRFs, questionnaire-based risk factors.

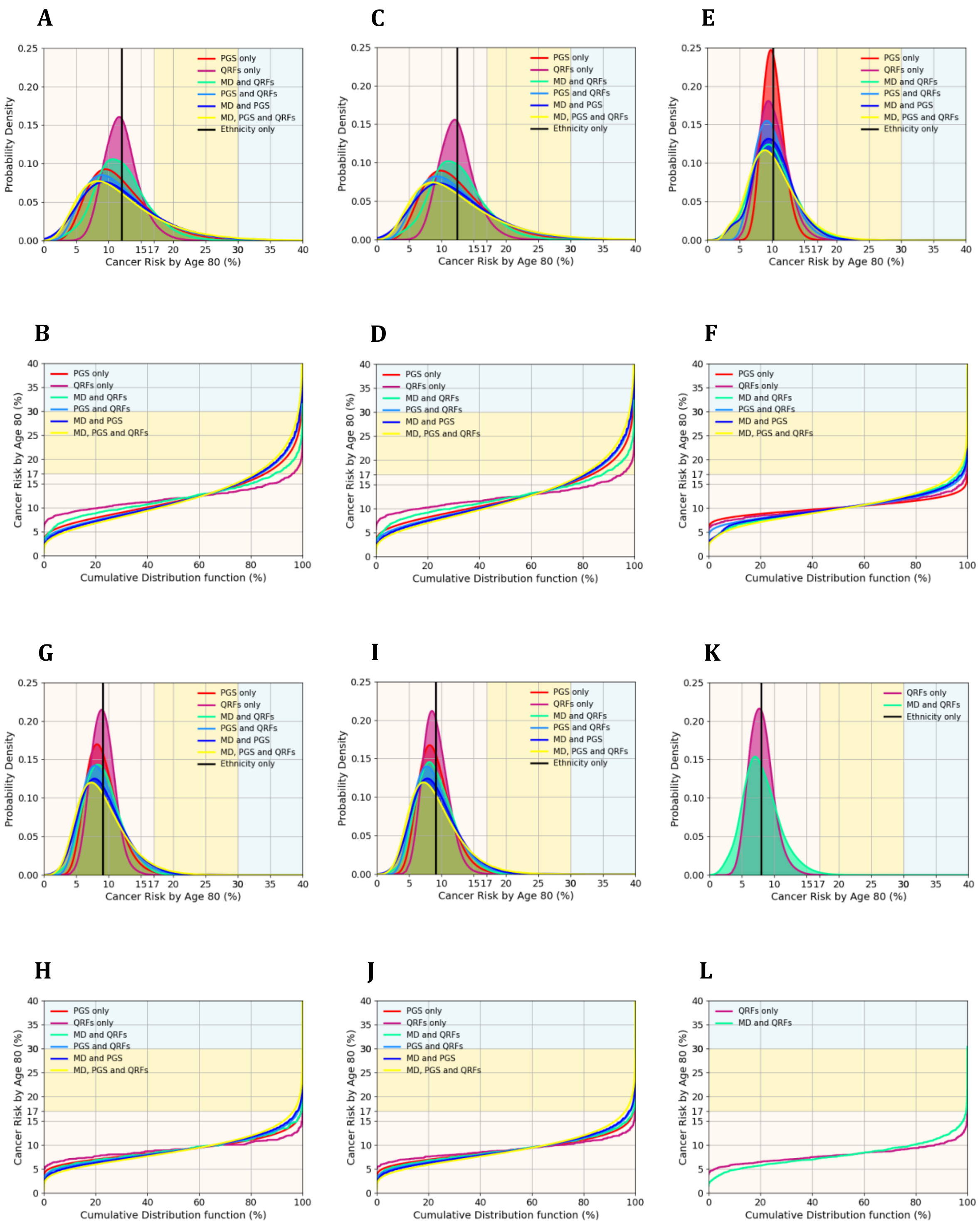

**Figure S17.** Predicted lifetime (age 20-80 years) BC risks for women of different ethnicities, untested for PVs and unknown family history, based on different combinations of risk predictors. Distributions for each ethnicity are drawn in separate panels: (A,B) Population/Unspecified; (C,D) White; (E,F) Black; (G,H) East Asian; (I,J) South Asian; (K,L) Mixed. The straight lines (labelled 'ethnicity only') are equivalent to the "population" risk of BC for the specific ethnicity plotted. (A,C,E,G,I,K) Probability density function against absolute risk; (B,D,F,H,J,L) absolute risk against cumulative distribution. BC, breast cancer; PGS, Polygenic Score; QRFs, questionnaire-based risk factors; MD, mammographic density. The backgrounds of the graphs are shaded to indicate three lifetime BC risk categories: less than 17% (light yellow); between 17% and 30% (yellow); above 30% (light blue).

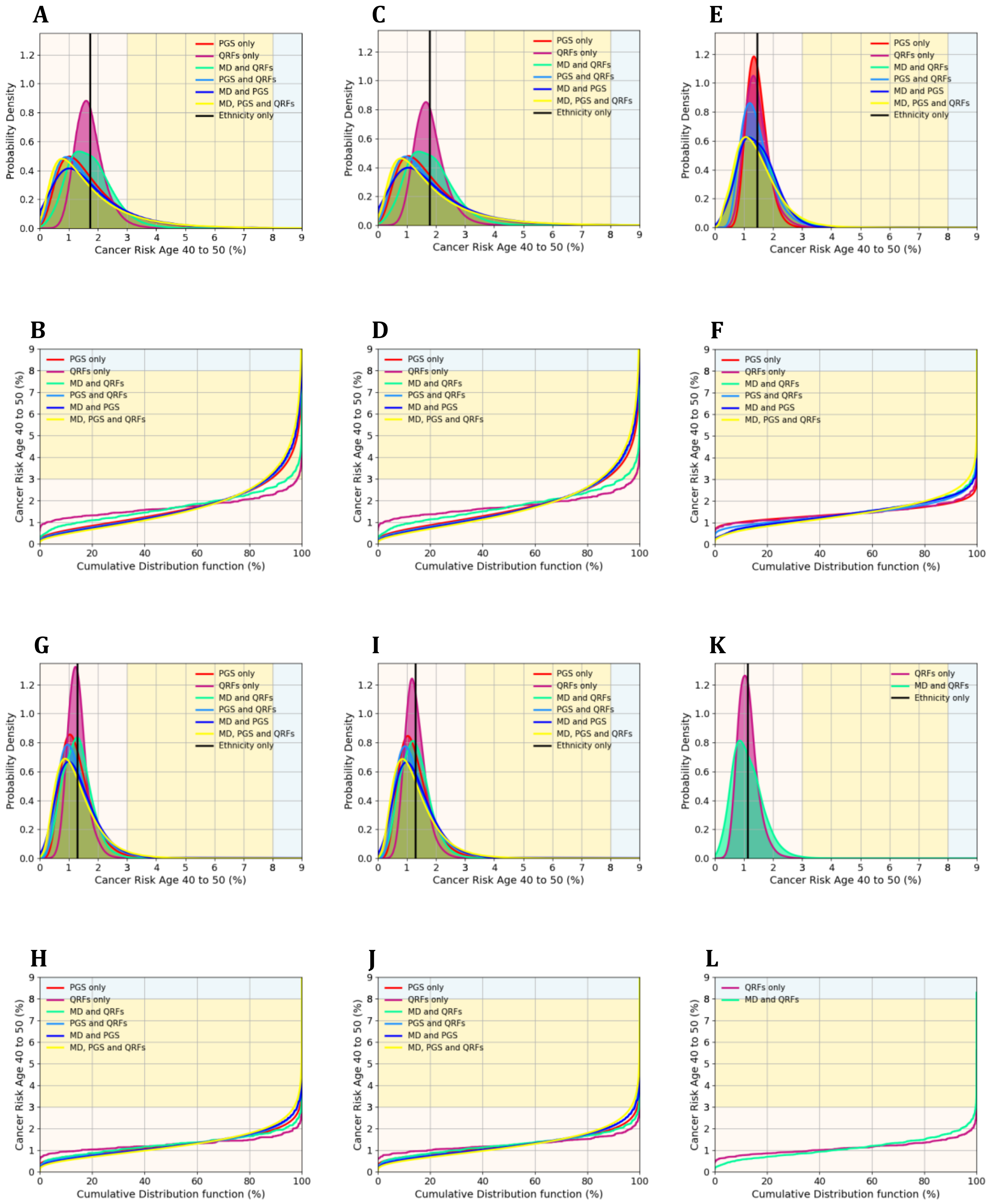

**Figure S18.** Predicted 10-year (to age 50 years) BC risks for women of different ethnicities, untested for PVs and unknown family history, based on different combinations of risk predictors. Distributions for each ethnicity are drawn in separate panels: (A,B) Population/Unspecified; (C,D) White; (E,F) Black; (G,H) East Asian; (I,J) South Asian; (K,L) Mixed. The straight lines (labelled ‘ethnicity only’) are equivalent to the “population” risk of BC for the specific plotted. (A,C,E,G,I,K) Probability density function against absolute risk; (B,D,F,H,J,L) absolute risk against cumulative distribution. BC, breast cancer; PGS, polygenic score; QRFs, questionnaire-based risk factors; MD, mammographic density. The backgrounds of the graphs are shaded to indicate three 10-year BC risk categories: less than 3% (light yellow); between 3% and 8% (yellow); above 8% (light blue).

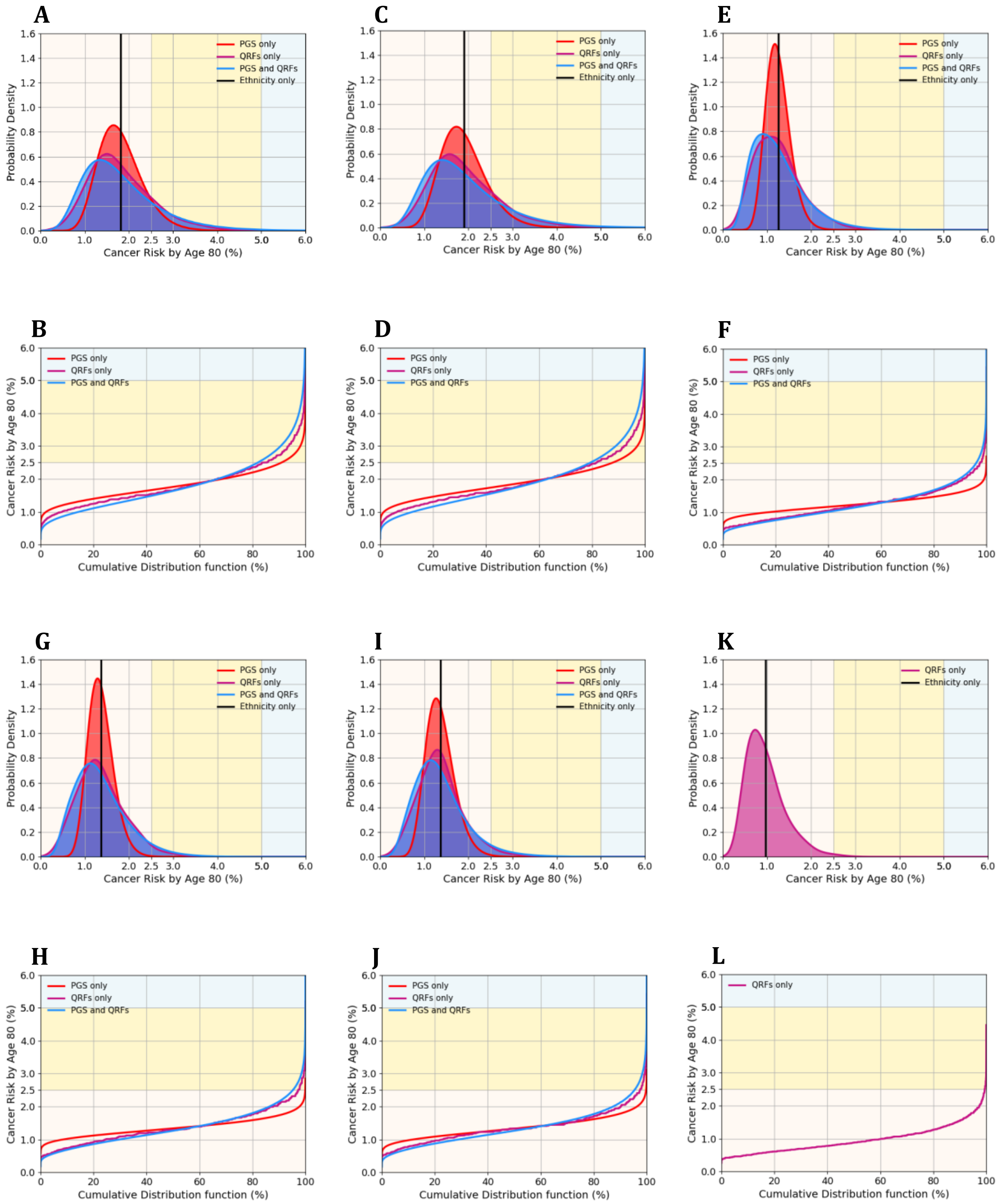

**Figure S19.** Predicted lifetime (age 20-80 years) EOC risks for women of different ethnicities, untested for PVs and unknown family history, based on different combinations of risk predictors. Distributions for each ethnicity are drawn in separate panels: (A,B) Population/Unspecified; (C,D) White; (E,F) Black; (G,H) East Asian; (I,J) South Asian; (K,L) Mixed. The straight lines (labelled 'ethnicity only') are equivalent to the "population" risk of EOC for the specific ethnicity plotted. (A,C,E,G,I,K) Probability density function against absolute risk; (B,D,F,H,J,L) absolute risk against cumulative distribution. EOC, epithelial ovarian cancer; PGS, polygenic score; QRFs, questionnaire-based risk factors. The backgrounds of the graphs are shaded to indicate three lifetime EOC risk categories: less than 2.5% (light yellow); between 2.5% and 5% (yellow); above 5% (light blue).

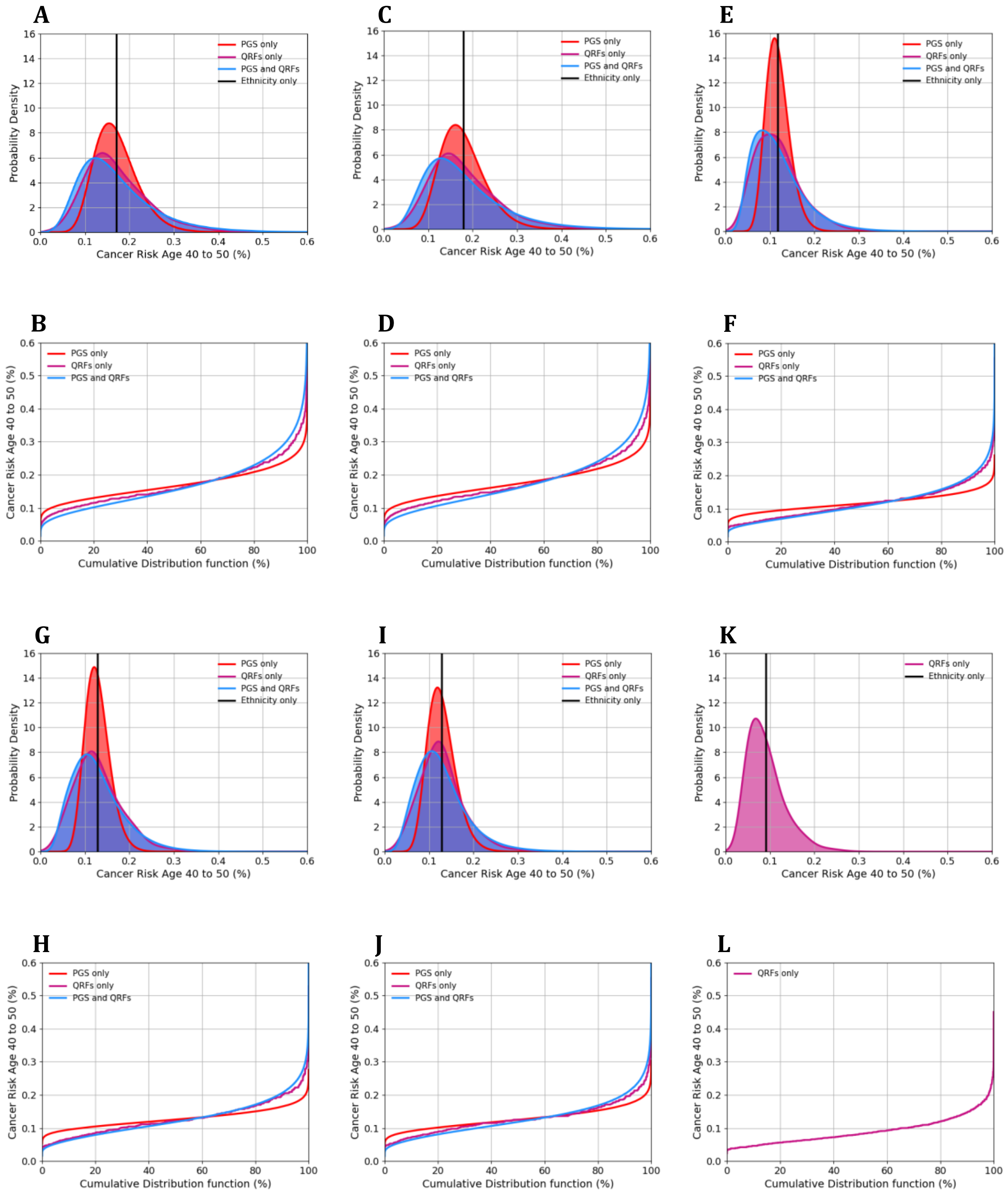

**Figure S20.** Predicted 10-year (to age 50 years) EOC risks for women of different ethnicities, untested for PVs and unknown family history, based on different combinations of risk predictors. Distributions for each ethnicity are drawn in separate panels: (A,B) Population/Unspecified; (C,D) White; (E,F) Black; (G,H) East Asian; (I,J) South Asian; (K,L) Mixed. The straight lines (labelled ‘ethnicity only’) are equivalent to the “population” risk of EOC for the specific ethnicity plotted. (A,C,E,G,I,K) Probability density function against absolute risk; (B,D,F,H,J,L) absolute risk against cumulative distribution. EOC, epithelial ovarian cancer; PGS, polygenic score; QRFs, questionnaire-based risk factors.

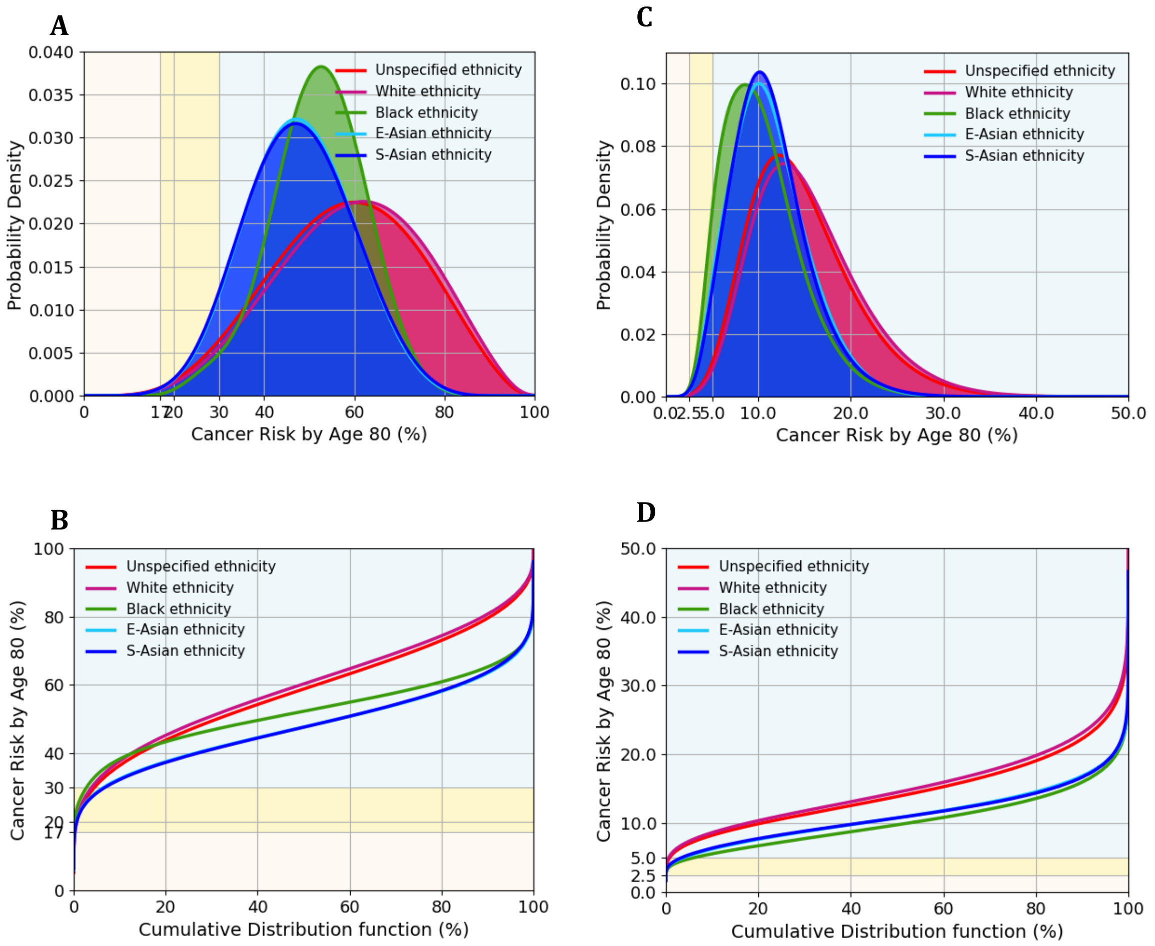

**Figure S21.** Predicted BC and EOC lifetime (age 20-80 years) risks for women of different ethnicities, *BRCA2* PV carrier and unknown family history; risks calculated using all risk predictors: PGS, QRFs, and MD. (A,B) BC risk; backgrounds are shaded to indicate three risk categories: <17% (light yellow); 17% - 30% (yellow); ≥ 30% (light blue). (C,D) EOC risk; backgrounds are shaded to indicate three risk categories: < 2.5% (light yellow); 2.5% - 5% (yellow); ≥ 5% (light blue). (A,C) probability density function against absolute risk; (B,D) absolute risk against cumulative distribution. BC, breast cancer; EOC, epithelial ovarian cancer; PGS, polygenic score; QRFs, questionnaire-based risk factors; MD, mammographic density.

### Tables

**Table S1.** Ethnicity proportions ($\boldsymbol{P_{ethnicity}}$) obtained from the ONS (21) and incidence rate ratios by ethnic group and cancer type ($\boldsymbol{R_{ethnicity}}$), relative to White (20). East and South Asian ethnicities were grouped in a single group due to small numbers.

| **Ethnicities** | *White* | *Black* | *Asian* | *Mixed* | *Other* |
|---|---|---|---|---|---|
| ***Proportion (2016 census)*** | 0.851 | 0.035 | 0.078 | 0.0176 | 0.018 |
| ***Breast cancer rate ratio (women)*** | 1.00 | 0.81 | 0.72 | 0.63 | NA |
| ***Ovarian cancer rate ratio*** | 1.00 | 0.66 | 0.72 | 0.51 | NA |
| ***Prostate cancer rate ratio*** | 1.00 | 2.10 | 0.54 | 0.78 | NA |
| ***Pancreatic cancer rate ratio (women)*** | 1.00 | 1.20 | 0.62 | 0.76 | NA |
| ***Pancreatic cancer rate ratio (men)*** | 1.00 | 1.09 | 0.67 | 0.76 | NA |

**Table S2.** Summary of the BCSC dataset used: distribution of women across all 'races' and 'ethnicities', by age group (split age: 50 years) and by diagnosis within 1 year from the mammogram screening.

| **'Race', 'ethnicity'** | **Age <50 years** | | **Age >=50 years** | |
|---|---|---|---|---|
| | *All women* | *BC within 1y* | *All women* | *BC within 1y* |
| *White, non-Hispanic* | 170520 | 523 | 240195 | 1913 |
| *Black, non-Hispanic* | 15310 | 37 | 15861 | 103 |
| *Asian, non-Hispanic* | 11648 | 20 | 16180 | 61 |
| *Mixed, non-Hispanic* | 4413 | 5 | 6950 | 26 |
| *Unknown, non-Hispanic* | 7342 | 23 | 11294 | 60 |
| *White, Hispanic* | 18103 | 54 | 29456 | 156 |
| *Black, Hispanic* | 89 | 0 | 74 | 1 |
| *Asian, Hispanic* | 167 | 0 | 241 | 3 |
| *Mixed, Hispanic* | 1470 | 4 | 1704 | 5 |
| *Unknown, Hispanic* | 4174 | 10 | 4515 | 19 |
| *White, unknown* | 15971 | 33 | 21222 | 144 |
| *Black, unknown* | 472 | 0 | 886 | 6 |
| *Asian, unknown* | 2259 | 8 | 5674 | 27 |
| *Mixed, unknown* | 2154 | 5 | 4415 | 6 |
| *Unknown, unknown* | 39425 | 120 | 66705 | 449 |

**Table S3.** Summary of the BCSC dataset used, after imputation: distribution of women across all ‘races’ and ‘ethnicities’, by age group (split age: 50 years) and by diagnosis within 1 year from the mammogram screening.

| **‘Race’, ‘ethnicity’** | **Age <50 years** | | **Age >=50 years** | |
|---|---|---|---|---|
| | *All women* | *BC within 1y* | *All women* | *BC within 1y* |
| *White, non-Hispanic* | 184793 | 553 | 258990 | 2041 |
| *Black, non-Hispanic* | 15732 | 37 | 16649 | 109 |
| *Asian, non-Hispanic* | 13680 | 27 | 21215 | 85 |
| *Mixed, non-Hispanic* | 6319 | 10 | 10837 | 32 |
| *Unknown, non-Hispanic* | 42602 | 130 | 70544 | 461 |
| *White, Hispanic* | 19801 | 57 | 31883 | 172 |
| *Black, Hispanic* | 139 | 0 | 172 | 1 |
| *Asian, Hispanic* | 394 | 1 | 880 | 6 |
| *Mixed, Hispanic* | 1718 | 4 | 2232 | 6 |
| *Unknown, Hispanic* | 8339 | 23 | 11970 | 67 |

**Table S16.** Summary of published associations between breast cancer and rare pathogenic variants by different ancestries. Odds ratios [95% confidence intervals] were reported.

| | **European ancestry** | **Asian ancestry** | | | **African ancestry** | |
|---|---|---|---|---|---|---|
| ***Data source*** | *BRIDGES (31)* | *BRIDGES (31)* | *Japanese study (35)* | *Asian families (36)* | *GBHS (33)* | *Afr. Am. study (34)* |
| ***Cases #*** | 42060 | 6279 | 7051 | 572 families | 871 | 5054 |
| ***Controls #*** | 44035 | 6047 | 11241 | | 1563 | 4993 |
| ***BRCA1*** | 9.3 [7.0-12.4] | 22.1 [6.9-70.5] | 33.0 [13.7-103.8] | 15.6 [12.6-19.4] | 13.7 [4.0-46.5] | 47.6 [10.4-100] |
| ***BRCA2*** | 5.4 [4.4-6.6] | 8.2 [4.9-13.6] | 16.4 [10.2-28.0] | 10.3 [8.3-12.7] | 7.0 [3.2-15.5] | 7.3 [4.1-14.1] |
| ***PALB2*** | 5.0 [3.6-6.9] | 4.5 [2.0-10.2] | 9.0 [3.4-29.7] | NA | 17.3 [2.2-138.1] | 8.5 [3.7-25.0] |
| ***ATM*** | 2.1 [1.7-2.6] | 2.3 [1.2-4.8] | 2.1 [1.0-4.1] | NA | 1.6 [0.4-6.1] | 1.8 [1.0-3.4] |
| ***CHEK2*** | 2.6 [2.2-3.0] | 1.5 [0.6-3.8] | 3.2 [1.6-6.8] | NA | NA | NA |
| ***BARD1*** | 2.0 [1.3-3.2] | 2.5 [0.8-8.0] | NA | NA | NA | 0.8 [0.3-2.3] |
| ***RAD51C*** | 1.9 [1.1-3.3] | 2.0 [0.8-5.0] | NA | NA | 2.8 [0.2-45.7] | 3.0 [0.9-13.9] |
| ***RAD51D*** | 1.7 [0.9-2.9] | 2.3 [0.9-5.9] | NA | NA | NA | 2.9 [0.7-19.9] |

**Table S17.** Summary of published pathogenic variant frequencies (%) in the population by different ancestries without large rearrangement sensitivity adjustment.

| | European ancestry | | Asian ancestry | | | African ancestry | |
|---|---|---|---|---|---|---|---|
| ***Data source*** | *BRIDGES (31)* | *CARRIERS (32)* | *BRIDGES (31)* | *CARRIERS (32)* | *Japanese (35)* | *GBHS (33)* | *CARRIERS (32)* |
| ***Sample size*** | 44035 | 24770 | 6047 | 1269 | 11241 | 1563 | 4954 |
| ***BRCA1*** | 0.06 | 0.14 | 0.02 | 0 | 0.04 | 0.19 | 0.02 |
| ***BRCA2*** | 0.13 | 0.23 | 0.14 | 0.39 | 0.17 | 0.51 | 0.24 |
| ***PALB2*** | 0.05 | 0.13 | 0.06 | 0.18 | 0.04 | 0.06 | 0.10 |
| ***ATM*** | 0.16 | 0.42 | 0.09 | 0.39 | 0.15 | 0.32 | 0.34 |
| ***CHEK2*** | 0.35 | 0.50 | 0.07 | 0.24 | 0.12 | 0 | 0.14 |
| ***BARD1*** | 0.03 | 0.10 | 0.03 | 0.08 | NA | 0 | 0.16 |
| ***RAD51C*** | 0.02 | 0.10 | 0.06 | 0.16 | NA | 0.06 | 0.08 |
| ***RAD51D*** | 0.02 | 0.04 | 0.05 | 0 | NA | 0 | 0.06 |

**Table S18.** Estimated QRFs distributions in UK Biobank and MD distribution from BCSC. QRFs: questionnaire-based risk factors; MD: mammographic density; HRT: hormone replacement therapy; OC: oral contraceptive.

| Risk factors | Category | White | Black | East Asian | South Asian | Mixed | BOADICEA (BC v6, EOC v2) |
|---|---|---|---|---|---|---|---|
| **Risk factors used in both breast and ovarian cancer risk prediction** | | | | | | | |
| **HRT use** | *Current C-type* | 0.189 | 0.081 | 0.091 | 0.09 | 0.134 | 0.076 |
| | *Current E-type* | 0.001 | 0.001 | 0.001 | 0 | 0 | 0.011 |
| | *Former* | 0.203 | 0.09 | 0.094 | 0.111 | 0.113 | 0.913 |
| | *Never* | 0.607 | 0.828 | 0.814 | 0.799 | 0.752 | |
| **Height** | *[0,153)* | 0.05 | 0.059 | 0.197 | 0.242 | 0.071 | 0.0625 |
| | *[153,160)* | 0.258 | 0.271 | 0.442 | 0.438 | 0.28 | 0.2500 |
| | *[160,166)* | 0.376 | 0.373 | 0.279 | 0.25 | 0.366 | 0.3750 |
| | *[166,173)* | 0.258 | 0.242 | 0.077 | 0.065 | 0.23 | 0.2500 |
| | *>=173* | 0.058 | 0.056 | 0.004 | 0.005 | 0.052 | 0.0625 |
| | *Mean* | 162.63 | 162.36 | 157.49 | 156.71 | 161.97 | 162.81 |
| | *SD* | 6.24 | 6.30 | 5.59 | 5.93 | 6.39 | 6.45 |
| **Parity** | *0* | 0.188 | 0.167 | 0.222 | 0.119 | 0.243 | 0.28 |
| | *1* | 0.133 | 0.175 | 0.247 | 0.108 | 0.173 | 0.16 |
| | *2* | 0.445 | 0.263 | 0.375 | 0.377 | 0.34 | 0.33 |
| | *>=3* | 0.235 | 0.395 | 0.156 | 0.395 | 0.244 | 0.23 |
| **Risk factors used in breast cancer risk prediction** | | | | | | | |
| **Age at menarche** | *<11* | 0.046 | 0.058 | 0.038 | 0.049 | 0.068 | 0.122 |
| | *11* | 0.156 | 0.118 | 0.11 | 0.116 | 0.157 | 0.216 |
| | *12* | 0.190 | 0.183 | 0.252 | 0.163 | 0.193 | 0.268 |
| | *13* | 0.245 | 0.211 | 0.227 | 0.237 | 0.236 | 0.215 |
| | *14* | 0.198 | 0.169 | 0.185 | 0.193 | 0.177 | 0.115 |
| | *15* | 0.109 | 0.136 | 0.103 | 0.129 | 0.104 | 0.044 |
| | *>15* | 0.056 | 0.124 | 0.085 | 0.112 | 0.065 | 0.019 |
| **Age at menopause** | *[0,40)* | 0.04 | 0.062 | 0.027 | 0.045 | 0.058 | 0.022 |
| | *[40,45)* | 0.09 | 0.131 | 0.07 | 0.128 | 0.102 | 0.070 |
| | *[45,50)* | 0.236 | 0.339 | 0.24 | 0.338 | 0.301 | 0.234 |
| | *[50,55)* | 0.481 | 0.343 | 0.539 | 0.377 | 0.435 | 0.533 |
| | *>=55* | 0.152 | 0.126 | 0.124 | 0.112 | 0.104 | 0.14 |
| **Age at first live birth** | *[0,20)* | 0.097 | 0.292 | 0.041 | 0.115 | 0.209 | 0.1667 |
| | *[20,25)* | 0.347 | 0.344 | 0.222 | 0.422 | 0.308 | 0.3194 |
| | *[25,30)* | 0.379 | 0.243 | 0.406 | 0.321 | 0.29 | 0.3194 |
| | *>=30* | 0.177 | 0.122 | 0.331 | 0.143 | 0.193 | 0.1944 |
| **MD (BIRADS $4^{th}$), age <50y** | *A* | 0.044 | 0.057 | *0.014* | *0.014* | 0.049 | 0.041 |
| | *B* | 0.351 | 0.367 | *0.226* | *0.226* | 0.394 | 0.352 |
| | *C* | 0.466 | 0.447 | *0.518* | *0.518* | 0.445 | 0.470 |
| | *D* | 0.139 | 0.099 | *0.242* | *0.242* | 0.113 | 0.137 |
| **MD (BIRADS $4^{th}$), age >=50y** | *A* | 0.097 | 0.105 | *0.108* | *0.108* | 0.125 | 0.041 |
| | *B* | 0.491 | 0.502 | *0.434* | *0.434* | 0.542 | 0.352 |
| | *C* | 0.359 | 0.355 | *0.368* | *0.368* | 0.296 | 0.470 |
| | *D* | 0.053 | 0.038 | *0.090* | *0.090* | 0.037 | 0.137 |

| | | | | | | | |
|---|---|---|---|---|---|---|---|
| **BMI (kg/m2)** | *[0,18.5)* | 0.008 | 0.002 | 0.03 | 0.007 | 0.009 | 0.033 |
| | *[18.5,25)* | 0.394 | 0.182 | 0.687 | 0.319 | 0.405 | 0.357 |
| | *[25,30)* | 0.367 | 0.346 | 0.242 | 0.414 | 0.343 | 0.358 |
| | *>=30* | 0.231 | 0.47 | 0.041 | 0.26 | 0.243 | 0.252 |
| **OC use** | *Current* | 0.038 | 0.05 | 0.043 | 0.031 | 0.073 | 0.131 |
| | *Former* | 0.783 | 0.677 | 0.512 | 0.484 | 0.772 | 0.684 |
| | *Never* | 0.179 | 0.273 | 0.445 | 0.485 | 0.155 | 0.185 |
| **Alcohol (g/day)** | *0* | 0.102 | 0.335 | 0.488 | 0.672 | 0.170 | 0.106 |
| | *(0,5)* | 0.207 | 0.372 | 0.318 | 0.200 | 0.276 | 0.307 |
| | *[5,15)* | 0.400 | 0.205 | 0.134 | 0.093 | 0.320 | 0.276 |
| | *[15,25)* | 0.179 | 0.054 | 0.040 | 0.019 | 0.138 | 0.133 |
| | *[25,35)* | 0.067 | 0.021 | 0.013 | 0.008 | 0.050 | 0.080 |
| | *[35,45)* | 0.025 | 0.006 | 0.003 | 0.004 | 0.015 | 0.036 |
| | *>=45* | 0.020 | 0.008 | 0.003 | 0.003 | 0.031 | 0.061 |
| **Risk factors used in ovarian cancer risk prediction** | | | | | | | |
| **Endometriosis** | *No* | 0.985 | 0.988 | 0.990 | 0.991 | 0.980 | 0.90 |
| | *Yes* | 0.015 | 0.012 | 0.010 | 0.009 | 0.020 | 0.10 |
| **Tubal ligation** | *No* | 0.994 | 0.991 | 0.999 | 0.996 | 0.994 | 0.77 |
| | *Yes* | 0.006 | 0.009 | 0.001 | 0.004 | 0.006 | 0.23 |
| **OC use duration** | *Never, <1* | 0.234 | 0.367 | 0.532 | 0.572 | 0.222 | 0.710 |
| | *1-4* | 0.170 | 0.148 | 0.158 | 0.149 | 0.156 | 0.138 |
| | *5-9* | 0.200 | 0.149 | 0.109 | 0.114 | 0.189 | 0.086 |
| | *10-14* | 0.172 | 0.129 | 0.081 | 0.077 | 0.149 | 0.046 |
| | *≥15* | 0.224 | 0.208 | 0.121 | 0.088 | 0.284 | 0.020 |
| **BMI** | *<22.5* | 0.172 | 0.059 | 0.433 | 0.128 | 0.190 | 0.325 |
| | *22.5-<30* | 0.598 | 0.470 | 0.526 | 0.612 | 0.567 | 0.529 |
| | *≥30* | 0.231 | 0.470 | 0.041 | 0.260 | 0.243 | 0.146 |

**Table S19.** Odds ratios [95% confidence intervals] for different BIRADS categories, estimated for each ethnicity separately. BIRADS B is the reference category for individuals of age less than 50 years (upper panel); BIRADS A is the reference category for individuals of age 50 years and over (lower panel).

| ORs | Category A | Category B | Category C | Category D |
|---|---|---|---|---|
| *White, <50* | 0.48 [0.19-1.24] | 1 | 1.23 [0.98-1.54] | 1.45 [1.04-2.01] |
| *Black, <50* | 0.75 [0.10-5.68] | 1 | 1.17 [0.51-2.65] | 1.76 [0.58-5.37] |
| *Asian, <50* | 1.76 [0.27-11.40] | 1 | 1.24 [0.31-5.01] | 1.03 [0.19-5.58] |
| *Mixed, <50* | NA | 1 | 1.06 [0.18-6.32] | 1.35 [0.21-8.66] |
| *White, >=50* | 1 | 1.39 [1.11-1.74] | 1.76 [1.39-2.22] | 1.79 [1.33-2.40] |
| *Black, >=50* | 1 | 1.68 [0.65-4.37] | 1.92 [0.640-5.735] | 1.756 [0.47-6.59] |
| *Asian, >=50* | 1 | 1.41 [0.53-3.74] | 1.52 [0.444-5.220] | 1.214 [0.21-6.98] |
| *Mixed, >=50* | 1 | 1.35 [0.34-5.36] | 1.51 [0.372-6.102] | NA |

**Table S20.** Ethnicity-specific Odds ratio estimates [95% confidence intervals], using entire BCSC population and considering an interaction term between MD and ethnicity. BIRADS B is the reference category for individuals of age less than 50 years (upper panel); BIRADS A is the reference category for individuals of age 50 years and over (lower panel).

| ORs | Category A | Category B | Category C | Category D |
|---|---|---|---|---|
| *White, <50* | 0.45 [0.19-1.35] | 1 | 1.24 [0.99-1.54] | 1.46 [1.03-2.04] |
| *Black, <50* | 1.67 [0.18-17.22] | 1 | 1.13 [0.33-4.03] | 0.95 [0.20-4.55] |
| *Asian, <50* | 1.12 [0.30-4.23] | 1 | 1.79 [0.98-3.16] | 2.08 [0.97-4.26] |
| *Mixed, <50* | 0.67 [0.08-6.44] | 1 | 1.16 [0.40-3.14] | 1.62 [0.47-5.69] |
| *White, >=50* | 1 | 1.38 [1.13-1.67] | 1.72 [1.39-2.13] | 1.75 [1.32-2.32] |
| *Black, >=50* | 1 | 1.50 [0.50-4.50] | 1.48 [0.48-4.61] | 1.28 [0.24-6.76] |
| *Asian, >=50* | 1 | 1.93 [1.13-3.31] | 2.15 [1.25-3.72] | 3.80 [1.98-7.27] |
| *Mixed, >=50* | 1 | 1.62 [0.57-4.61] | 1.82 [0.61-5.41] | 1.83 [0.41-8.11] |

**Table S21.** Proportion [95% confidence intervals] of BIRADS categories in different ethnic groups, for individuals of age less than 50 years (upper panel) and for individuals of age 50 years and over (lower panel).

| Proportions | Category A | Category B | Category C | Category D |
|---|---|---|---|---|
| *White, <50* | 0.044 [0.043-0.045] | 0.351 [0.348-0.353] | 0.466 [0.464-0.468] | 0.139 [0.137-0.141] |
| *Asian, <50* | 0.014 [0.011-0.017] | 0.226 [0.216-0.236] | 0.518 [0.506-0.529] | 0.242 [0.232-0.252] |
| *Black, <50* | 0.057 [0.053-0.060] | 0.367 [0.360-0.374] | 0.477 [0.470-0.484] | 0.099 [0.095-0.104] |
| *Mixed, <50* | 0.049 [0.042-0.056] | 0.394 [0.377-0.410] | 0.445 [0.428-0.461] | 0.113 [0.102-0.123] |
| *White, >=50* | 0.097 [0.096-0.098] | 0.491 [0.489-0.493] | 0.359 [0.357-0.360] | 0.053 [0.052-0.054] |
| *Asian, >=50* | 0.108 [0.103-0.113] | 0.434 [0.426-0.442] | 0.368 [0.361-0.376] | 0.090 [0.085-0.094] |
| *Black, >=50* | 0.105 [0.101-0.109] | 0.502 [0.496-0.508] | 0.355 [0.349-0.361] | 0.038 [0.036-0.041] |
| *Mixed, >=50* | 0.125 [0.117-0.134] | 0.542 [0.530-0.555] | 0.296 [0.285-0.307] | 0.037 [0.032-0.041] |

**Table S22.** Source of the model parameters for each ethnic/ancestry group (as described in this manuscript). *'default'* indicates parameters in previous BOADICEA models (BC v6, EOC v2).

| **Ethnicity** | **CHEK2 PV frequencies** | **PGS parameters** | **MD distribution** | **Other QRFs distribution** | **ER and TN status** | **Incidences** |
|---|---|---|---|---|---|---|
| *Source (section)* | 'Rare genetic variants' | Table 2 | Table S18 | Table S18 | Tables S4-S6 | Table 1 |
| *Unspecified* | *default* | *default* | *default* | *default* | *default* | *default* |
| *White* | *default* | European | White | *default* | White | White |
| *Black* | non-European | African | Black | Black | Black | Black |
| *East Asian* | non-European | East Asian | Asian | East Asian | East Asian | Asian |
| *South Asian* | non-European | South Asian | Asian | South Asian | South Asian | Asian |
| *Mixed* | non-European | *(derived)* | Mixed | Mixed | Mixed | Mixed |
| *Other* | non-European | *default* | *default* | *default* | *default* | *default* |

**Table S23.** Proportion of women falling in different risk categories depending on their lifetime cancer risk (age 20 to 80 years) and their ethnicity, using different risk predictors. Unknown genetic background, mother with BC/EOC at 50y. First panel: lifetime BC risk; the thresholds between risk categories are 17%, between ‘population’ and ‘moderate’, and 30%, between ‘moderate’ and ‘high’ (NICE guidelines). Second panel: lifetime EOC risk, with thresholds at 3.5% and 5% (threshold for risk-reducing surgery, NICE guidelines).

| **Predictors** | **Risk categories** | **Proportions in each category based on lifetime BC risk** | | | | | |
|---|---|---|---|---|---|---|---|
| | | *UK pop* | *White* | *Black* | *East Asian* | *South Asian* | *Mixed* |
| PGS only | *Population* | 0.519 | 0.483 | 0.635 | 0.797 | 0.795 | --- |
| | *Moderate* | 0.446 | 0.473 | 0.365 | 0.203 | 0.205 | --- |
| | *High* | 0.036 | 0.044 | 0.000 | 0.001 | 0.000 | --- |
| QRFs only | *Population* | 0.272 | 0.232 | 0.571 | 0.790 | 0.776 | 0.911 |
| | *Moderate* | 0.721 | 0.762 | 0.429 | 0.210 | 0.224 | 0.089 |
| | *High* | 0.006 | 0.006 | 0.001 | 0.000 | 0.000 | 0.000 |
| PGS+QRFs | *Population* | 0.532 | 0.498 | 0.604 | 0.757 | 0.755 | --- |
| | *Moderate* | 0.413 | 0.439 | 0.394 | 0.241 | 0.243 | --- |
| | *High* | 0.055 | 0.063 | 0.002 | 0.001 | 0.002 | --- |
| PGS+MD | *Population* | 0.535 | 0.503 | 0.571 | 0.741 | 0.741 | --- |
| | *Moderate* | 0.402 | 0.424 | 0.427 | 0.258 | 0.258 | --- |
| | *High* | 0.063 | 0.072 | 0.001 | 0.001 | 0.001 | --- |
| QRFs+MD | *Population* | 0.376 | 0.336 | 0.560 | 0.700 | 0.695 | 0.813 |
| | *Moderate* | 0.587 | 0.616 | 0.429 | 0.300 | 0.303 | 0.186 |
| | *High* | 0.037 | 0.048 | 0.010 | 0.001 | 0.001 | 0.001 |
| PGS+QRFs+MD | *Population* | 0.542 | 0.513 | 0.585 | 0.724 | 0.724 | --- |
| | *Moderate* | 0.380 | 0.399 | 0.400 | 0.268 | 0.266 | --- |
| | *High* | 0.078 | 0.088 | 0.015 | 0.008 | 0.009 | --- |

| **Predictors** | **Risk categories** | **Proportions in each category based on lifetime EOC risk** | | | | | |
|---|---|---|---|---|---|---|---|
| | | *UK pop* | *White* | *Black* | *East Asian* | *South Asian* | *Mixed* |
| PGS only | *<3.5%* | 0.038 | 0.026 | 0.348 | 0.157 | 0.199 | --- |
| | *3.5-5%* | 0.503 | 0.444 | 0.634 | 0.782 | 0.721 | --- |
| | *≥5%* | 0.459 | 0.530 | 0.018 | 0.060 | 0.080 | --- |
| QRFs only | *<3.5%* | 0.148 | 0.144 | 0.447 | 0.310 | 0.285 | 0.689 |
| | *3.5-5%* | 0.405 | 0.365 | 0.389 | 0.462 | 0.516 | 0.240 |
| | *≥5%* | 0.447 | 0.491 | 0.164 | 0.228 | 0.200 | 0.071 |
| PGS+QRFs | *<3.5%* | 0.186 | 0.188 | 0.486 | 0.375 | 0.356 | --- |
| | *3.5-5%* | 0.379 | 0.346 | 0.345 | 0.395 | 0.436 | --- |
| | *≥5%* | 0.435 | 0.466 | 0.169 | 0.229 | 0.208 | --- |

**Table S24.** Proportion of women falling in different risk categories depending on their 10-year BC risk (age 40 to 50 years) and their ethnicity, using different risk predictors. Unknown genetic background, unknown FH. The thresholds between risk categories are 3%, between 'population' and 'moderate', and 8%, between 'moderate' and 'high' (NICE guidelines).

| **Predictors** | **Risk categories** | **Proportions in each category based on 10-year BC risk** | | | | | |
|---|---|---|---|---|---|---|---|
| | | *UK pop* | *White* | *Black* | *East Asian* | *South Asian* | *Mixed* |
| PGS only | *Population* | 0.882 | 0.870 | 1.000 | 0.992 | 0.991 | --- |
| | *Moderate* | 0.117 | 0.129 | 0.000 | 0.008 | 0.009 | --- |
| | *High* | 0.001 | 0.001 | 0.000 | 0.000 | 0.000 | --- |
| QRFs only | *Population* | 0.982 | 0.983 | 0.997 | 1.000 | 1.000 | 1.000 |
| | *Moderate* | 0.018 | 0.017 | 0.003 | 0.000 | 0.000 | 0.000 |
| | *High* | 0.000 | 0.000 | 0.000 | 0.000 | 0.000 | 0.000 |
| PGS+QRFs | *Population* | 0.873 | 0.861 | 0.987 | 0.982 | 0.979 | --- |
| | *Moderate* | 0.125 | 0.137 | 0.013 | 0.018 | 0.021 | --- |
| | *High* | 0.002 | 0.003 | 0.000 | 0.000 | 0.000 | --- |
| PGS+MD | *Population* | 0.869 | 0.854 | 0.987 | 0.982 | 0.982 | --- |
| | *Moderate* | 0.130 | 0.146 | 0.013 | 0.018 | 0.018 | --- |
| | *High* | 0.001 | 0.001 | 0.000 | 0.000 | 0.000 | --- |
| QRFs+MD | *Population* | 0.944 | 0.936 | 0.984 | 0.998 | 0.996 | 0.997 |
| | *Moderate* | 0.056 | 0.064 | 0.016 | 0.002 | 0.004 | 0.003 |
| | *High* | 0.000 | 0.000 | 0.000 | 0.000 | 0.000 | 0.000 |
| PGS+QRFs+MD | *Population* | 0.860 | 0.848 | 0.964 | 0.971 | 0.967 | --- |
| | *Moderate* | 0.135 | 0.146 | 0.036 | 0.029 | 0.033 | --- |
| | *High* | 0.005 | 0.006 | 0.000 | 0.000 | 0.000 | --- |

**Table S25.** Proportion of women falling in different risk categories depending on their 10-year BC risk (age 40 to 50 years) and their ethnicity, using different risk predictors. Unknown genetic background, mother with BC at 50y. The thresholds between risk categories are 3%, between 'population' and 'moderate', and 8%, between 'moderate' and 'high' (NICE guidelines).

| **Predictors** | **Risk categories** | **Proportions in each category based on 10-year BC risk** | | | | | |
|---|---|---|---|---|---|---|---|
| | | *UK pop* | *White* | *Black* | *East Asian* | *South Asian* | *Mixed* |
| PGS only | *Population* | 0.576 | 0.552 | 0.609 | 0.756 | 0.755 | --- |
| | *Moderate* | 0.414 | 0.436 | 0.391 | 0.244 | 0.245 | --- |
| | *High* | 0.010 | 0.012 | 0.000 | 0.000 | 0.000 | --- |
| QRFs only | *Population* | 0.344 | 0.297 | 0.601 | 0.792 | 0.722 | 0.852 |
| | *Moderate* | 0.655 | 0.702 | 0.399 | 0.208 | 0.278 | 0.148 |
| | *High* | 0.001 | 0.001 | 0.000 | 0.000 | 0.000 | 0.000 |
| PGS+QRFs | *Population* | 0.593 | 0.571 | 0.610 | 0.739 | 0.737 | --- |
| | *Moderate* | 0.387 | 0.406 | 0.389 | 0.260 | 0.262 | --- |
| | *High* | 0.020 | 0.023 | 0.000 | 0.000 | 0.001 | --- |
| PGS+MD | *Population* | 0.587 | 0.569 | 0.550 | 0.714 | 0.716 | --- |
| | *Moderate* | 0.393 | 0.406 | 0.450 | 0.286 | 0.284 | --- |
| | *High* | 0.019 | 0.026 | 0.000 | 0.000 | 0.000 | --- |
| QRFs+MD | *Population* | 0.417 | 0.386 | 0.545 | 0.655 | 0.673 | 0.750 |
| | *Moderate* | 0.578 | 0.609 | 0.454 | 0.345 | 0.326 | 0.250 |
| | *High* | 0.005 | 0.006 | 0.001 | 0.000 | 0.000 | 0.000 |
| PGS+QRFs+MD | *Population* | 0.603 | 0.584 | 0.589 | 0.714 | 0.714 | --- |
| | *Moderate* | 0.366 | 0.381 | 0.408 | 0.285 | 0.284 | --- |
| | *High* | 0.031 | 0.035 | 0.003 | 0.002 | 0.002 | --- |

### Formula derivation - PGS for women of mixed genetic ancestry

#### Preliminary considerations

In BOADICEA, we assume that the PGS explains a proportion of the overall polygenic component $\alpha_i^2$ in ancestry $i$, that is the polygenic component is of the form:

$$\tau(t)\left(\alpha_i Z_i + \sqrt{1-\alpha_i^2}Y_i\right)$$

where $Y_i$ is the corresponding unknown polygenic component (also normalised) and $\tau(t)$ is the age-specific standard deviation of the polygenic component.

Assuming populations of similar structure, $\alpha_i$ is also proportional to the effect size per 1 SD, say $\gamma_i$. For mixed ancestry samples, we assume we have estimates $P_i$ of the proportion of each ancestry $i$.

For each individual, given a PGS model, the same raw PGS ($R_{pgs}$) is computed regardless of their ancestry. $R_{pgs}$ is a weighted sum of SNPs-specific genotypes:

$$R_{pgs} = \sum_{k=1}^{K} w_k g_k$$

where $g_k$ is the genotype for SNP $k$, $w_k$ is the corresponding weight and $K$ is the total number of SNPs. The weights $w_k$ are typically the $\log(relative\ risks)$ per allele, but this may not be true in general.

The raw PGS ($R_{pgs}$) is then normalised separately for each ancestry; so, the normalised PGS $Z_i$ differ by ancestry:

$$Z_i = \frac{(R_{pgs} - \mu_i)}{\sigma_i}, \qquad \mu_i = E(R_{pgs}|pop\ i), \qquad \sigma_i^2 = var(R_{pgs}|pop\ i)$$

#### Calculating *alpha* and *Z-score*

The question is how to combine appropriately the ancestry-specific Z-scores for a woman of mixed ancestry. Since the key aim is to define an appropriate risk level, the natural combined score is of the form:

$$S = \sum_{i=1}^{M} P_i \gamma_i Z_i$$

where $P_i$ is the proportion of an individual's genome derived from population $i$ (which we assume can be estimated from ancestry informative markers) and $M$ is the number of populations (ancestries) considered. The weighting by the effect size $\gamma_i$ (or, equivalently, by the $\alpha_i$) makes sense because the ancestry component in which the effect size is larger will make a greater contribution to the risk.

Since the component Z-scores $Z_i$ have mean zero, $S$ will also have mean zero.

Instead, the variance of $S$ is given by:

$$var(S) = \sum_{i=1}^{M}\sum_{j=1}^{M} cov\left(P_i\gamma_i\frac{R_{pgs}-\mu_i}{\sigma_i}, P_j\gamma_j\frac{R_{pgs}-\mu_j}{\sigma_j}\right)$$

$$= \sum_{i=1}^{M}\sum_{j=1}^{M}\frac{P_i\gamma_i}{\sigma_i}\frac{P_j\gamma_j}{\sigma_j} cov\left(R_{pgs}-\mu_i, R_{pgs}-\mu_j\right) = \sum_{i=1}^{M}\sum_{j=1}^{M}\frac{P_i\gamma_i}{\sigma_i}\frac{P_j\gamma_j}{\sigma_j} var\left(R_{pgs}\right)$$

$$= \left(\sum_{i=}^{M}\frac{P_i\gamma_i}{\sigma_i}\right)^2 var\left(R_{pgs}\right) \equiv \left(\sum_{i=}^{M}\frac{P_i\gamma_i}{\sigma_i}\right)^2 \sigma^2$$

where $\sigma^2$is the variance of the raw PGS in the admixed population.

Therefore, the final normalised Z-score is:

$$Z = \frac{S}{sd(S)} = \frac{\sum_{i=1}^{M} P_i\gamma_i Z_i}{\sigma \sum_{i=}^{M}\frac{P_i\gamma_i}{\sigma_i}}$$

The (raw) PGS effect size in population $i$, defined as the mean difference in the raw PGS between cases and controls is:

$$2\sum_{k=1}^{K} q_{ik}(1-q_{ik})w_k\beta_{ik} = \gamma_i\sigma_i$$

where $\beta_{ik}$is the per-allele effect size for SNP $i$ in population $k$ and $q_{ik}$is the corresponding allele frequency.

Therefore, the overall normalised effect size per SD for an admixed population is:

$$\gamma = \frac{\sum_{i=1}^{M} P_i\gamma_i\sigma_i}{\sigma}$$

and hence the corresponding $\alpha$, which is proportional to $\gamma$, is:

$$\alpha = \frac{\sum_{i=1}^{M} P_i\alpha_i\sigma_i}{\sigma}$$

#### Derivation of $\sigma$

We note that the variance of the raw PGS ($\sigma$) in the admixed population will depend not only on the variance within the populations but also in the variation in genotype frequencies among the populations and the extent to which ancestry at different loci is correlated. To derive $\sigma$, we consider three possible models for the population structure:

*Model A*

*An individual in an admixed population is from a given (but unknown) population i, with probability $P_i$ (i.e. the population from which the genotypes are drawn is the same for each locus).*

In this case:

$$\sigma^2 = var(R_{pgs}) = E(var(R_{pgs}|pop\ i)) + var\left(E(R_{pgs}|pop\ i)\right) = E(\sigma_i^2) + var(\mu_i)$$

$$= \sum_{i=1}^{M} P_i\sigma_i^2 + \sum_{i=1}^{M} P_i(\mu_i - \mu)^2$$

where $\mu = \sum_{i=1}^{M} P_i\mu_i$ is the overall mean of the raw PGS in the admixed population.

*Model B*

*The genotypes for each SNP are independently drawn from the M populations with probability $P_i$.* Thus, $P_i$ represents the proportion of loci for which the genotypes are drawn from population $i$, with all SNPs in the PGS being independent (we make the simplifying assumption here that the population distribution for SNPs in the PGS is likely to be similar to that for SNPs defining the ancestry proportions).

In this case:

$$\sigma_i^2 = var(R_{pgs}|pop\ i) = 2\sum_{k=1}^{K} q_{ik}(1-q_{ik})w_k^2$$

$$\sigma^2 = var(R_{pgs}) = var\left(\sum_{k=1}^{K} w_k g_k\right) = \sum_{k=1}^{K} w_k^2 var(g_k)$$

$$= \sum_{k=1}^{K} w_k^2 \left(E(var(g_k|pop\ i)) + var(E(g_k|pop\ i))\right)$$

$$= \sum_{k=1}^{K} w_k^2 \cdot E(2q_{ik}(1-q_{ik})) + \sum_{k=1}^{K} w_k^2 \cdot var(2q_{ik})$$

$$= E\left(2\sum_{k=1}^{K} q_{ik}(1-q_{ik})w_k^2\right) + 4\sum_{k=1}^{K} w_k^2 \cdot var(q_{ik})$$

$$= E(\sigma_i^2) + 4\sum_{k=1}^{K} w_k^2(E(q_{ik}{}^2) - E(q_{ik})^2)$$

$$= \sum_{i=1}^{M} P_i\sigma_i^2 + 4\sum_{k=1}^{K} w_k^2\left(\sum_{i=1}^{M} P_i q_{ik}{}^2 - \left(\sum_{i=1}^{M} P_i q_{ik}\right)^2\right)$$

*Model C*

As in model B, but allowing for some SNPs to be correlated, i.e. they are in linkage disequilibrium (LD). In this case we write the PGS a sum of contributions from *J* independent blocks of SNPs:

$$R_{pgs} = \sum_{j=1}^{J} \sum_{l} w_{jl} g_{jl}$$

where $g_{jl}$is the genotype for SNP *l* in block *j* and $w_{jl}$ is the corresponding weight.

We then define the block- and population-specific means, and the block-specific means, as:

$$\mu_{ij} = 2 \sum_{l} w_{jl} q_{ijl}\,, \qquad \mu_j = \sum_{i=1}^{M} P_i \mu_{ij}$$

So, the (conditional) mean of $R_{prs}$ in population $i$ is

$$\mu_i = E(R_{pgs} | pop\ i) = 2 \sum_{j=1}^{J} \sum_{l} w_{jl} q_{ijl} = \sum_{j=1}^{J} 2 \sum_{l} w_{jl} q_{ijl} = \sum_{j=1}^{J} \mu_{ij}$$

If we make the simplifying assumption that genotypes in the same LD block are drawn from the same population, the formula for model B can be straightforwardly generalised to:

$$\sigma^2 = var(R_{pgs}) = var\left( \sum_{j=1}^{J} \sum_{l} w_{jl} g_{jl} \right) = \sum_{j=1}^{J} \sum_{l} w_{jl}^2 var(g_{jl})$$

$$= \sum_{j=1}^{J} \sum_{l} w_{jl}^2 \Big( E\big( var(g_{jl} | pop\ i) \big) + var\big( E(g_{jl} | pop\ i) \big) \Big)$$

$$= \sum_{j=1}^{J} \sum_{l} w_{jl}^2 \cdot E\big( 2q_{ijl}(1 - q_{ijl}) \big) + \sum_{j=1}^{J} \sum_{l} w_{jl}^2 \cdot var(2q_{ijl})$$

$$= E\left( 2 \sum_{j=1}^{J} \sum_{l} q_{ijl}(1 - q_{ijl}) w_{jl}^2 \right) + \sum_{j=1}^{J} var\left( 2 \sum_{l} w_{jl} q_{ijl} \right)$$

$$= E(\sigma_i^2) + \sum_{j=1}^{J} var(\mu_{ij})$$

$$= \sum_{i=1}^{M} P_i \sigma_i^2 + \sum_{i=1}^{M} P_i \sum_{j=1}^{J} (\mu_{ij} - \mu_j)^2$$

That is, the SNP-specific means are replaced by block-specific means.

*Final remarks*

In practice, most SNPs in the PGS are uncorrelated and there is very little difference between models B and C. Model B is somewhat simpler to use in practice since it only requires the SNP allele frequencies by population. The results in the manuscript assume model B, and the script provided assumes this.

Model C can also be implemented straightforwardly, simply by adding the allele frequencies for SNPs considered in the same block. We also note, however, that model C is still a simplification since genotypes from the same block are assumed to come from a single population.

Model A is the simplest to implement since it only requires the PGS mean by population. However, model B is likely to be the more realistic model. The adjustment to $\sigma$ may be larger or smaller under model A than model B (or C), depending on the pattern of variation of allele frequencies.
