## Supplementary Figures S2-S7 for "Adapting the BOADICEA breast and ovarian cancer risk models for the ethnically diverse UK population"

**Figure S2.** ER-positive predicted proportions by age at cancer diagnosis and ethnicity, based on published estimates from NCRAS data. Age at diagnosis was modelled using quadratic splines with knots placed at 35, 45 and 55 years.

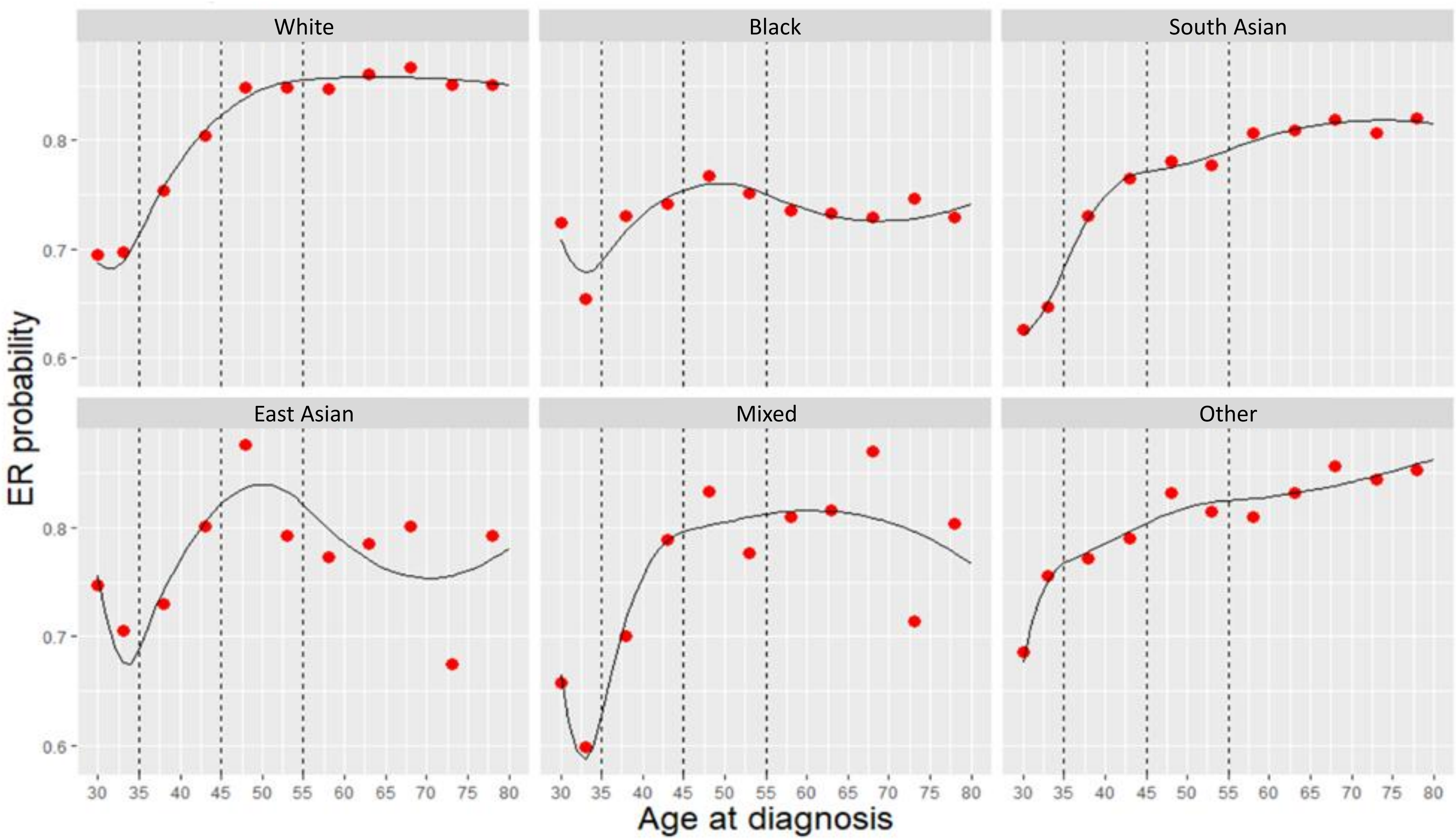

**Figure S3.** ER-negative non-TN predicted proportions by age at cancer diagnosis and ethnicity, based on published estimates from NCRAS data. Age at diagnosis was modelled using quadratic splines with knots placed at 35 and 45 years.

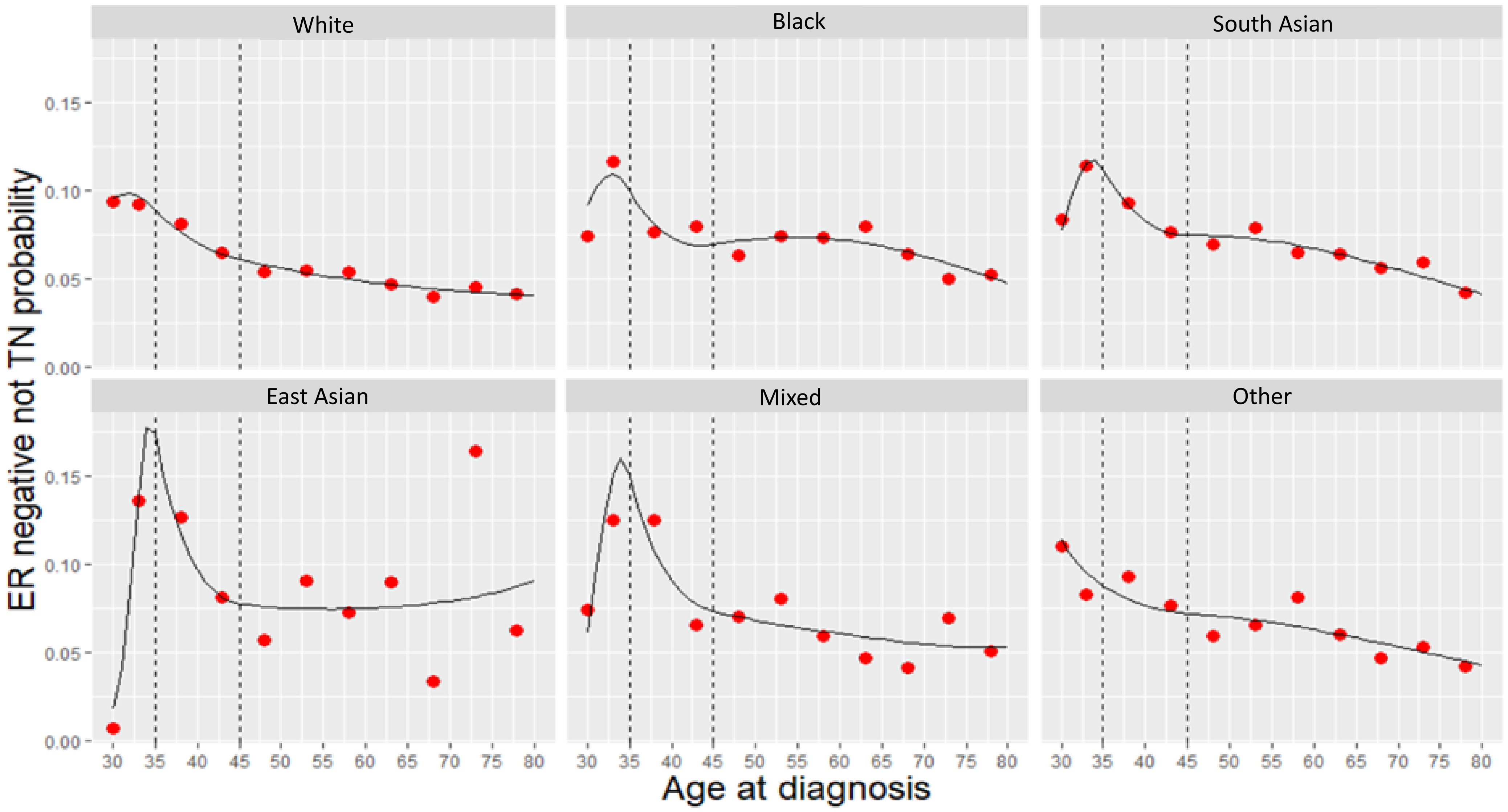

**Figure S4.** TN predicted proportions by age at cancer diagnosis and ethnicity, based on published estimates from NCRAS data. Age at diagnosis was modelled using quadratic splines with knots placed at 35 and 45 years.

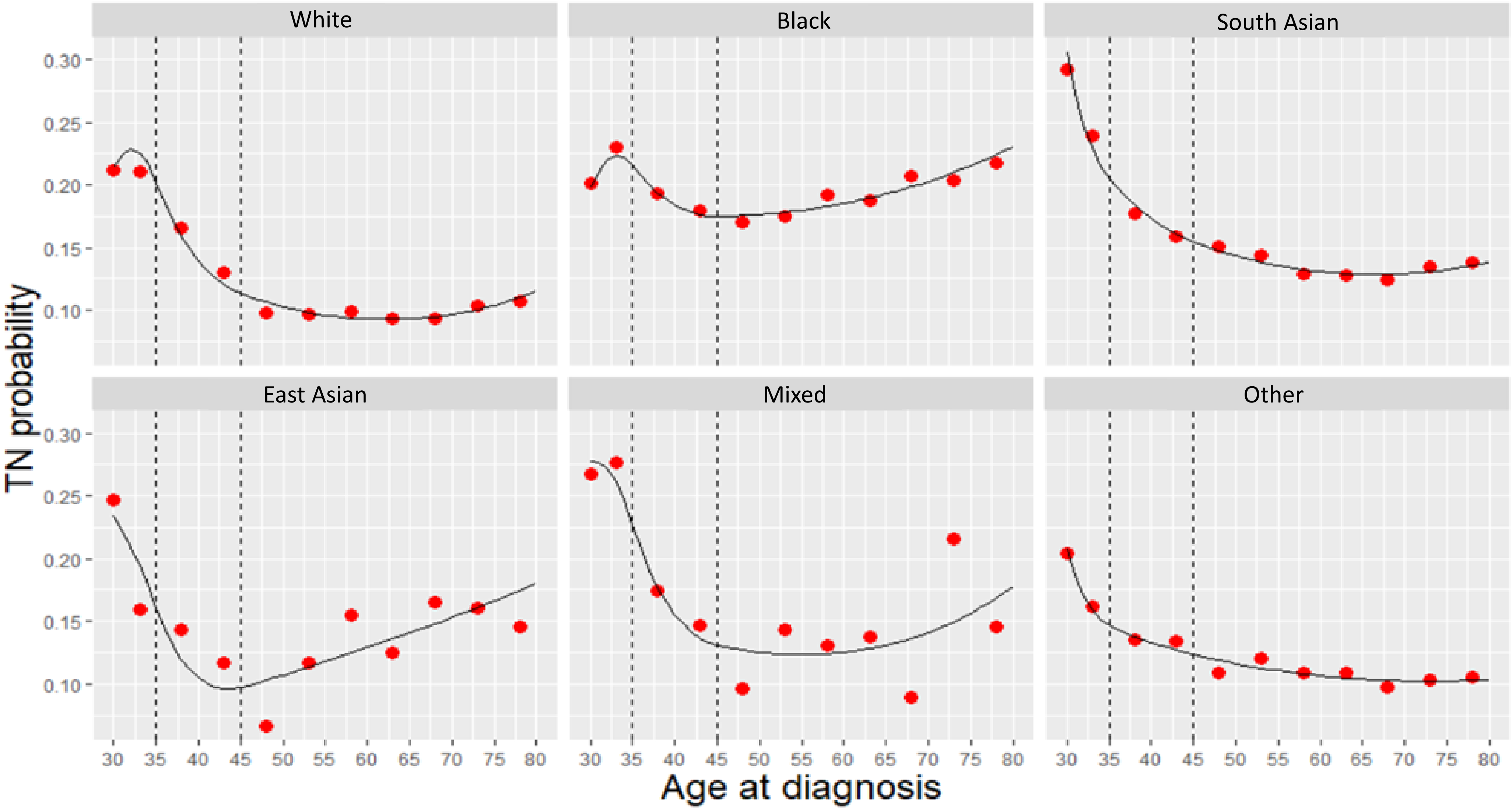

**Figure S5.** ER-positive predicted proportions by age at cancer diagnosis and ethnicity, in PV carriers and in the general population.

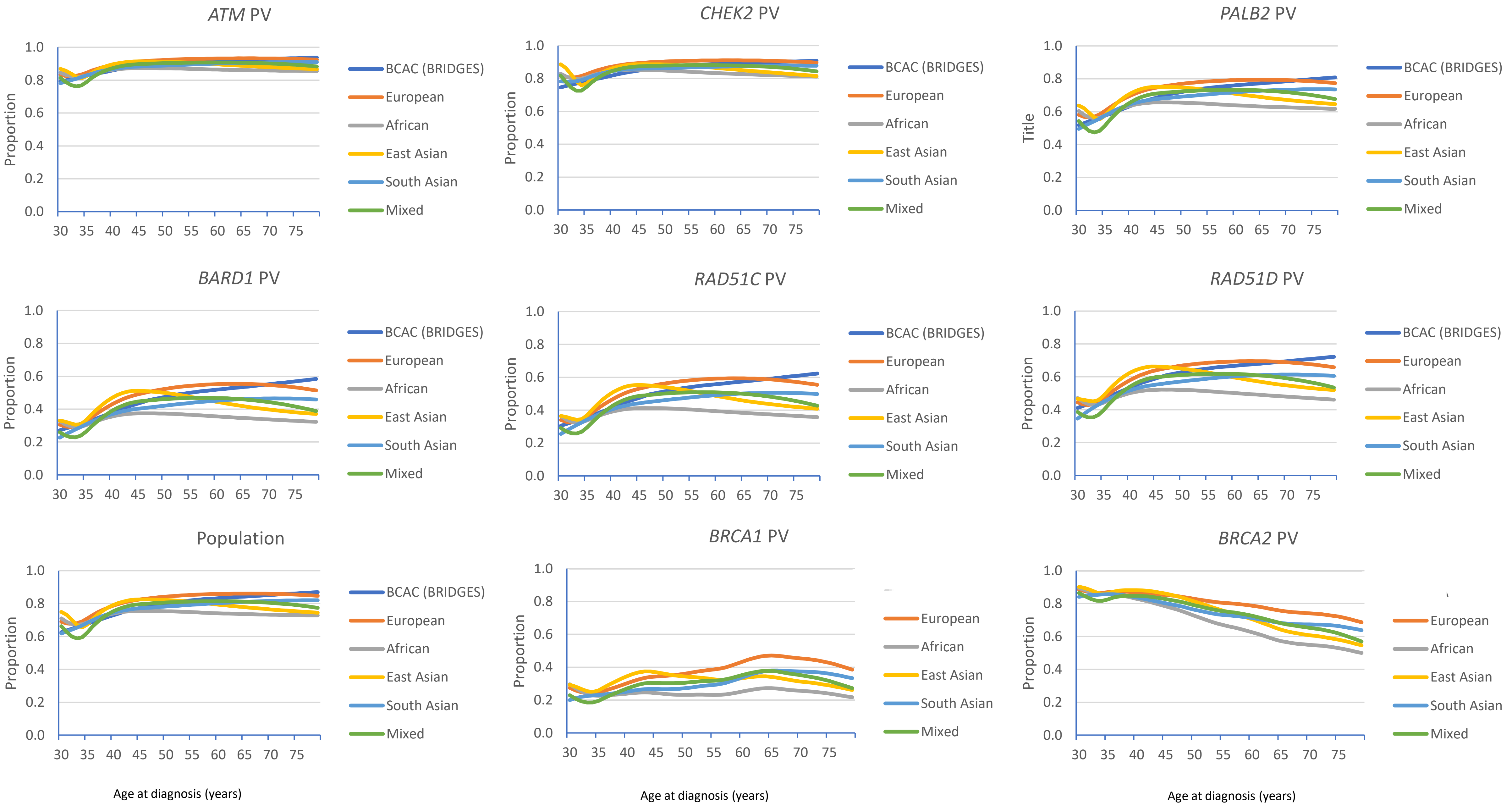

**Figure S6.** ER-negative non-TN predicted proportions by age at cancer diagnosis and ethnicity, in PV carriers and in the general population.

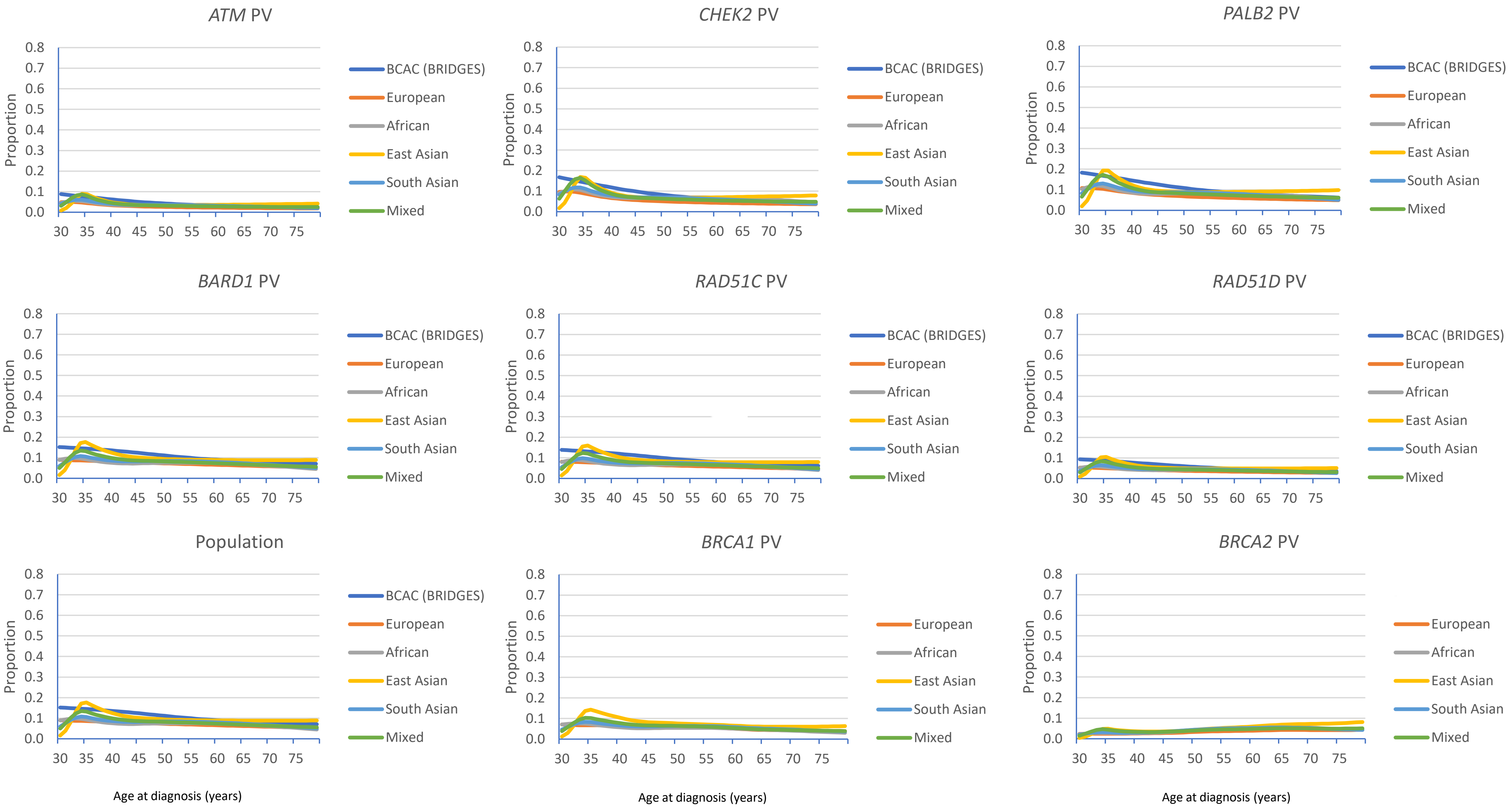

**Figure S7.** TN predicted proportions by age at cancer diagnosis and ethnicity, in PV carriers and in the general population.

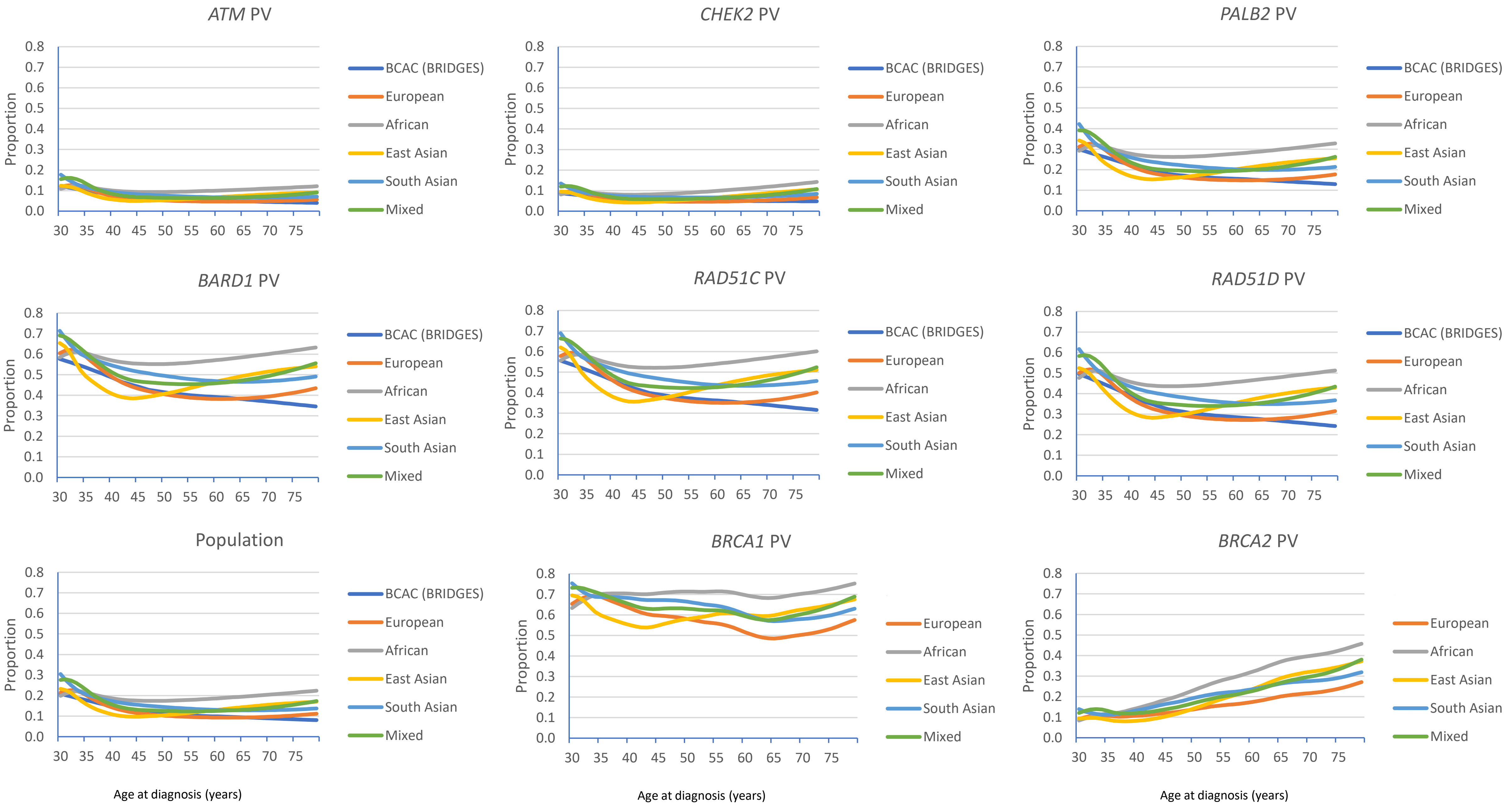
